# Landscape Analysis of Pregnancy Exposure Registries in Low- and Middle-Income Countries: a Scoping Review

**DOI:** 10.1101/2025.09.18.25336113

**Authors:** Niranjan Bhat, Sophie Knudson, Rahmeh AbuShweimeh, Hilma Nakambale, Jessica Mooney, Nancy Salts, Ushma Mehta, Esperança Sevene, Deshayne Fell, Smaragda Lamprianou, Shanthi Narayan Pal, Andy Stergachis

**Author notes:** Corresponding author: Niranjan Bhat, MD MHS, 2201 Westlake Ave., Suite 200, Seattle, WA 98121, USA.

## Abstract

**Introduction:** Drug and vaccine safety information relevant to pregnant individuals is typically insufficient, especially so for persons living in low- and middle-income countries (LMICs). Pregnancy exposure registries (PERs) and similar systems are used to monitor medical products safety. A better understanding of the landscape of PERs in LMICs can support medicines regulatory system strengthening and preparation for new vaccine and drug introductions.

**Objectives:** To identify PERs and related health data collection platforms in LMICs that systematically record pregnancy exposures to medical products and pregnancy outcomes to inform how future efforts, such as new vaccine introductions and treatment programs can better support maternal populations in these countries.

**Design:** Scoping review based on methodology outlined in the Joanna Briggs Institute manual for scoping reviews.

**Data sources:** Electronic search of Medline/PubMed, Embase, CINAHL, and Global Index Medicus in June 2022, and key informants via online survey in July 2022 and interviews.

**Eligibility criteria:** Eligible resources included registries, surveillance systems, and databases that collect information on exposures to medical products during pregnancy and on subsequent maternal, perinatal, and neonatal outcomes in populations located entirely or partially in LMICs. Eligible records were published from January 2000 through June 2022.

**Data extraction and synthesis:** Search results were screened and data extracted using a standardized form by two independent reviewers. Instances of discordance were resolved by a third reviewer. Identified systems were categorized by resource type.

**Results:** A total of 7,515 records from electronic searches were screened, with 396 selected for full-text review and 47 additional records obtained from other sources. From these, 45 data collection systems located in Africa, Asia, and Latin America LMICs were identified, with 36 currently in operation. These resources were grouped into six categories based on structure and approach and summarized according to key features, strengths and weaknesses.

**Conclusions:** This scoping review identified several resources in LMICs dedicated to drug and vaccine safety in pregnancy, but findings indicate that more investment will be needed to ensure such efforts are widespread and sustainable. Understanding the current landscape of such resources in these settings is an important step toward improving safe, world-wide access to life-saving interventions for pregnant populations.

**Study registration:** The protocol for this review has been registered with Open Science Framework (DOI: https://doi.org/10.17605/OSF.IO/FU5AT).

**Article Summary: Strengths and limitations of this study:**

- This analysis documents pregnancy registries and similar systems in low- and middle-income countries for monitoring the safety of drugs and vaccines.
- This scoping review employed a structured search of the published scientific literature, augmented by a grey literature search, online survey and expert consultations.
- Some registries, particularly those without publications or accessible websites, may nevertheless have been missed in this review.
- Registries were not always thorough in reporting the details of their methods, strengths, and limitations in their publications.

## Introduction

Vaccines and drugs have the potential to reduce mortality and morbidity among pregnant individuals, and their offspring, worldwide. However, sufficient information on the safety of drugs and biologic products used during pregnancy is rarely available at the time of licensure.[1,2] To account for this, active safety monitoring during the post-licensure phase strengthens their benefit-risk assessments throughout their product life-cycle. While such safety data are critical for clinical and policy decision-making, they are rarely available from low- and middle-income countries (LMICs), which often have differing background rates of obstetric and infant adverse outcomes, prevalent conditions such as malaria and HIV,[3,4] and limited healthcare access.

Pregnancy exposure registries (PER) are a common type of observational study that systematically collects health information following exposure to medical products during pregnancy, typically after product licensure.[5] PERs are frequently established in high income countries (HICs)[6,7] but are much less common in LMICs, where they typically focus on medicines particularly relevant to the health burden of the populations, such as antiretrovirals and antimalarials.[2,8–10] New vaccines developed specifically for use by pregnant individuals, such as those for respiratory syncytial virus (RSV) and Group B *Streptococcus*, have been approved or are anticipated for introduction in LMICs, but pre-licensure safety data are primarily from HICs with some exceptions.[11–13] To prepare for new medical products, a better understanding of the presence and characteristics of PERs in LMICs is important.

To address this need, an earlier scoping review was conducted to identify existing perinatal data collection systems in LMICs that could support pharmacovigilance in maternal populations, focusing on eight surveillance systems and platforms that held promise for supporting future introductions.[14] Building upon this work, we adopted a complementary and expanded approach that encompasses a broader variety of systems supporting pregnancy safety monitoring in LMICs, designed to inform how future efforts, such as new vaccine introductions and treatment programs, can better support maternal populations in LMICs. The resulting scoping review has now been posted online [15] and is summarized here.

## Methods

The methods and tools for this review have been published.[16] Briefly, a scoping review was conducted to identify pregnancy exposure registries, databases and other systems in LMICs that systematically record exposures to medical products during pregnancy and maternal and infant outcomes. We conducted a systematic search of the scientific and grey literature, supplemented by an online survey and interviews with key informants, as needed. We followed the Joanna Briggs Institute (JBI) manual for scoping reviews, and the search strategy is reported using the Preferred Reporting Items for Systematic reviews and Meta-Analyses extension for Scoping Reviews (PRISMA-ScR) Checklist.[17,18] This protocol was registered with the Open Science Framework.[19]

### Search strategy and information sources

Using an iterative process, the search strategy used predefined criteria for a search in PubMed incorporating controlled vocabulary and free text, as previously described.[16] The full search strategies are provided in Supplemental File 1. An independent information specialist peer reviewed the strategy using the PRESS Checklist.[20] The strategy was then translated to Embase, CINAHL, and WHO’s Global Index Medicus. Reference lists of potentially relevant records and articles were also reviewed.

Additionally, a grey literature search was conducted, encompassing a Google Scholar search and a review of relevant websites, including industry and professional organizations, associations, and alliances; selected public sector agencies (Ministries of Health, regulatory agencies, and pharmacovigilance centers) in LMICs; and selected HIC organizations, academic and other non-governmental groups.

### Study selection and data extraction

Retrieved records were downloaded to EndNote Version 9.3.3 (Clarivate) for de-duplication, following which Covidence was used for screening. Each title and abstract were screened by two independent reviewer authors to determine eligibility. Sources available in the databases used were mainly in the English language, although some records in Spanish and Portuguese were captured. Abstracts and full-texts for these articles were evaluated through automated translation. Disagreements between reviewers were resolved by a third reviewer. Full-text reviews were conducted by two reviewers then selected records proceeded for data extraction. A PRISMA flow diagram[18] was constructed to summarize record disposition. Key information was recorded using a pilot-tested data extraction form and entered into an electronic database (Smartsheet).

### Informant survey, interviews, and expert consultation

An online survey was distributed to experts to identify additional resources in LMICs that may not have been captured, or to provide additional detail for resources already identified. An online survey instrument was developed and shared by members of the WHO Pharmacovigilance Team with counterparts at the WHO regional offices and then delivered on July 7, 2022, to all members of the WHO Programme for International Drug Monitoring ([PIDM](https://www.who.int/teams/regulation-prequalification/regulation-and-safety/pharmacovigilance/health-professionals-info/pidm)),[21] as these are the national pharmacovigilance authorities for their respective countries, and are therefore expected to be knowledgeable about registries in their countries as operators, stakeholders, and/or users of the data. We considered it likely that teams that run safety registries in a country would consult with national pharmacovigilance staff during the establishment or operation of their systems. The survey was also sent to members of the WHO Expert Steering Committee (ESC) on Safety Surveillance in Pregnancy in LMICs. The ESC is a group of experts in maternal immunization safety who are members of the following WHO advisory committees: Strategic Advisory Group of Experts on Immunization (SAGE), Advisory Committee on Safety of Medicinal Products (ACSoMP), Global Advisory Committee on Vaccine Safety (GACSV) and Mother and Newborn Information for Tracking Outcomes & Results. ESC members assessed and advised WHO on a number of topics managed by the WHO’s pharmacovigilance team, including this study.

Additional experts were identified to receive the survey based on the published and grey literature review as well as personal referrals. Semi-structured interviews were conducted when additional information about the registries was required. A multi-disciplinary technical working group was established to assist and guide the review, and the protocol and results were reviewed by the ESC. The data extraction form, survey questionnaire, and interview guide are provided in Supplemental File 1.

### Data analysis

Identified resources were summarized in tables according to relevant characteristics, and the selected PERs were further evaluated based on additional questions, i.e., strengths, weaknesses, ability to add new interventions, and ability to combine data with other systems. Criteria to determine whether the described system could be utilized or adapted for future PERs include whether the system was currently operational, designed with flexibility to accommodate new vaccines or drugs, capable of timely identification of adverse events, employ electronic or mobile tools for data collection and reporting, incorporate mechanisms for follow-up and reminders, or had prior use in post-marketing safety surveillance. The ease with which new exposures could be added could also vary based on the complexity of the exposure, such as if multiple doses are given, or if the drug or vaccine can be administered at different locations, or if additional verifications are needed. Geographic coverage was assessed using maps.

### Patient and public involvement

Patients were not involved in this scoping review.

## Results

### Literature search results

A total of 9,016 records were identified in our search; 7,515 records after de-duplication were imported for title and abstract screening and 396 were selected for full-text review (Figure 1). Of these, 156 met criteria for data extraction. Eighteen individuals responded to the survey, yielding 21 potential resources. After excluding duplicates and negative responses, these results were incorporated into the total. Twelve research studies that were stand-alone, time-limited analyses of retrospectively collected clinical data to evaluate maternal and/or infant outcomes following exposure to drugs or vaccines during pregnancy did not meet eligibility and were not included in the analysis.[22–33] An additional 47 records, publications, and other sources were identified through reference lists, websites, and informant interviews, resulting in a total of 203 records with relevant information. These records included multiple papers or other sources that referred to a single resource (PER or similar data collection system), as well as individual papers that described multiple resources. The 203 records yielded 45 relevant systems that met our criteria for inclusion.

**Figure 1.**
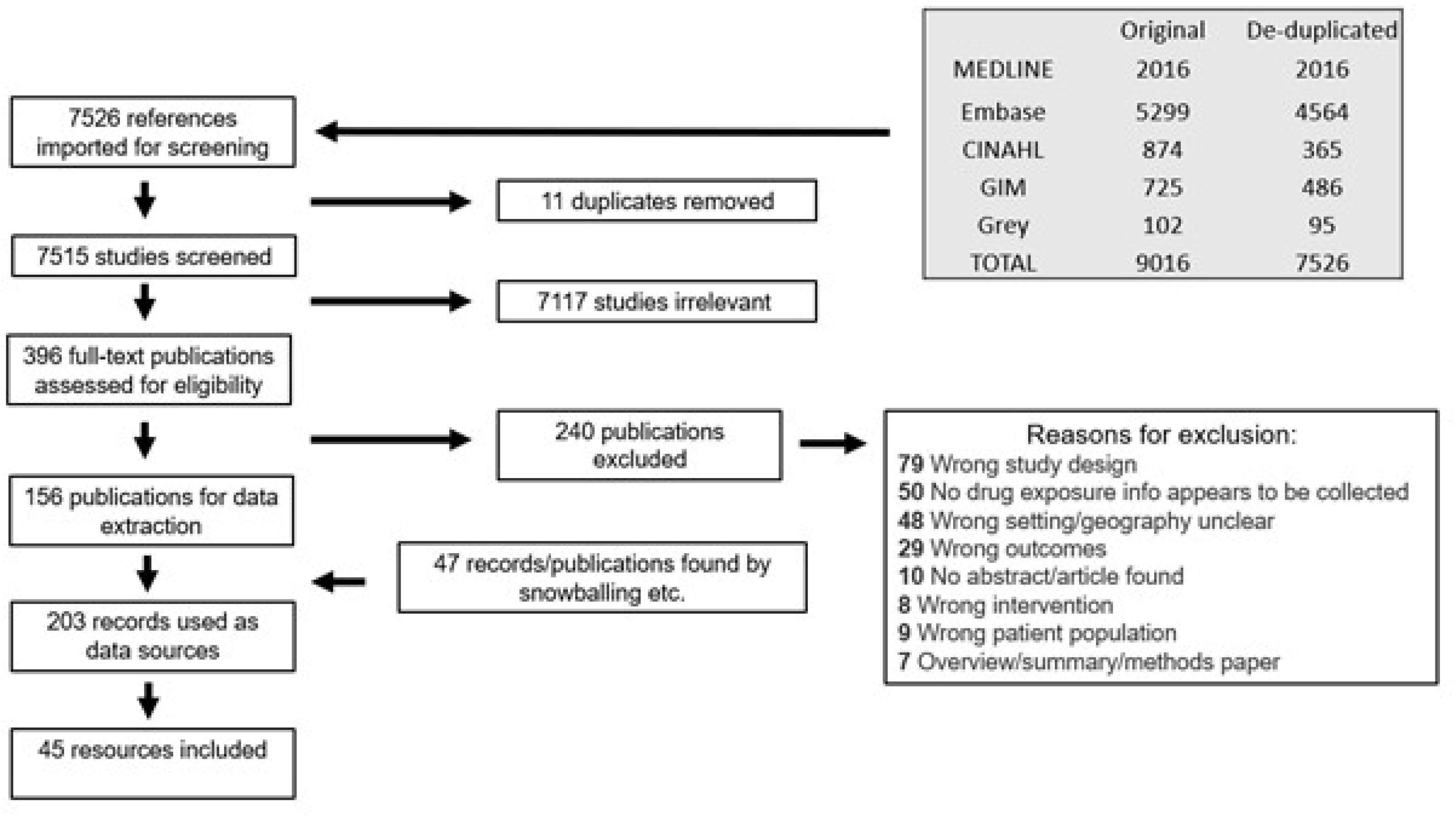
Preferred Reporting Items for Systematic Reviews and Meta-Analyses (PRISMA) flow diagram of records screened and included.**[18]**

### Resource categorization and geographic distribution

Resources were grouped into categories based on broad characteristics, as summarized in Table 1. These systems were distributed across several LMICs, as demonstrated in Figure 2. More than one resource was identified in some countries; conversely, some resources were found to operate in multiple countries. The thirty-six resources that were determined to be currently active are summarized in Table S1 (in Supplemental File 1) and are described in further detail in the following sections according to the resource categories in which they were grouped.

**Figure 2.**
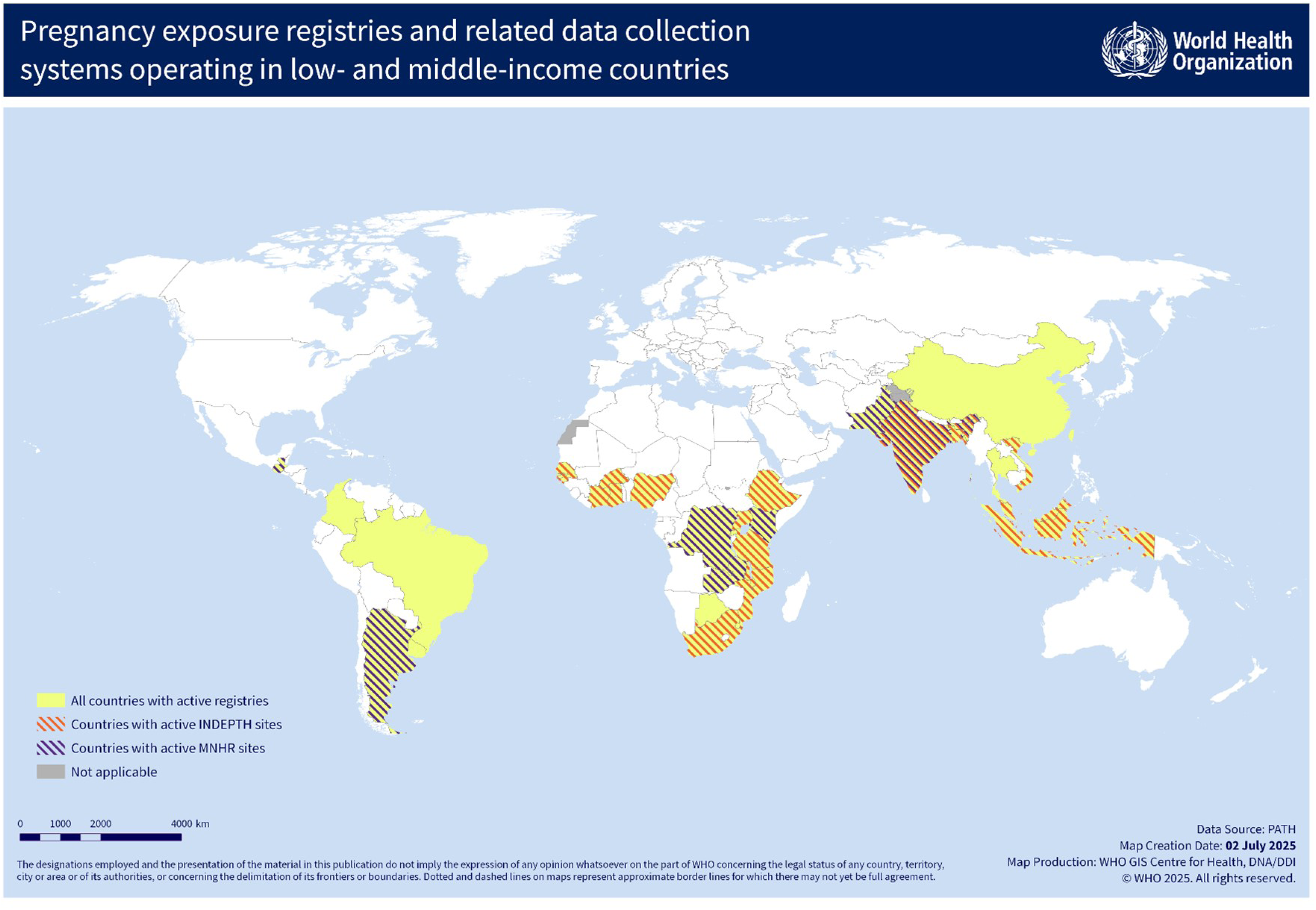
Low- and middle-income countries with active pregnancy exposure registries and related data collection systems.

**Table 1.** Categorization of the pregnancy exposure registries and other resources identified through study methods.

| **Resource Category** | **Brief Description** | **Number of Resources (Number currently active)** |
| --- | --- | --- |
| Pregnancy exposure registries | Self-designated PERs with prospective enrollment and a stated aim to record exposures and outcomes | 11 (7) |
| Health and demographic surveillance systems and other observational cohorts | Population-based cohorts with prospective collection of clinical and epidemiologic data | 7 (7) |
| Outcomes-based registries | Registries that focus on outcomes, such as congenital anomalies | 7 (7) |
| Maternal condition-based registries | Registries that enroll pregnant women with specific underlying health conditions | 6 (3) |
| Manufacturer registries | Registries established by a drug or vaccine manufacturer, often for regulatory purposes | 8 (6) |
| Electronic medical record databases and clinical software platforms | Electronic platforms that prospectively record clinical information within a health care institution or system | 6 (6) |
| **Total** |  | **45 (36)** |

### Pregnancy exposure registries

Resources included in this category met the conventional definition of pregnancy exposure registries, (PER) in that they were prospective observational cohorts focused on the enrollment and follow-up of pregnant women who have received one or more specific drug(s) or vaccine(s) of interest.[5] Enrollment typically occurs before exposure or at least before any pregnancy outcomes are known. Data are systematically collected on all exposures and outcomes for the woman and child. An unexposed or non-pregnant population may be enrolled for comparison, allowing calculation of risk for specific health events.

The majority of PERs in LMICs are currently active and located in sub-Saharan Africa. Many PERs have been in operation for fewer than five years with the exception of the Western Cape Pregnancy Exposure Registry (now a contributing site to UBOMI BUHLE).[34,35] Nine of the PERs are funded through public or donor sources and are run by academic or non-governmental organizations (NGOs).

Most PERs focus on antiretroviral drugs, with a minority assessing antimalarials, COVID-19 therapeutics, or vaccines. In some instances, products of interest being monitored are not fixed; for example, one registry, Malaria in Mothers and Babies (MiMBa) Pregnancy Exposure Registry,[36,37] added COVID-19 vaccinations during the pandemic. Sites of operation extend from single hospitals to multi-national networks, and sample sizes range from less than 500 to more than 25,000 participants accumulated over time. As seen in Table S1, some registries enroll a comparison group such as non-pregnant women of childbearing age, while others enroll only pregnant women, but follow participants exposed and unexposed to the interventions of interest.

The amount of detail provided in the available publications and other reports regarding which outcomes were monitored varied; for instance, conditions such as preterm birth or congenital malformations were not always precisely defined or classified. Most PERs only follow the infants through the neonatal period, with a focus on identifying congenital anomalies, but a small number follow through one year or more to follow growth and development or detect late-appearing congenital anomalies.

Pregnancy exposure registries were among the stronger study designs identified in this scoping review. Key features include systematic, prospective data collection; a focus on pregnant populations; and an emphasis on specific exposures of interest. Enrollment before outcomes are known avoids the risk of recall and reporting biases, allows for systematic history review, and permits standardization of methods such as gestational dating and the dose and timing of exposures. The availability of both numerator and denominator data allows calculations of event rates and disease incidence, and comparator groups allow for estimation of risk.

Limitations of PERs include voluntary enrollment, which may bias data toward high-risk pregnancies, and selection bias if refusals are high and participating women have different characteristics from those who do not. Moreover, abnormal outcomes are more likely to be reported than normal outcomes. Limiting enrollment to those attending antenatal care may bias results and diminish the generalizability in some settings. Late disclosure of pregnancy and late initiation of antenatal care limit information regarding the first trimester of pregnancy, gestational age dating, and early pregnancy loss. Finally, home births and migration increase loss to follow-up, which may bias results.

### Health and Demographic Surveillance Systems and other Observational cohorts

Several population-based observational cohorts identified in this analysis engaged in maternal pharmacovigilance, but do not meet the strict definition of a PER. These cohorts are located in countries throughout the LMIC regions, and all are currently active. Most are designated as health and demographic surveillance systems (HDSS), indicating that they collect demographic and health events from a geographically defined population, of which pregnant women can be a subset. These sites generally conduct longitudinal surveillance, collecting clinical and epidemiologic data prospectively at regular intervals, usually focused on overall health rather than specific exposures. Most of these systems may have the ability to conduct pharmacovigilance in pregnant participants, yet this report only includes those that have published on this topic. A majority of the HDSS resources identified in this review are members of INDEPTH (http://www.indepth-network.org/)[19], a network that currently encompasses 42 independent health research centers and 49 field sites in 19 LMICs.

The Maternal Newborn Health Registry (MNHR) is another large observational cohort with sites in 7 LMICs[38]. The MNHR is a prospective, population-based research registry that collects data to assess trends in pregnancy outcomes and inform research studies within the network.[39] The MNHR primarily monitors the outcomes of maternal mortality, neonatal mortality, and stillbirth. Specific sites in INDEPTH and MNHR were recently evaluated for their capabilities to conduct safety surveillance following immunization in pregnancy.[40]

The Child Health and Mortality Prevention Surveillance Network (CHAMPS) Pregnancy Surveillance is a cohort study operating at HDSS sites in several LMICs. Established within the infrastructure of the main CHAMPS study (described separately below), this evaluation is modeled after the MNHR and enrolls prospectively and retrospectively to monitor for major birth outcomes, including mortality. Two additional observational cohorts focus on maternal pharmacovigilance, including the Shoklo Malaria Research Unit,[41–43] based on the Thai-Myanmar border, and PREPARE,[44] created in preparation for planned clinical trials of a candidate maternal vaccine against Group B *Streptococcus*.

The resources in this section pay particular attention to maternal outcomes, but also capture major infant outcomes, with a few extending into the first year of life. Major obstetric outcomes (including spontaneous abortion, stillbirth, and maternal death) are universally recorded, but some groups capture a larger range of maternal and/or newborn clinical conditions. Most infant outcomes captured in these systems derive from the early neonatal period, including preterm birth, low birth weight, and congenital anomalies.

Also included in this category is IeDEA (International Epidemiology Databases to Evaluate AIDS), an international research and data exchange consortium that combines observational cohort datasets from over 2.2 million people in 44 countries living with and at risk for HIV. This collaboration combines data from HIV programs to study antiretroviral treatment, including pregnant individuals. The group has published studies from Brazil,[45,46] Malawi,[47] South Africa,[48] and the West African region.[49]

### Outcomes-based registries

Outcomes-based registries focus on the capture of specific outcomes, such as congenital malformations. These studies may be open to an entire hospital catchment area, or may target certain groups, such as those receiving antiretrovirals. These systems typically enroll at the time of birth, where exposures are identified from clinical records or via retrospective methods. Some registries involve a single visit while others follow infants for longer periods, and some may include unaffected infants as a comparison group. A single visit with a focus on examination and classification of surface anomalies allows them to efficiently enroll a large sample size, while longer monitoring can identify late-appearing congenital conditions or monitor growth and development. Some registries that only enroll pregnancies resulting in a live birth do not capture all maternal outcomes (including miscarriages and stillbirths). Unaffected mother-infant pairs from the screened population can be compared with affected pairs to assess for risks associated with exposures of interest.

Follow-up care and support to families may be important roles of the registry. In addition, several birth defects surveillance programs in Africa have joined together to form the sub-Saharan African Congenital Anomalies Network (sSCAN) [50] to provide a forum for technical support and collaboration through resource sharing and workshops.

Most registries identified in sub-Saharan Africa focus on antiretroviral drugs, while many in East Asia and South America assess incidence rates of congenital anomalies in the broader population and do not emphasize particular interventions or exposures. A limited number (e.g., in Uganda, Eswatini, and Botswana) were established to assess the teratogenic risks of specific antiretroviral medications such as dolutegravir and cabotegravir. In addition, the CHAMPS study is uniquely focused on stillbirths and neonatal deaths in LMICs.[51,52]

### Maternal condition-based registries

Registries in this category monitor the safety of treatments given to pregnant women with underlying health conditions such as epilepsy, HIV, or cardiac disease. These registries may emphasize specific infant outcomes due to a known or suspected safety signal associated with an individual drug or class of pharmacologic products. Some also monitor maternal outcomes associated with the condition, such as seizure frequency or worsening heart failure. Such registries are more likely to be prospective, as participants are already being followed. While registries operated by a single clinic or hospital enroll smaller populations, even national or multi-national registries generally do not reach large sample sizes. Unexposed pregnant women or exposed non-pregnant women with these conditions may be enrolled as comparator groups.

Outside of HIV-specific registries, systems identified in this category are more commonly located in middle-income countries such as India, Brazil, and Argentina, which may reflect that physicians in wealthier countries are able to assemble larger patient cohorts in fields such as neurology or cardiology.

### Manufacturer-initiated registries

Registries in this category are usually funded and owned by the manufacturer of the drug or vaccine of interest, and either focus on an individual drug or vaccine, or can monitor all products in the same therapeutic class as the manufacturer’s product. The US Food and Drug Administration (FDA) and the European Medicines Agency (EMA) recommend registries for products likely to be used during pregnancy, [20,21] and these systems are often established to comply with a regulatory requirement.

They can contribute to benefit-harm assessments, support product labeling, and inform guidance throughout a medical product’s life-cycle. Most manufacturer registries focus on high-income markets,[22] but participants from other areas of the world may also be included. Those that indicated some recruitment from LMICs have been included in this review. Active surveillance studies and other research efforts intended to answer specific safety questions that are usually classified as Phase 4 studies were not included.

The registries identified are operated by range of groups, including contract research organizations, academic groups, and the manufacturer themselves. Some use passive surveillance and receive sporadic reports of exposures directly from patients or providers, although some may conduct outreach. Instructions on how to enroll are provided in the product literature or company website. The voluntary nature of enrollment may affect completeness, but once a participant is enrolled, the registries maintain contact to solicit outcomes. Some registries are designed for a specific scientific question and thus may limit the timeframe or sample size, while others appear to operate indefinitely.

### Electronic health records databases and clinical software platforms

Systems were included in this review if a published study or report relied on an electronic clinical care database or software platform. In these cases, investigators may have developed programming to identify pregnancies, extract data, and classify outcomes appropriately. Electronic health records (EHR) databases and clinical software platforms typically record information prospectively on all patients within a health care system.

EHR platforms may be implemented by individual facilities, health systems or sectors, or nationally. By collecting and storing data electronically, databases can be searched and analyzed using structured, reproducible methods. Some EHRs may be deployed in targeted facilities, such as maternal and newborn clinics, or pregnant women and their infants may represent a subset of the total covered population. Linking records across time and databases allows identification of large numbers of relevant exposures and outcomes. Identified databases include SmartCare in Zambia, the Baobab Health Antiretroviral Therapy (BART) system in Malawi, and the Provincial Health Data Centre (PHDC) system in the Western Cape province of South Africa.

Software platforms, such as DHIS2, comprise another set of systems. DHIS2 is an open-source web-based software platform that collects and analyzes data at the population and individual levels, and can be designed for facilities, health systems, or national programs. By the end of 2022, DHIS2 was used in more than 75 LMICs, with 69 countries using DHIS2 at a national scale.[167,168] DHIS2 can provide for individual patient care and conduct epidemiological analyses.[169] Data are entered per routine care rather than through studies, and therefore capture multiple aspects of healthcare. Specific health topics such as HIV, tuberculosis, or immunizations can be analyzed through pre-configured metadata packages. For instance, the Reproductive, Maternal, Newborn, Child and Adolescent Health (RMNCAH) package can track exposures and outcomes during pregnancy. This review identified one clear example in Palestine where DHIS2 was used to track maternal exposures and outcomes,[49] but other deployments may exist.

Other open-source clinical software platforms, such as OpenMRS and OpenEMR, could provide similar functionality.[11] However, no publications or online sources describing their use for monitoring drug safety during pregnancy were found in our search. The Perinatal Information System (Sistema Informatico Perinatal (SIP)) is a free standardized perinatal clinical record run by PAHO’s Latin American Center for Perinatology/Women’s Health and Reproductive Health (CLAP/WR).[170,171] Facilities throughout Latin America and the Caribbean use this resource to produce reports and combine data across facilities, conducting studies at the regional or national level.[171–173] While no publications related specifically to pharmacovigilance using SIP were identified, the capability appears possible.

One study analyzed data from a national insurance claims database in China to assess the safety of medication use in pregnancy. This approach is common in HICs where methods are well developed and coverage is high. While national health insurance is expanding in LMICs [174][175] this growth has been limited in the lower-resource areas of Africa and Southeast Asia. Research using these databases is limited[176–179] and none have involved maternal pharmacovigilance, but opportunities may arise as health insurance programs become more widespread.

Table 2 lists key features of each resource type, highlighting the relative advantages and disadvantages of specific designs.

**Table 2.**
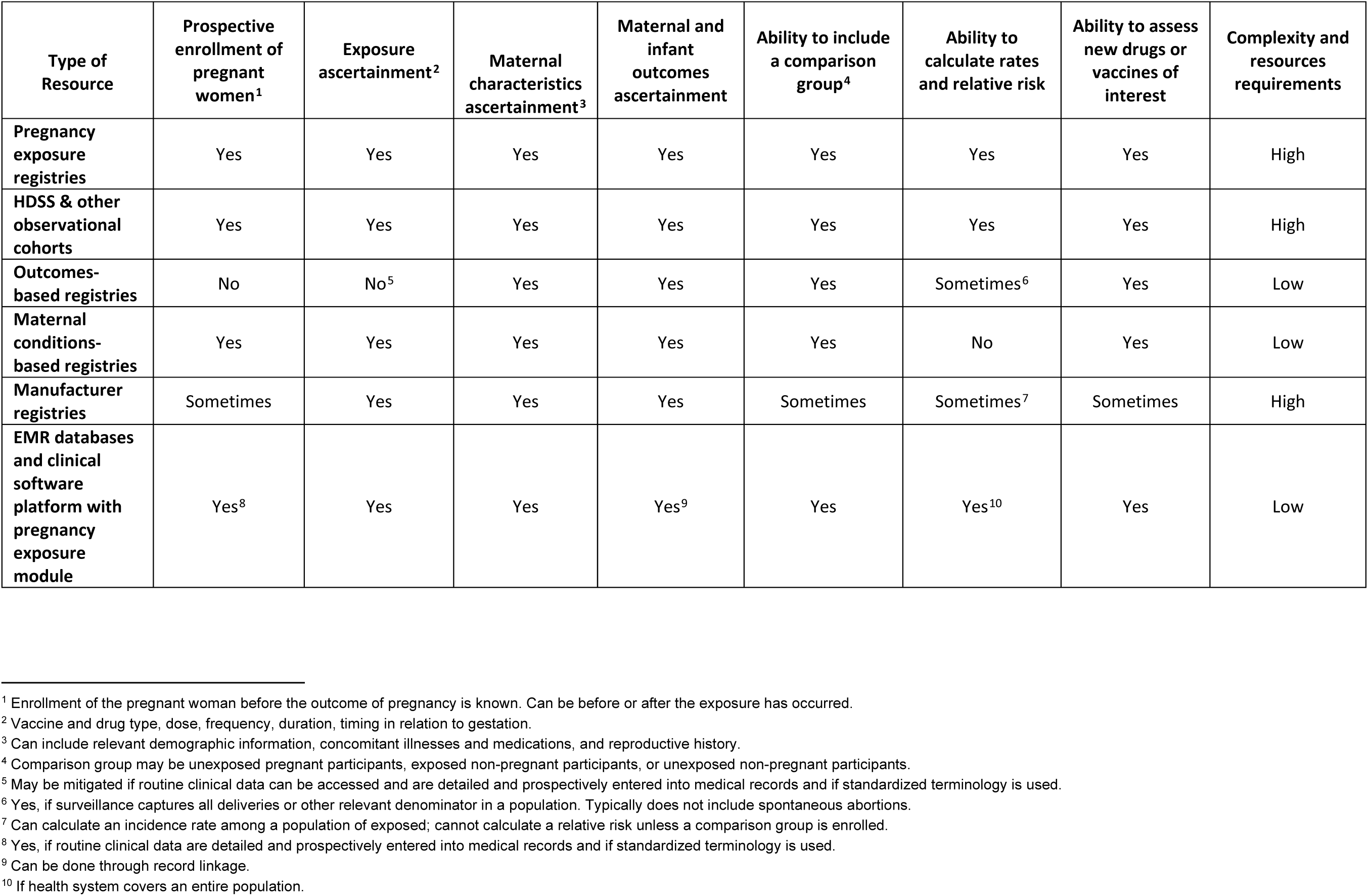
Key features of pregnancy exposure registries and related data collection systems, by resource type.

| Type of Resource | Prospective enrollment of pregnant women<sup>1</sup> | Exposure ascertainment<sup>2</sup> | Maternal characteristics ascertainment<sup>3</sup> | Maternal and infant outcomes ascertainment | Ability to include a comparison group<sup>4</sup> | Ability to calculate rates and relative risk | Ability to assess new drugs or vaccines of interest | Complexity and resources requirements |
| --- | --- | --- | --- | --- | --- | --- | --- | --- |
| **Pregnancy exposure registries** | Yes | Yes | Yes | Yes | Yes | Yes | Yes | High |
| **HDSS & other observational cohorts** | Yes | Yes | Yes | Yes | Yes | Yes | Yes | High |
| **Outcomes-based registries** | No | No<sup>5</sup> | Yes | Yes | Yes | Sometimes<sup>6</sup> | Yes | Low |
| **Maternal conditions-based registries** | Yes | Yes | Yes | Yes | Yes | No | Yes | Low |
| **Manufacturer registries** | Sometimes | Yes | Yes | Yes | Sometimes | Sometimes<sup>7</sup> | Sometimes | High |
| **EMR databases and clinical software platform with pregnancy exposure module** | Yes<sup>8</sup> | Yes | Yes | Yes<sup>9</sup> | Yes | Yes<sup>10</sup> | Yes | Low |

<sup>1</sup> Enrollment of the pregnant woman before the outcome of pregnancy is known. Can be before or after the exposure has occurred.
<sup>2</sup> Vaccine and drug type, dose, frequency, duration, timing in relation to gestation.
<sup>3</sup> Can include relevant demographic information, concomitant illnesses and medications, and reproductive history.
<sup>4</sup> Comparison group may be unexposed pregnant participants, exposed non-pregnant participants, or unexposed non-pregnant participants.
<sup>5</sup> May be mitigated if routine clinical data can be accessed and are detailed and prospectively entered into medical records and if standardized terminology is used.
<sup>6</sup> Yes, if surveillance captures all deliveries or other relevant denominator in a population. Typically does not include spontaneous abortions.
<sup>7</sup> Can calculate an incidence rate among a population of exposed; cannot calculate a relative risk unless a comparison group is enrolled.
<sup>8</sup> Yes, if routine clinical data are detailed and prospectively entered into medical records and if standardized terminology is used.
<sup>9</sup> Can be done through record linkage.
<sup>10</sup> If health system covers an entire population.

## Discussion

This scoping review identified 45 PERs and related systems pertinent to maternal pharmacovigilance in LMICs, 36 of which remain active. These systems cover a range of approaches and areas of focus ranging from conventional PERs to electronic health information systems.

Of these resource types, traditional PERs remain essential for monitoring safety in pregnancy by employing rigorous designs and methods and including comparison groups to calculate risk. However, they are resource-intensive, which can limit their duration, size, and/or geographic coverage. The majority of registries identified focus on antiretrovirals, reflecting the importance of these interventions in many LMIC populations, particularly during pregnancy. Currently active PERs might be adapted to include new exposures of interest, as had been done for COVID-19 vaccines.[36]

HDSS cohorts are widespread throughout LMICs and have a long history of operation.[53] While most are centered around basic vital statistics reporting, some have incorporated additional activities and investment to identify pregnancies early, perform gestational dating, ensure accurate exposure reporting, and capture maternal and infant outcomes more completely. Many sites include community surveillance, and thus can monitor pregnancies and deliveries outside health facilities. These systems can often establish comparator populations, determine background rates, and calculate relative risk estimates. Ultimately, these sites require substantial resources and community engagement but provide the expertise and infrastructure necessary to conduct pharmacovigilance studies in pregnant populations.

Outcomes-based registries are ideal to detect uncommon outcomes but are often streamlined and follow participants only when outcomes of interest are detected. Efforts may be required to avoid incompleteness or recall bias due to retrospective data collection, and they might not always capture early obstetric events. Electronic medical record systems represent another avenue that integrates activities within routine clinical care. However, quality relies on training and oversight, and analyses benefit from standardized terminologies and methods. The addition of maternal pharmacovigilance could require a comparatively small incremental investment.

This scoping review employed a structured comprehensive search of published scientific literature, but was subject to some limitations. A number of registries, particularly in LMICs, may not have published their data and may have a minimal online presence. In addition, the databases selected for our search may not have contained all relevant publications. At the time of designing our search strategy, we considered that additional databases such as the Web of Science and Scopus would likely overlap considerably with our chosen data sources. However, these repositories might include some unique references and could be considered for future reviews. Even when publications or websites are available, registries were not always thorough in reporting the details of their methods, strengths, and limitations. Our literature search was augmented by grey literature, surveys, expert consultations, and outreach to a limited number of registry informants to account for these issues, however some resources may still have been missed or incompletely characterized.

This landscape identified several resources dedicated to drug and vaccine safety in pregnancy in LMICs, but it is clear that more investment is needed to ensure such efforts are inclusive and sustainable. Most of the registries identified could be strengthened or modified to incorporate new exposures, if adequate resources are provided. The inclusion of desirable features, such as early enrollment, prospective data collection, accurate gestational dating, capture of maternal events, and enumerated populations to generate denominator-based rates, should be introduced or reinforced wherever possible. However, specific approaches must be matched to their local context, taking into account the questions to be answered, funding available, and existing infrastructure. Registries with flexible design might combine analyses across regions, but such efforts will require a more granular understanding of structure, data variables, and methodologic approaches. One consideration for such an undertaking would be whether the registries use standardized case definitions such as those developed by the Brighton Collaboration and the GAIA initiative to promote harmonization, enhance reproducibility, and ensure scientifically robust assessments of safety. Other design elements, such as the duration of follow-up and the degree of detail in describing the exposure would need to be matched. In the situation where multiple registries are operating in the same region, procedures would need to be established to identify individuals with overlapping participation, perhaps by developing data queries that search for records with matching elements such as name, date of birth, and contact information. Ultimately, improving the safe access of pregnant populations in LMICs to life-saving drugs and vaccines must begin with a clear understanding of where the greatest needs are in order to develop effective strategies to safeguard against possible harms that may be associated with their use.

## Acknowledgements

The authors would like to thank Pierre Buekens, Jessica Fleming, Lauren Newhouse, Sonia Chaabane, Becky Skidmore, Kate Fay, and Anna Seale for their advice and support. The authors alone are responsible for the views expressed in this article and they do not necessarily represent the views, decisions or policies of the institutions with which they are affiliated.

## Author Contributions

NB was responsible for conception and design of the review. UM, DF, SL, SNP, and AS provided expert review and input into the protocol. SK, RA, HN, and JM conducted the initial screening of records, with NB and AS are second reviewers. HN, JM, and NS conducted the full text reviews and data extraction, and NB, AS, JM, and HN analyzed the data. NB and AS were responsible for producing the initial draft of the manuscript. All authors provided significant editorial comments on the protocol drafts and read and approved the final manuscript. NB is responsible for the overall content as guarantor, and as such accepts full responsibility for the finished work and/or the conduct of the study, had access to the data, and controlled the decision to publish.

## Funding statement

This work was supported by a grant from the Bill & Melinda Gates Foundation [grant number INV-037810]. Under the grant conditions of the Foundation, a Creative Commons Attribution 4.0 Generic License has already been assigned to the Author Accepted Manuscript version that might arise from this submission.

## Competing interests

None declared.

## Patient and public involvement

Patients and/or the public were not involved in the design, conduct, reporting, or dissemination plans of this research.

## Ethics approval

This scoping review did not involve human participants and therefore did not require approval by an Ethics Committee or Institutional Review Board.

## Patient consent for publication

Not applicable.

## Data availability statement

All data relevant to the study are included in the article, uploaded as supplementary information, or available with the previous publication describing the methods (doi:10.1136/bmjopen-2022-070543). All data underlying the results are available as part of the article and no additional source data are required.

## Supplemental Material

### Search strategies by database

#### PubMed

| Set # | Query | Results |
| --- | --- | --- |
| 132 | #130 NOT #131 | 2,016 |
| 131 | address[Publication Type] OR autobiography[Publication Type] OR bibliography[Publication Type] OR biography[Publication Type] OR comment[Publication Type] OR dictionary[Publication Type] OR directory[Publication Type] OR editorial[Publication Type] OR "expression of concern"[Publication Type] OR festschrift[Publication Type] OR historical article[Publication Type] OR interactive tutorial[Publication Type] OR lecture[Publication Type] OR news[Publication Type] OR newspaper article[Publication Type] OR portrait[Publication Type] OR video-audio media[Publication Type] OR webcast[Publication Type] | 2,050,653 |
| 130 | #128 NOT #129 | 2,048 |
| 129 | Animals[mesh] NOT Humans[mesh] | 5,016,862 |
| 128 | #126 AND #127 | 2,051 |
| 127 | ("2000"[Date - Publication] : "3000"[Date - Publication]) | 20,721,844 |
| 126 | #61 AND #125 | 2,597 |
| 125 | #62 OR #63 OR #64 OR #65 OR #66 OR #67 OR #68 OR #69 OR #70 OR #71 OR #72 OR #73 OR #74 OR #75 OR #76 OR #77 OR #78 OR #79 OR #80 OR #81 OR #82 OR #83 OR #84 OR #85 OR #86 OR #87 OR #88 OR #89 OR #90 OR #91 OR #92 OR #93 OR #94 OR #95 OR #96 OR #97 OR #98 OR #99 OR #100 OR #101 OR #102 OR #103 OR #104 OR #105 OR #106 OR #107 OR #108 OR #109 OR #110 OR #111 OR #112 OR #113 OR #114 OR #115 OR #116 OR #117 OR #118 OR #119 OR #120 OR #121 OR #122 OR #123 OR #124 | 7,704,889 |
| 124 | MNCH[Other Term] AND (maternal*[Other Term] OR newborn*[Other Term] OR child*[Other Term]) | 11 |
| 123 | MNCH[Title] AND (maternal*[Title] OR newborn*[Title] OR child*[Title]) | 22 |
| 122 | "Maternal Health"[mesh] | 2,142 |
| 121 | "Child Health"[mesh] | 4,447 |
| 120 | "Infant Health"[mesh] | 1,175 |

|  |  |  |
| --- | --- | --- |
| 119 | (birth[Other Term] OR births[Other Term]) AND defect*[Other Term] | 1,029 |
| 118 | (birth[Title] OR births[Title]) AND defect*[Title] | 2,671 |
| 117 | (birth[Other Term] OR births[Other Term] OR matern*[Other Term] OR neonat*[Other Term] OR neo-nat*[Other Term] OR perinatal*[Other Term] OR peri-natal*[Other Term] OR peripartum[Other Term] OR "peri-partum"[Other Term] OR postnatal*[Other Term] OR post-natal*[Other Term] OR postpartum[Other Term] OR "post-partum"[Other Term] OR post-birth*[Other Term] OR pregnanc*[Other Term]) AND outcome*[Other Term] | 9,426 |
| 116 | (birth[Title] OR births[Title] OR matern*[Title] OR neonat*[Title] OR neo-nat*[Title] OR perinatal*[Title] OR peri-natal*[Title] OR peripartum[Title] OR "peri-partum"[Title] OR postnatal*[Title] OR post-natal*[Title] OR postpartum[Title] OR "post-partum"[Title] OR post-birth*[Title] OR pregnanc*[Title]) AND outcome*[Title] | 31,066 |
| 115 | Pregnancy Outcome[mesh] | 82,242 |
| 114 | (fetal[OT] OR foetal[OT] OR fetus*[OT] OR foetus*[OT] OR prenatal*[OT] OR pre-natal*[OT]) AND expos*[OT] | 2,033 |
| 113 | (fetal[Title] OR foetal[Title] OR fetus*[Title] OR foetus*[Title] OR prenatal*[Title] OR pre-natal*[Title]) AND expos*[Title] | 10,858 |
| 112 | "Prenatal Exposure Delayed Effects"[mesh] | 33,016 |
| 111 | (fetal[Other Term] OR foetal[Other Term] OR fetus*[Other Term] OR foetus*[Other Term]) AND develop*[Other Term] | 2,531 |
| 110 | (fetal[Title] OR foetal[Title] OR fetus*[Title] OR foetus*[Title]) AND develop*[Title] | 8,734 |
| 109 | "Fetal Development"[mesh:noexp] | 9,246 |
| 108 | antiretroviral*[Other Term] OR anti-retroviral*[Other Term] OR "anti-HIV"[Other Term] OR "anti-AIDS"[Other Term] OR "AIDS drug"[Other Term] OR "AIDS drugs"[Other Term] | 7,307 |
| 107 | (antiretroviral*[Title] OR anti-retroviral*[Title] OR "anti-HIV"[Title] OR "anti-AIDS"[Title] OR "AIDS drug"[Title] OR "AIDS drugs"[Title]) | 29,826 |
| 106 | "Anti-Retroviral Agents"[mesh] | 69,532 |
| 105 | "Zika Virus Infection/prevention and control"[MeSH] | 1,096 |
| 104 | "Streptococcal Infections/prevention and control"[MeSH:noexp] | 2,944 |

|  |  |  |
| --- | --- | --- |
| 103 | "Respiratory Syncytial Virus Infections/prevention and control"[MeSH Terms] | 1,832 |
| 102 | "Hepatitis E/prevention and control"[MeSH Terms] | 346 |
| 101 | "Cytomegalovirus Infections/prevention and control"[MeSH Terms] | 3,130 |
| 100 | antimalarial*[Other Term] OR anti-malarial*[Other Term] | 2,577 |
| 99 | antimalarial*[Title] OR anti-malarial*[Title] | 8,562 |
| 98 | Antimalarials[mesh] | 28,226 |
| 97 | "Malaria/prevention and control"[MeSH Terms] | 17,528 |
| 96 | (maternal[Other Term] OR pregnan*[Other Term]) AND (immunisation*[Other Term] OR immunization*[Other Term]) | 754 |
| 95 | (maternal[Title] OR pregnan*[Title]) AND (immunisation*[Title] OR immunization*[Title]) | 1,153 |
| 94 | (antibod*[Other Term] OR anti-bod*[Other Term]) AND transfer*[Other Term] | 338 |
| 93 | (antibod*[Title] OR anti-bod*[Title]) AND transfer*[Title] | 1,620 |
| 92 | immunit*[Other Term] AND maternally-acqui*[Other Term] | 8 |
| 91 | immunit*[Title] AND maternally-acqui*[Title] | 8 |
| 90 | immunit*[Other Term] AND transfer*[Other Term] | 161 |
| 89 | immunit*[Title] AND transfer*[Title] | 882 |
| 88 | "Immunity, Maternally-Acquired"[mesh] | 5,842 |
| 87 | "Immunization Programs"[mesh:noexp] | 12,435 |
| 86 | Immunization[mesh:noexp] | 144,668 |
| 85 | vaccin*[ti] OR vaccin*[ot] | 220,223 |
| 84 | "Viral Vaccines"[mesh] | 132,729 |
| 83 | "Vaccines, Live, Unattenuated"[mesh] | 82 |
| 82 | "Vaccines, Synthetic"[mesh] | 31,589 |
| 81 | "Vaccines, Inactivated"[mesh:noexp] | 6,082 |
| 80 | "Vaccines, Combined"[mesh] | 10,562 |
| 79 | "Vaccines, Attenuated"[mesh] | 12,810 |
| 78 | "Protozoan Vaccines"[mesh] | 6,894 |
| 77 | "Bacterial Vaccines"[mesh] | 72,365 |

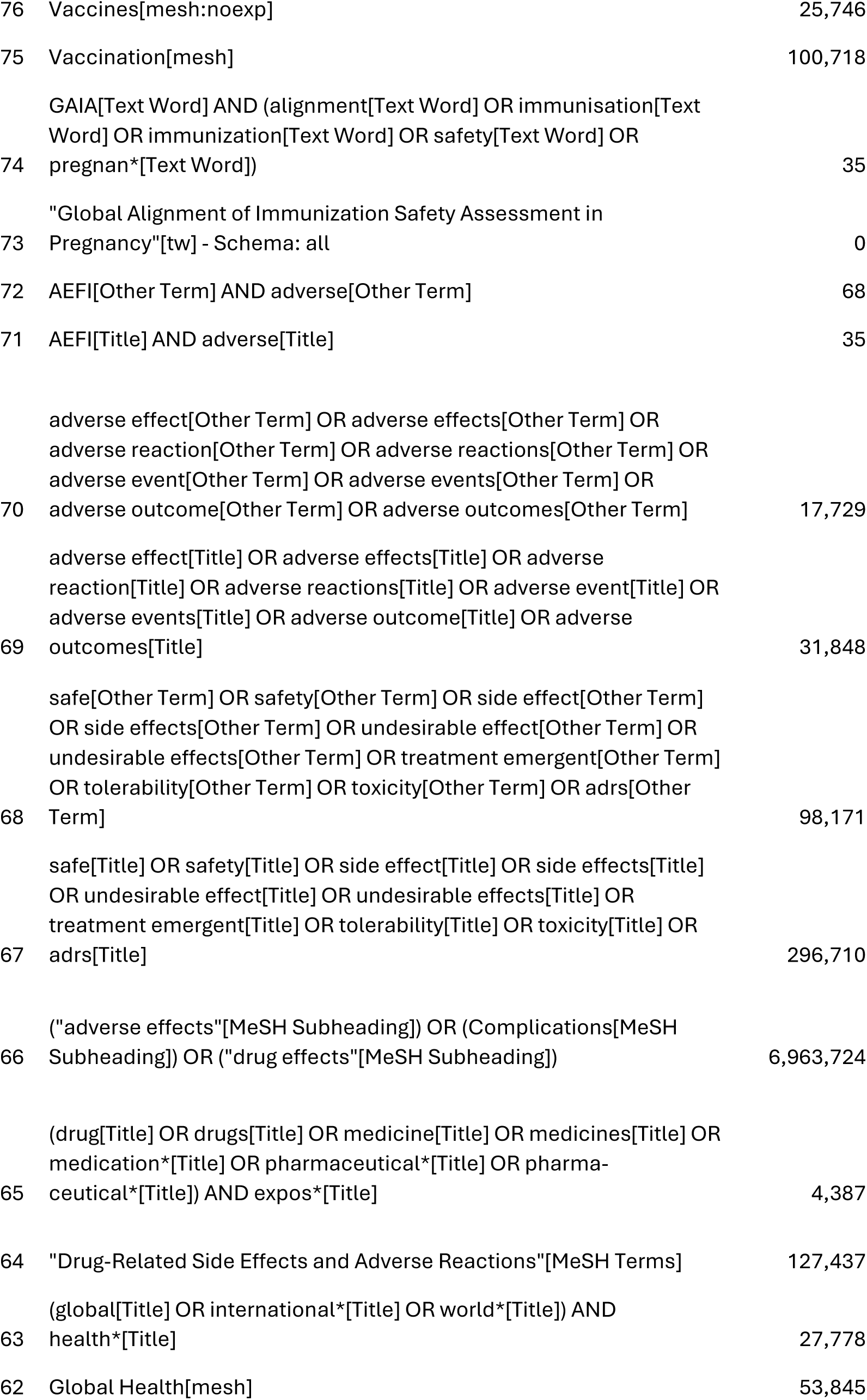

|  |  |  |
| --- | --- | --- |
| 76 | Vaccines[mesh:noexp] | 25,746 |
| 75 | Vaccination[mesh] | 100,718 |
|  | GAIA[Text Word] AND (alignment[Text Word] OR immunisation[Text Word] OR immunization[Text Word] OR safety[Text Word] OR |  |
| 74 | pregnan*[Text Word]) | 35 |
|  | "Global Alignment of Immunization Safety Assessment in |  |
| 73 | Pregnancy"[tw] - Schema: all | 0 |
| 72 | AEFI[Other Term] AND adverse[Other Term] | 68 |
| 71 | AEFI[Title] AND adverse[Title] | 35 |
|  | adverse effect[Other Term] OR adverse effects[Other Term] OR |  |
|  | adverse reaction[Other Term] OR adverse reactions[Other Term] OR |  |
|  | adverse event[Other Term] OR adverse events[Other Term] OR |  |
| 70 | adverse outcome[Other Term] OR adverse outcomes[Other Term] | 17,729 |
|  | adverse effect[Title] OR adverse effects[Title] OR adverse |  |
|  | reaction[Title] OR adverse reactions[Title] OR adverse event[Title] OR |  |
|  | adverse events[Title] OR adverse outcome[Title] OR adverse |  |
| 69 | outcomes[Title] | 31,848 |
|  | safe[Other Term] OR safety[Other Term] OR side effect[Other Term] |  |
|  | OR side effects[Other Term] OR undesirable effect[Other Term] OR |  |
|  | undesirable effects[Other Term] OR treatment emergent[Other Term] |  |
|  | OR tolerability[Other Term] OR toxicity[Other Term] OR adrs[Other |  |
| 68 | Term] | 98,171 |
|  | safe[Title] OR safety[Title] OR side effect[Title] OR side effects[Title] |  |
|  | OR undesirable effect[Title] OR undesirable effects[Title] OR |  |
|  | treatment emergent[Title] OR tolerability[Title] OR toxicity[Title] OR |  |
| 67 | adrs[Title] | 296,710 |
|  | ("adverse effects"[MeSH Subheading]) OR (Complications[MeSH |  |
| 66 | Subheading]) OR ("drug effects"[MeSH Subheading]) | 6,963,724 |
|  | (drug[Title] OR drugs[Title] OR medicine[Title] OR medicines[Title] OR |  |
|  | medication*[Title] OR pharmaceutical*[Title] OR pharma- |  |
| 65 | ceutical*[Title]) AND expos*[Title] | 4,387 |
| 64 | "Drug-Related Side Effects and Adverse Reactions"[MeSH Terms] | 127,437 |
|  | (global[Title] OR international*[Title] OR world*[Title]) AND |  |
| 63 | health*[Title] | 27,778 |
| 62 | Global Health[mesh] | 53,845 |

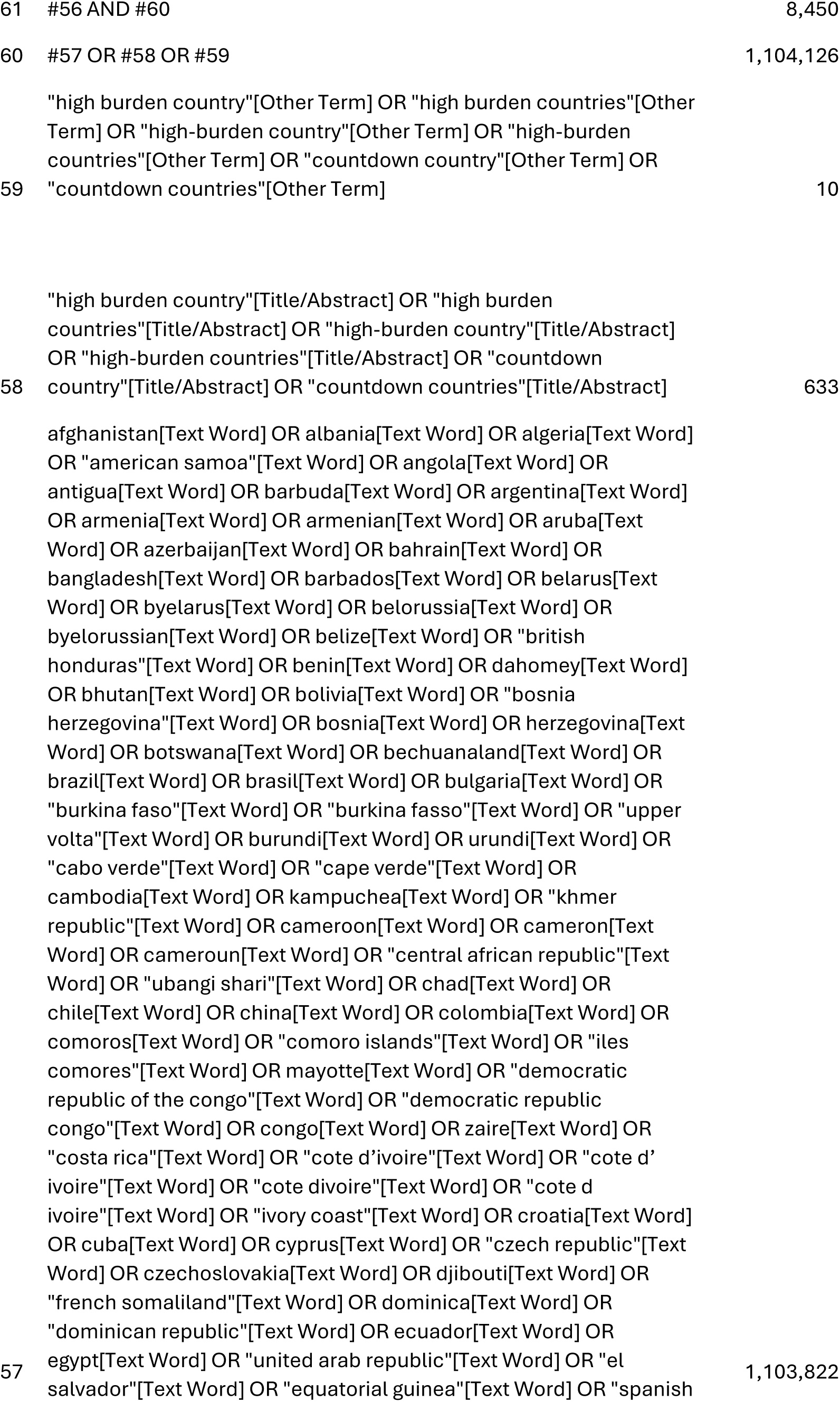

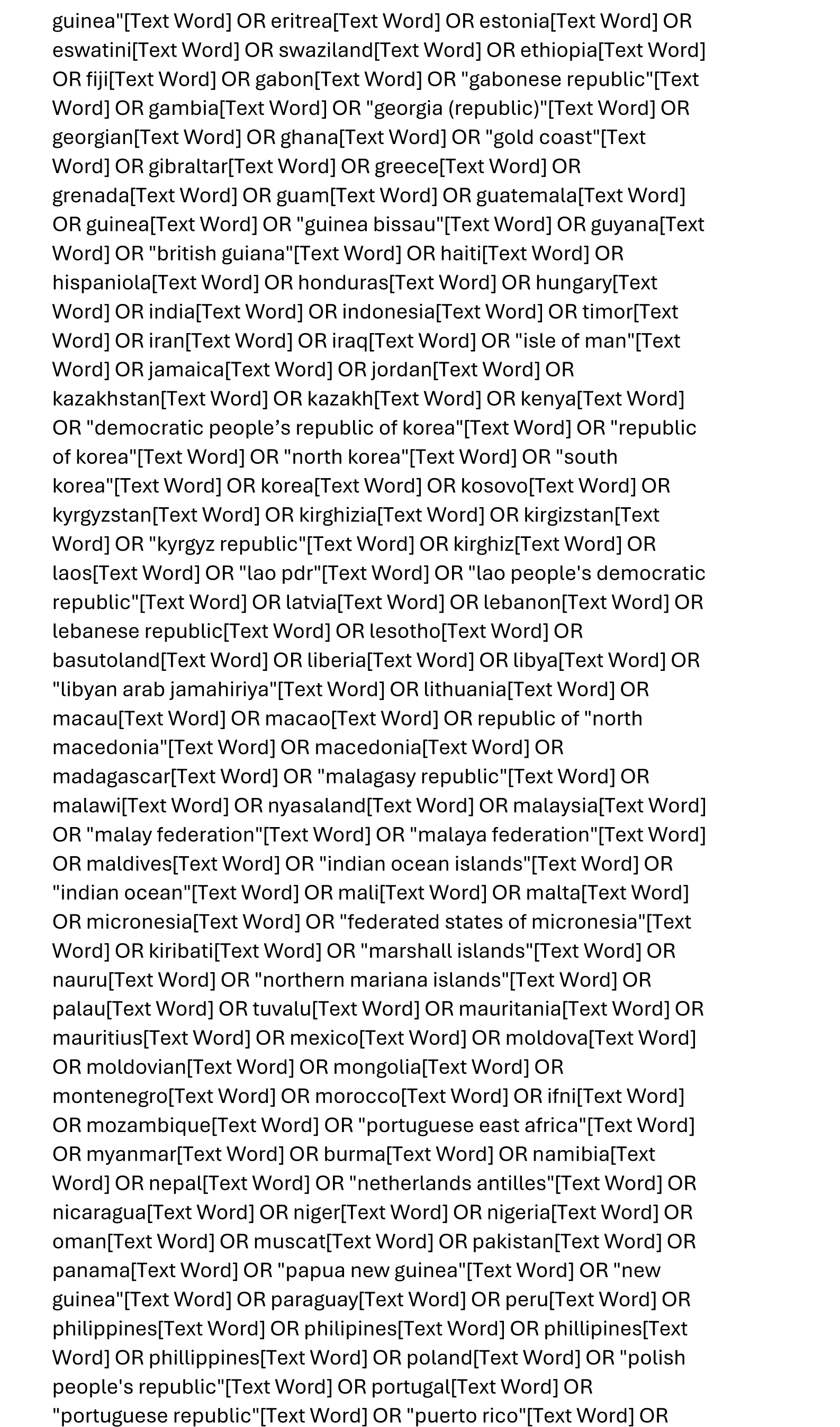

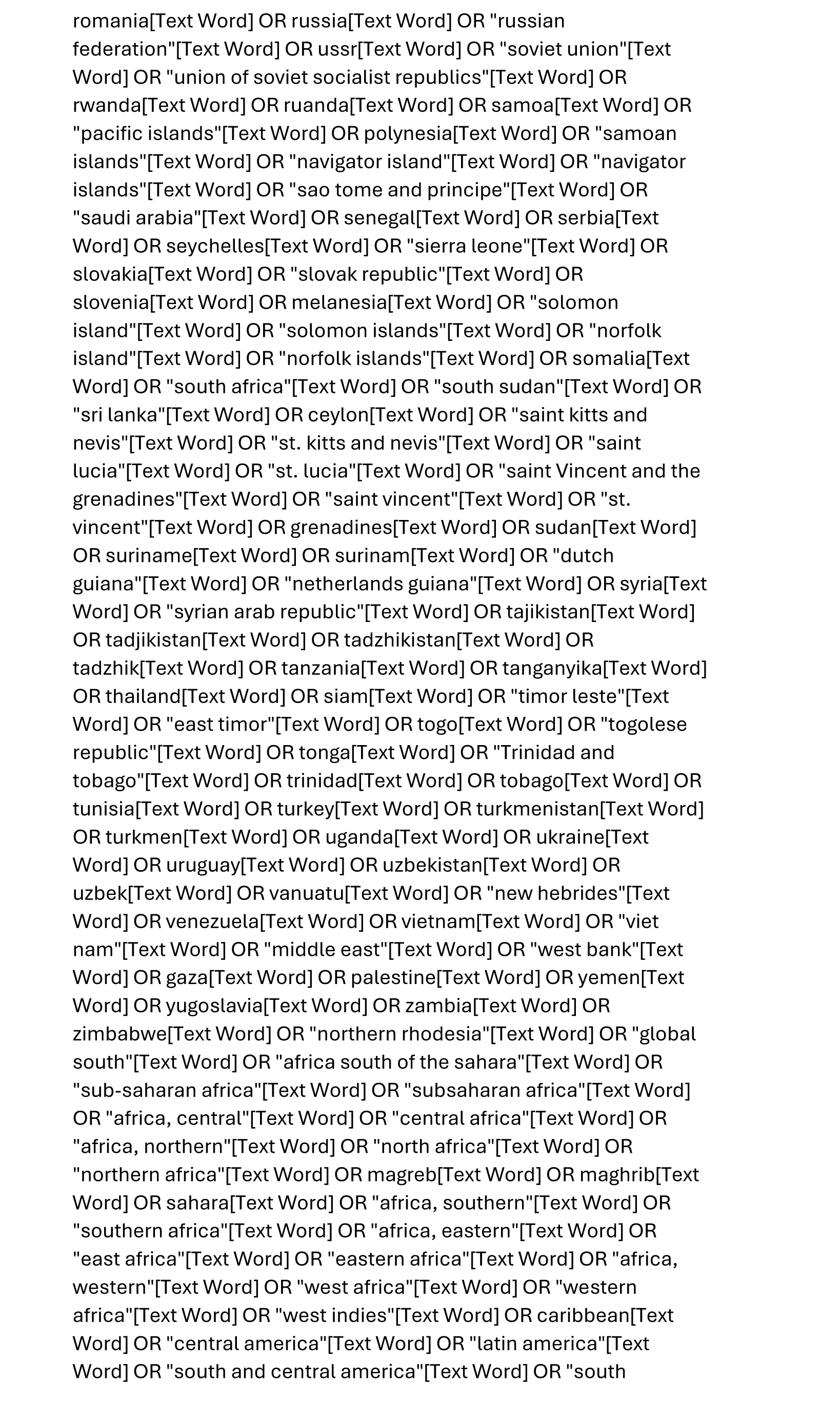

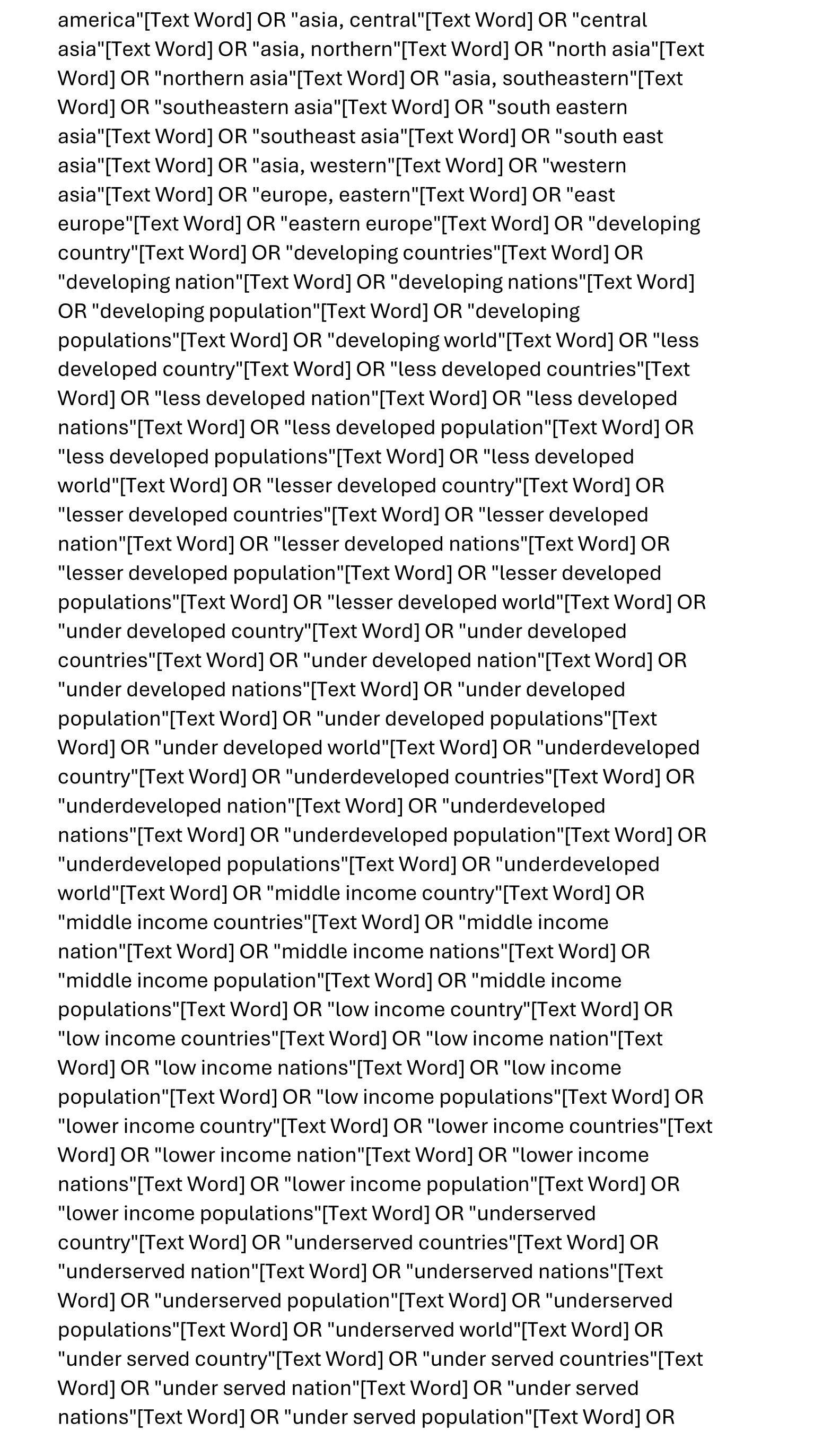

|  |  |  |
| --- | --- | --- |
|  | "under served populations"[Text Word] OR "under served world"[Text Word] OR "deprived country"[Text Word] OR "deprived countries"[Text Word] OR "deprived nation"[Text Word] OR "deprived nations"[Text Word] OR "deprived population"[Text Word] OR "deprived populations"[Text Word] OR "deprived world"[Text Word] OR "poor country"[Text Word] OR "poor countries"[Text Word] OR "poor nation"[Text Word] OR "poor nations"[Text Word] OR "poor population"[Text Word] OR "poor populations"[Text Word] OR "poor world"[Text Word] OR "poorer country"[Text Word] OR "poorer countries"[Text Word] OR "poorer nation"[Text Word] OR "poorer nations"[Text Word] OR "poorer population"[Text Word] OR "poorer populations"[Text Word] OR "poorer world"[Text Word] OR "developing economy"[Text Word] OR "developing economies"[Text Word] OR "less developed economy"[Text Word] OR "less developed economics"[Text Word] OR "lesser developed economy"[Text Word] OR "lesser developed economies"[Text Word] OR "under developed economy"[Text Word] OR "under developed economies"[Text Word] OR "underdeveloped economy"[Text Word] OR "underdeveloped economies"[Text Word] OR "middle income economy"[Text Word] OR "middle income economies"[Text Word] OR "low income economy"[Text Word] OR "low income economies"[Text Word] OR "lower income economy"[Text Word] OR "lower income economies"[Text Word] OR "low gdp"[Text Word] OR "low gnp"[Text Word] OR "low gross domestic"[Text Word] OR "low gross national"[Text Word] OR "lower gdp"[Text Word] OR "lower gnp"[Text Word] OR "lower gross domestic"[Text Word] OR "lower gross national"[Text Word] OR lmic[Text Word] OR lmics[Text Word] OR "third world"[Text Word] OR "lami country"[Text Word] OR "lami countries"[Text Word] OR "transitional country"[Text Word] OR "transitional economies"[Text Word] OR "emerging economy"[Text Word] OR "emerging economies"[Text Word] OR "emerging nation"[Text Word] OR "emerging nations"[Text Word] |  |
| 56 | #1 OR #55 | 39,866 |
| 55 | #21 AND #54 | 39,859 |
|  | #22 OR #23 OR #24 OR #25 OR #26 OR #27 OR #28 OR #29 OR #30 OR #31 OR #32 OR #33 OR #34 OR #35 OR #36 OR #37 OR #38 OR #39 OR #40 OR #41 OR #42 OR #43 OR #44 OR #45 OR #46 OR #47 |  |
| 54 | OR #48 OR #49 OR #50 OR #51 OR #52 OR #53 | 850,027 |
| 53 | "Product Surveillance, Postmarketing/statistics and numerical data"[MeSH Terms] | 2,361 |
| 52 | "Maternal Health Services/statistics and numerical data"[MeSH Terms] | 7,290 |
| 51 | "Maternal Exposure/statistics and numerical data"[MeSH Terms] | 1,123 |

|  |  |  |
| --- | --- | --- |
| 50 | pharmacovigilan*[ti] OR pharmaco-vigilan*[ti] OR<br>pharmacovigilan*[ot] OR pharmaco-vigilan*[ot] | 3,751 |
| 49 | "Product Surveillance, Postmarketing"[mesh:noexp] | 7,553 |
| 48 | "data system"[ti] OR "data systems"[ti] OR "information system"[ti]<br>OR "information systems"[ti] OR "data system"[ot] OR "data<br>systems"[ot] OR "information system"[ot] OR "information<br>systems"[ot] | 13,581 |
| 47 | "Health Information Systems"[mesh] | 1,546 |
| 46 | (decision*[ti] AND support*[ti] AND clinical*[ti]) OR (decision*[ot]<br>AND support*[ot] AND clinical*[ot]) | 4,467 |
| 45 | "Decision Support Systems, Clinical"[mesh] | 9,106 |
| 44 | "Databases, Factual"[mesh] | 163,862 |
| 43 | "Databases as Topic"[mesh:noexp] | 9,688 |
| 42 | surveillance*[ti] OR surveillance*[ot] | 61,304 |
| 41 | "Population Surveillance"[mesh] | 74,026 |
| 40 | survey*[ti] OR survey*[ot] | 189,840 |
| 39 | "Health Surveys"[mesh:noexp] | 66,266 |
| 38 | "Health Care Surveys"[MeSH Major Topic] | 10,167 |
| 37 | "Surveys and Questionnaires"[MeSH Major Topic:noexp] | 49,876 |
| 36 | registry[ot] OR registries[ot] OR eregistr*[ot] OR "e-registry"[ot] OR<br>"e-registries"[ot] | 8,509 |
| 35 | registry[ti] OR registries[ti] OR eregistr*[ti] OR "e-registry"[ti] OR "e-registries"[ti] | 34,311 |
| 34 | Registries[mesh:noexp] | 104,749 |
| 33 | (preliminary[ot] OR "pilot project"[ot] OR "pilot projects"[ot]) AND<br>data[ot] | 35 |
| 32 | (preliminary[ti] OR "pilot project"[ti] OR "pilot projects"[ti]) AND<br>data[ti] | 2,947 |
| 31 | "Preliminary Data"[mesh] | 724 |
| 30 | "focus group"[ti] OR "focus groups"[ti] OR "focus group"[ot] OR<br>"focus groups"[ot] | 6,048 |
| 29 | "Focus Groups"[mesh] | 34,384 |
| 28 | databas*[ot] OR "data base"[ot] OR "data bases"[ot] OR<br>databank*[ot] OR "data bank"[ot] OR "data banks"[ot] OR<br>dataset*[ot] OR "data set"[ot] OR "data sets"[ot] | 14,502 |

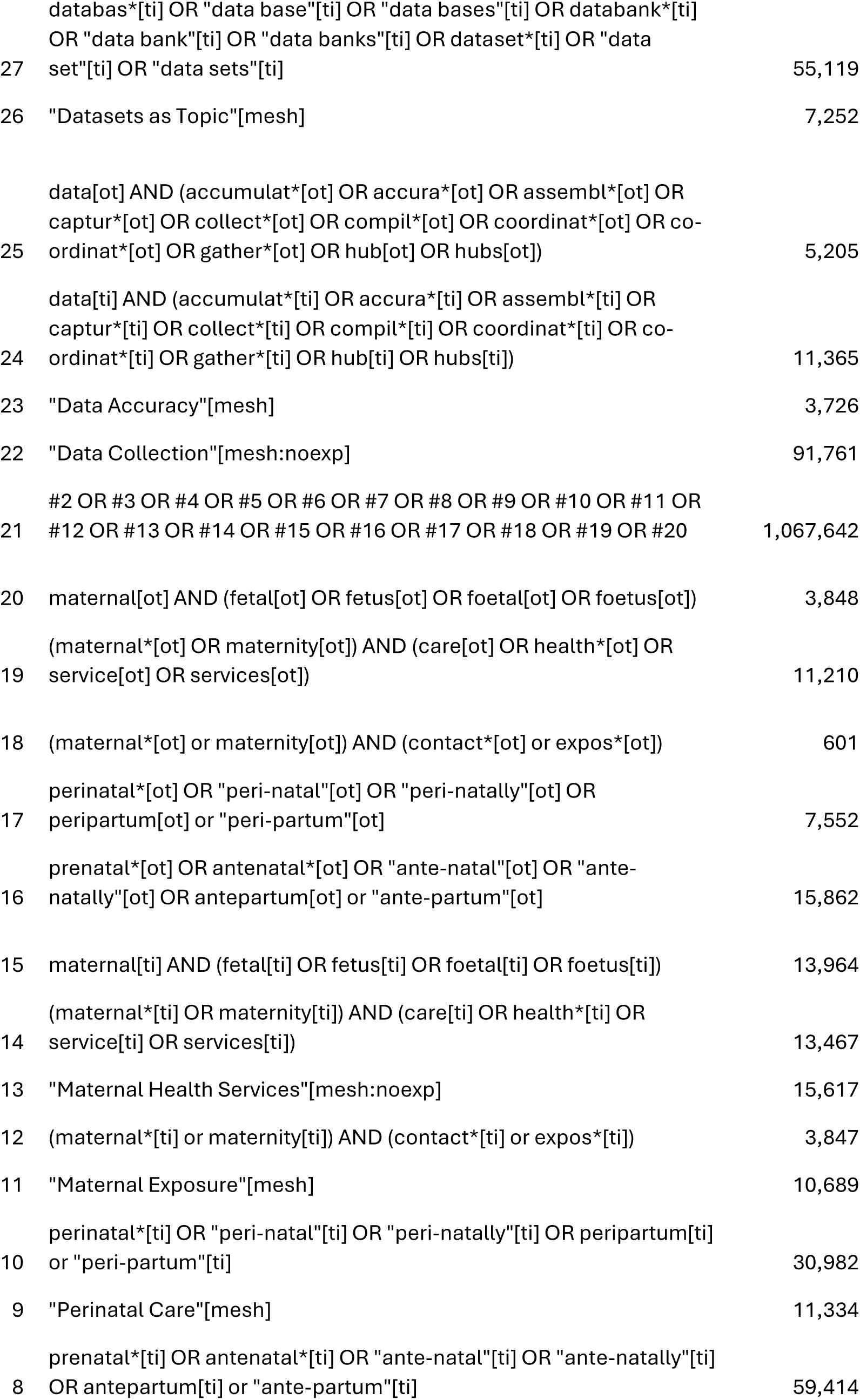

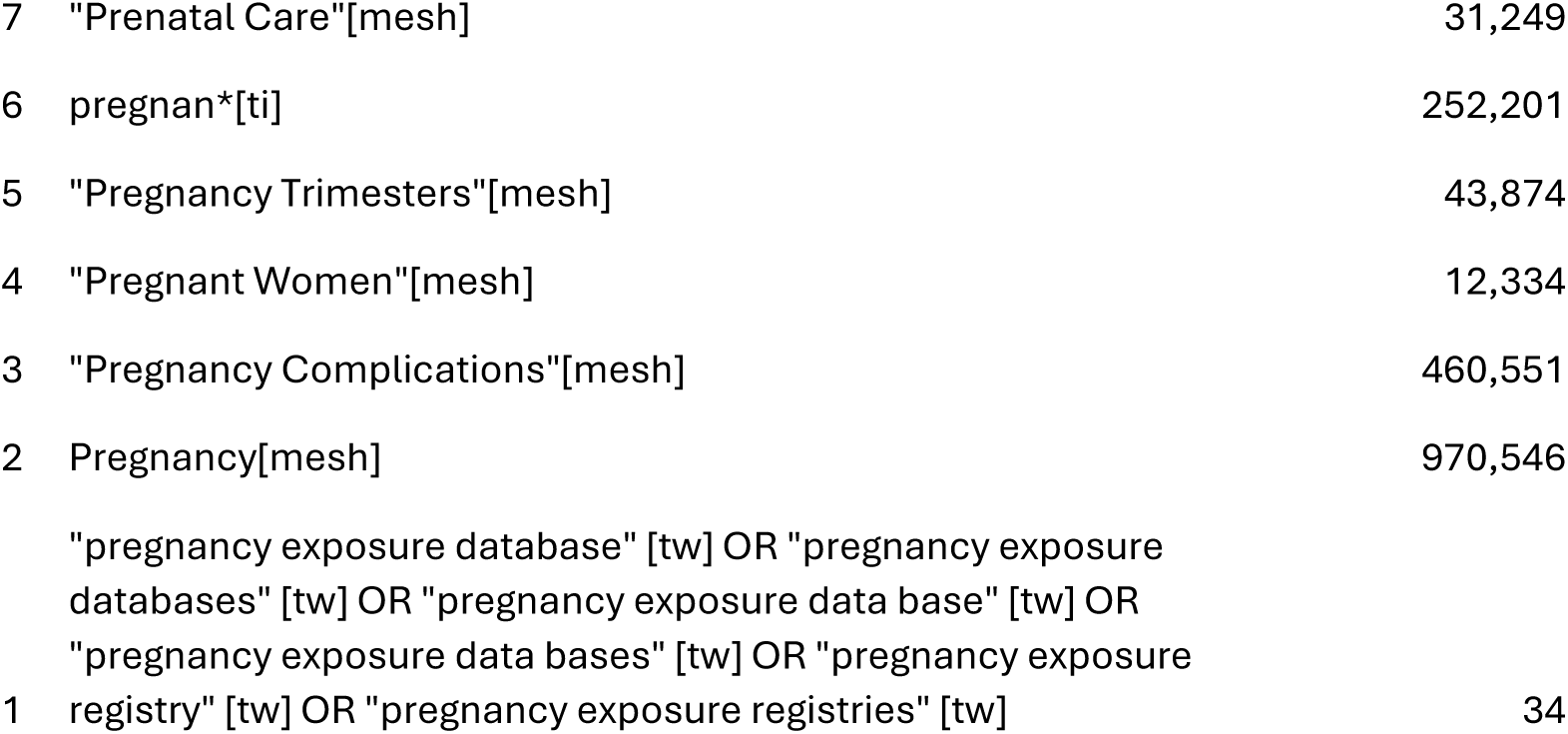

|  |  |  |
| --- | --- | --- |
| 7 | "Prenatal Care"[mesh] | 31,249 |
| 6 | pregnan*[ti] | 252,201 |
| 5 | "Pregnancy Trimesters"[mesh] | 43,874 |
| 4 | "Pregnant Women"[mesh] | 12,334 |
| 3 | "Pregnancy Complications"[mesh] | 460,551 |
| 2 | Pregnancy[mesh] | 970,546 |
| 1 | "pregnancy exposure database" [tw] OR "pregnancy exposure databases" [tw] OR "pregnancy exposure data base" [tw] OR "pregnancy exposure data bases" [tw] OR "pregnancy exposure registry" [tw] OR "pregnancy exposure registries" [tw] | 34 |

#### Embase

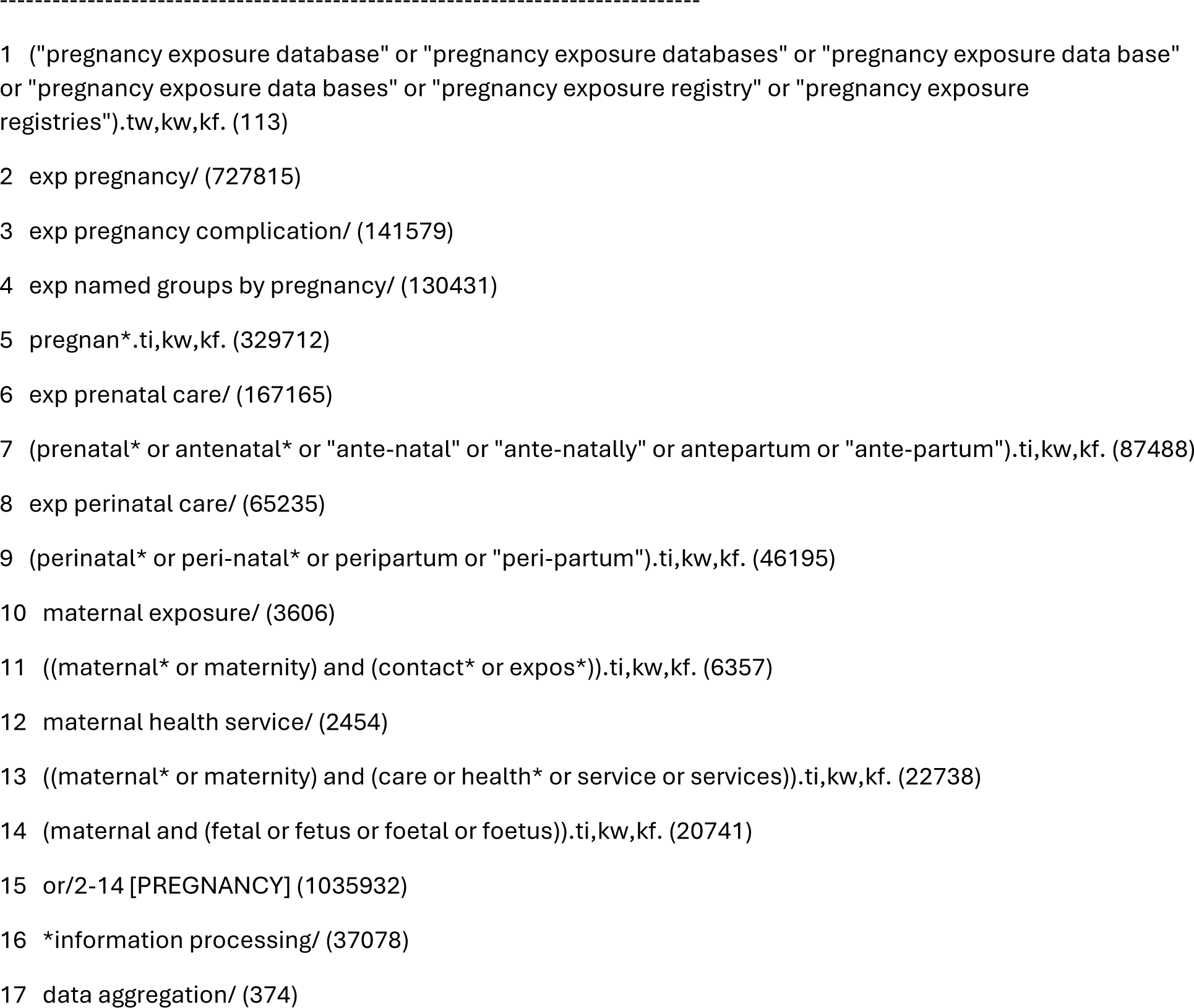

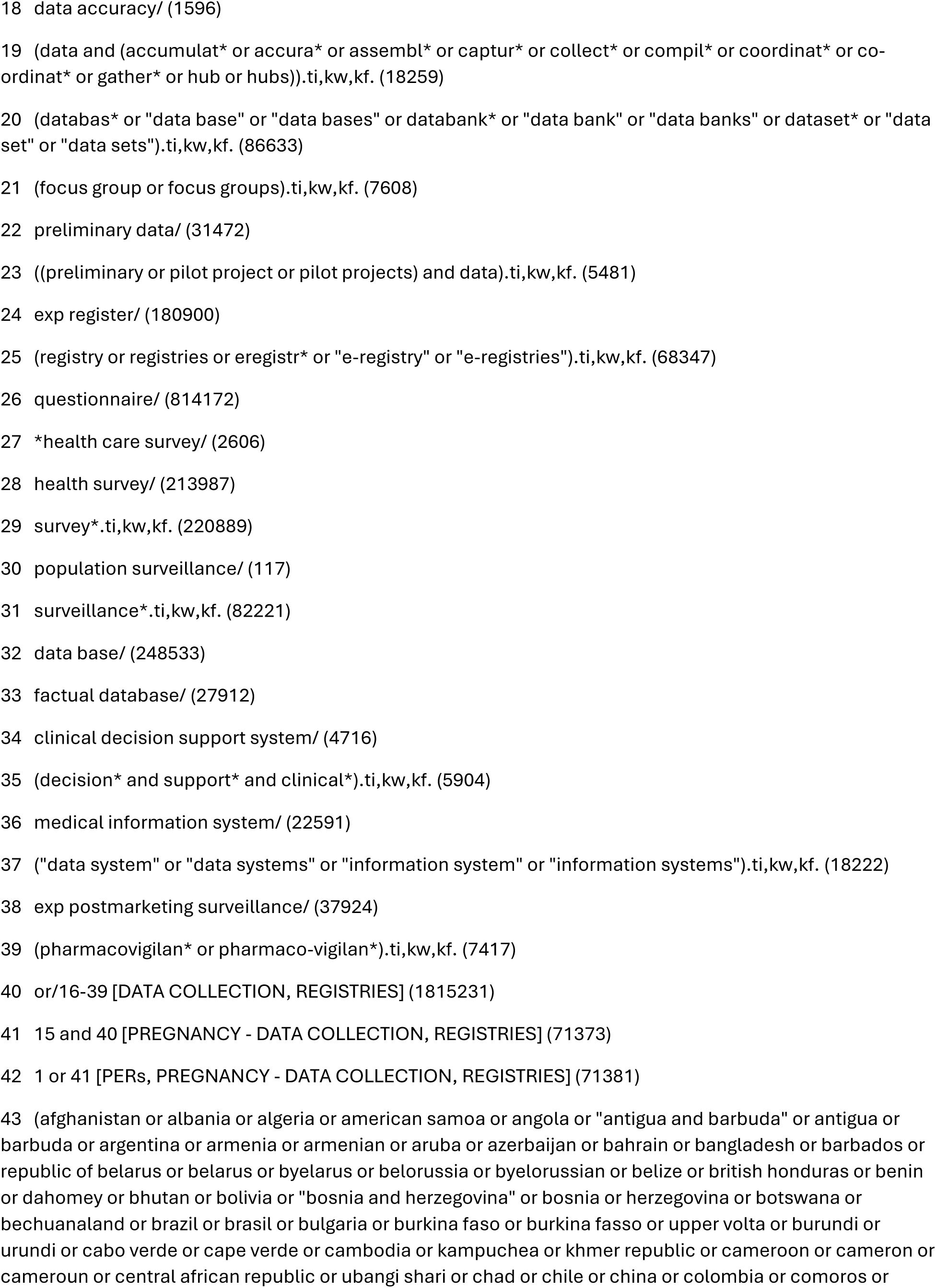

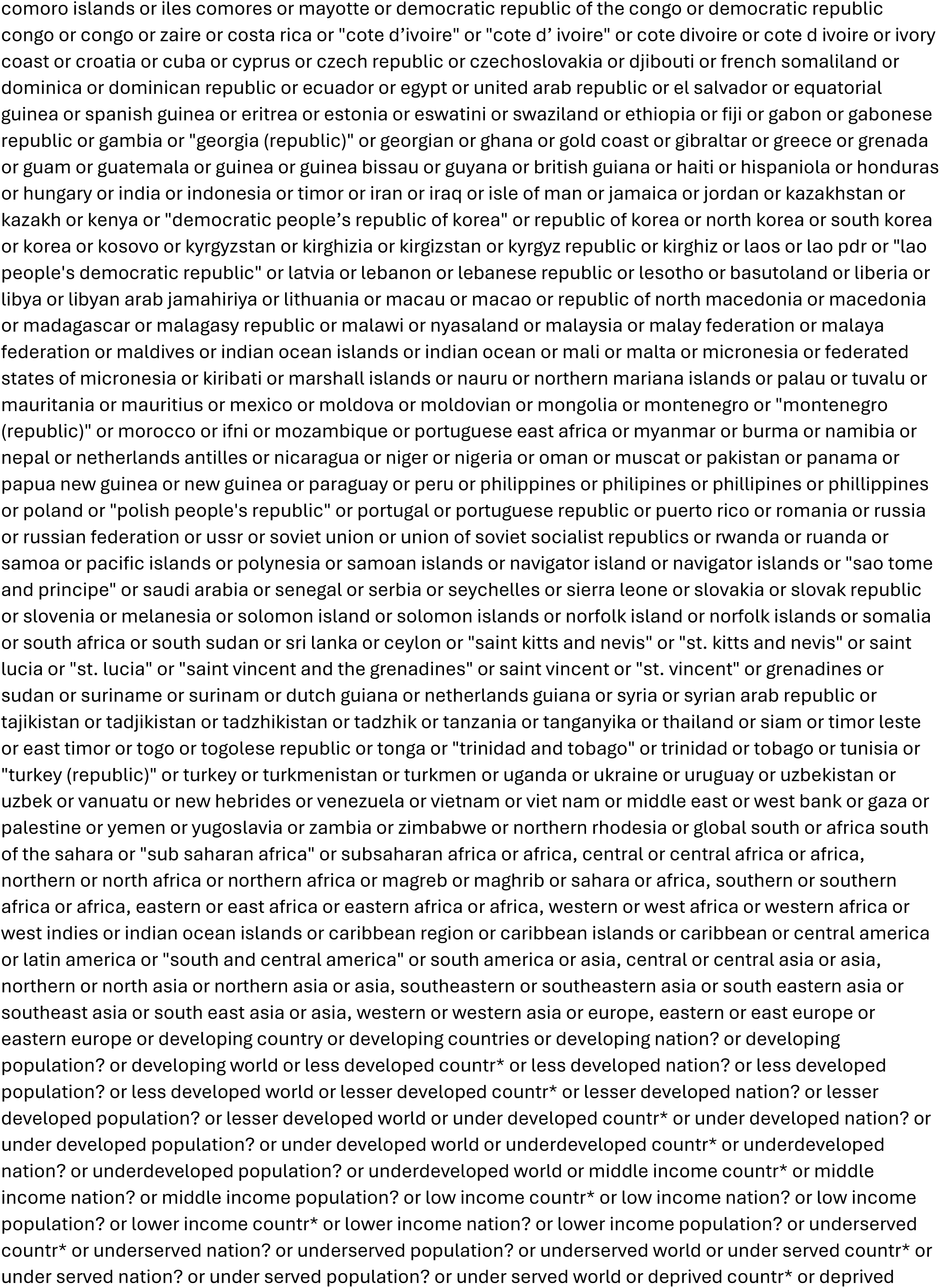

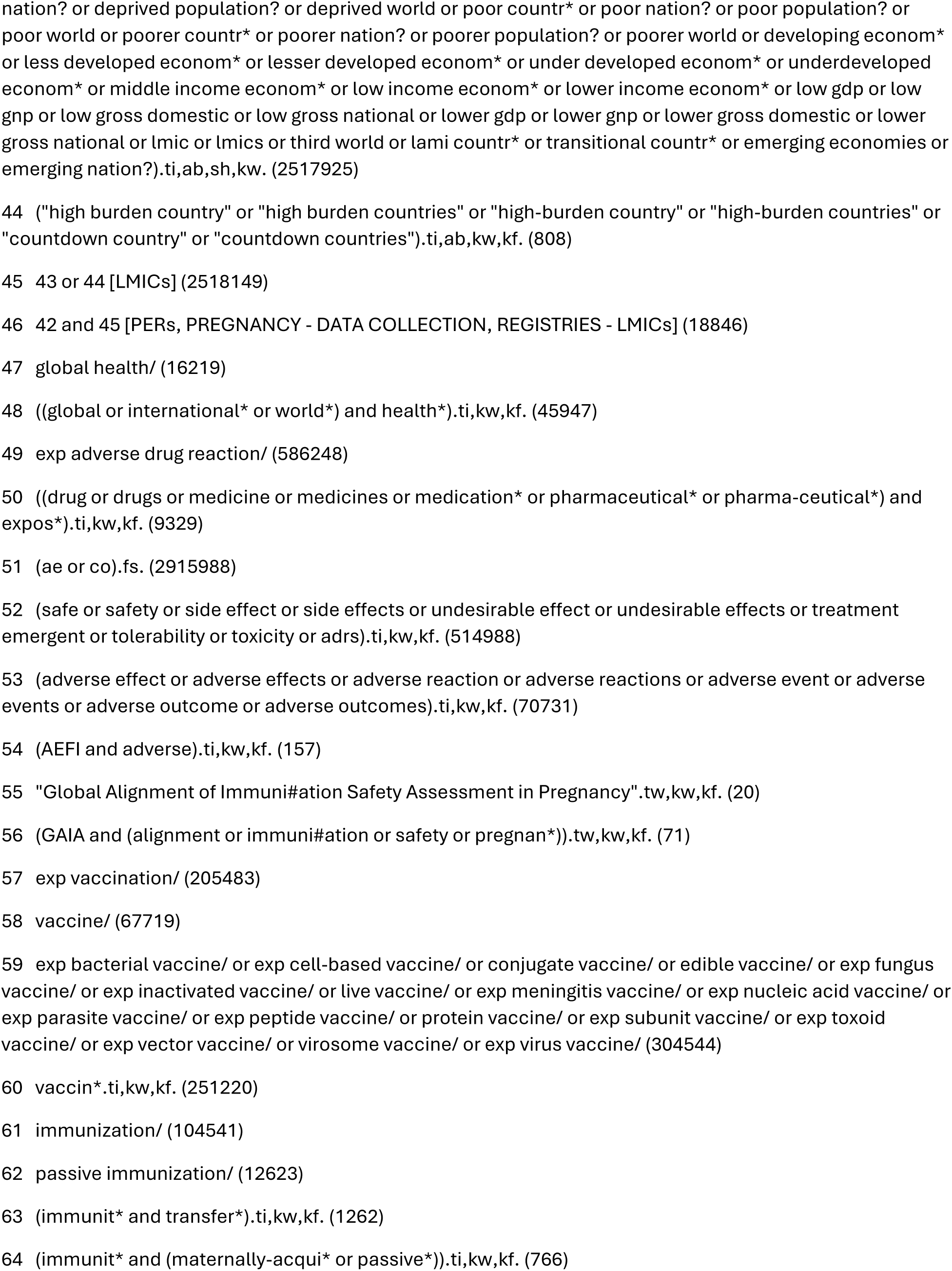

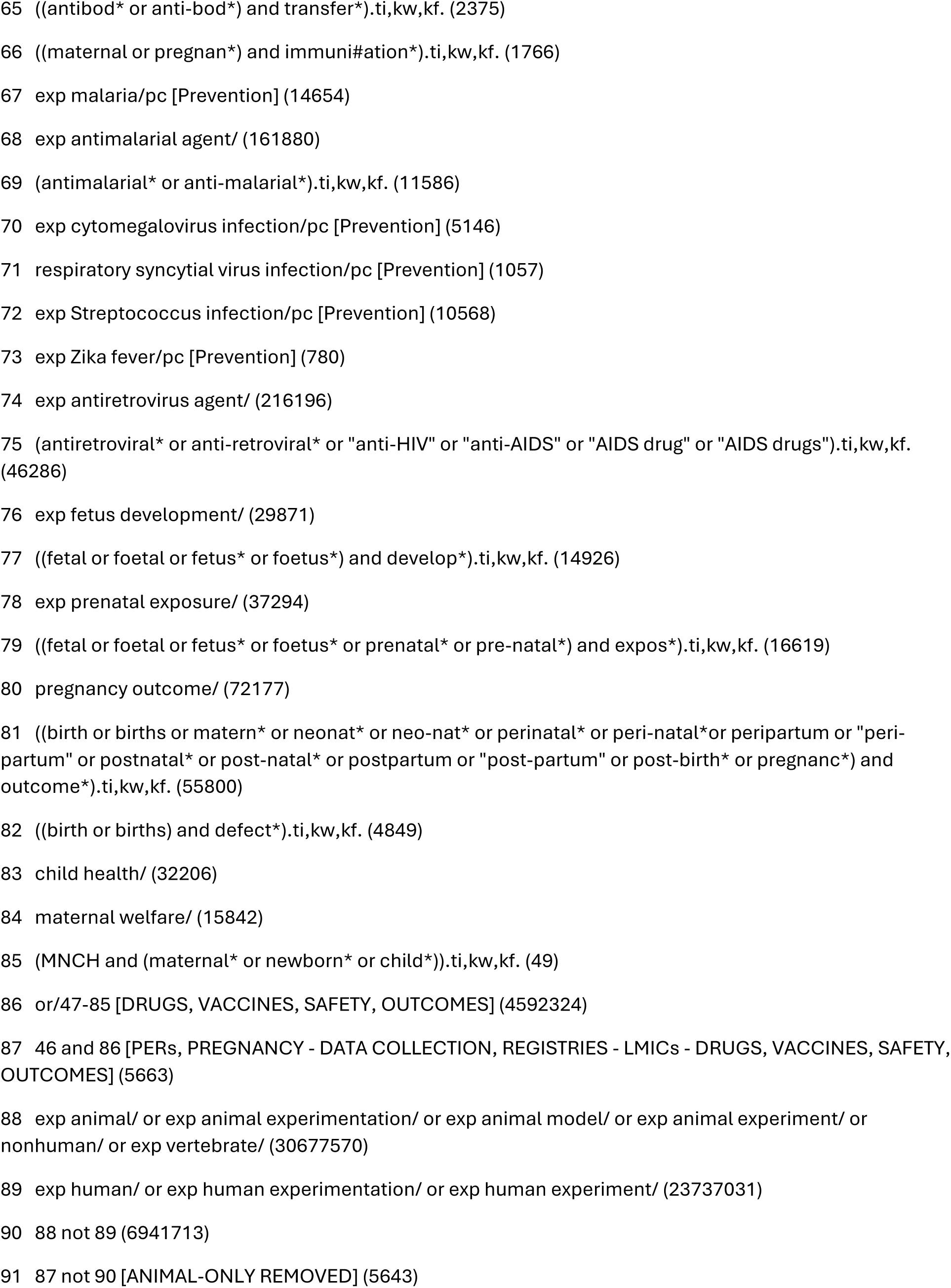

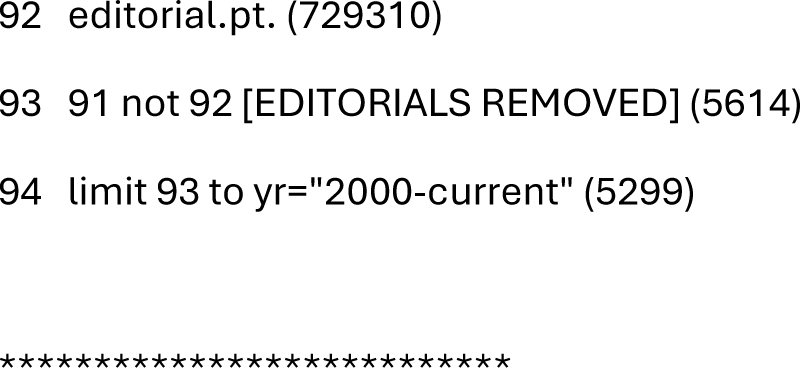

#### CINAHL

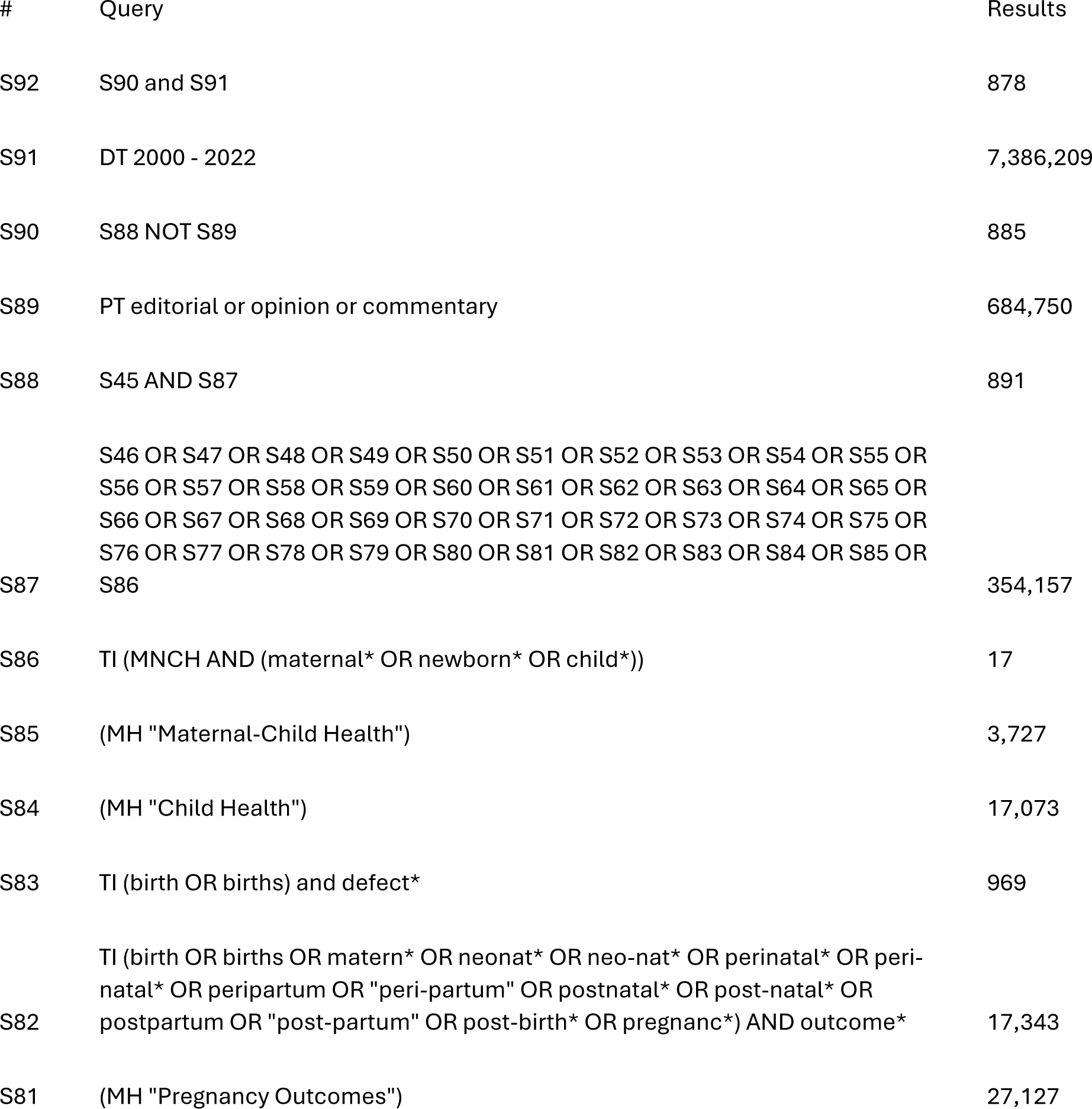

|  |  |  |
| --- | --- | --- |
| S80 | TI (fetal OR foetal OR fetus* OR foetus* or prenatal* or pre-natal*) AND expos* | 2,603 |
| S79 | (MH "Prenatal Exposure Delayed Effects") | 6,314 |
| S78 | TI (fetal OR foetal OR fetus* OR foetus*) AND develop* | 1,028 |
| S77 | (MH "Fetal Development") | 7,851 |
| S76 | TI antiretroviral* OR anti-retroviral* OR "anti-HIV" OR "anti-AIDS" OR "AIDS drug" OR "AIDS drugs" | 9,126 |
| S75 | (MH "Anti-Retroviral Agents+") | 24,911 |
| S74 | (MH "Zika Virus Infections/PC") | 168 |
| S73 | (MH "Streptococcal Infections+/PC") | 2,280 |
| S72 | (MH "Respiratory Syncytial Virus Infections/PC") | 583 |
| S71 | (MH "Hepatitis E/PC") | 69 |
| S70 | (MH "Cytomegalovirus Infections+/PC") | 509 |
| S69 | TI antimalarial* or (anti W0 malarial*) | 714 |
| S68 | (MH "Antimalarials+") | 8,320 |
| S67 | (MH "Malaria/PC") | 3,293 |
| S66 | TI (maternal* or pregnan*) and immuni?ation* | 253 |
| S65 | TI ((antibod* or (anti W0 bod*)) and transfer*) | 79 |
| S64 | TI immunit* AND (passive* or "maternally-acquired") | 13 |
| S63 | TI immunit* AND transfer* | 16 |
| S62 | (MH "Immunity, Maternally Acquired") | 232 |
| S61 | (MH "Immunization Programs") | 6,426 |
| S60 | (MH "Viral Vaccines+") | 36,206 |

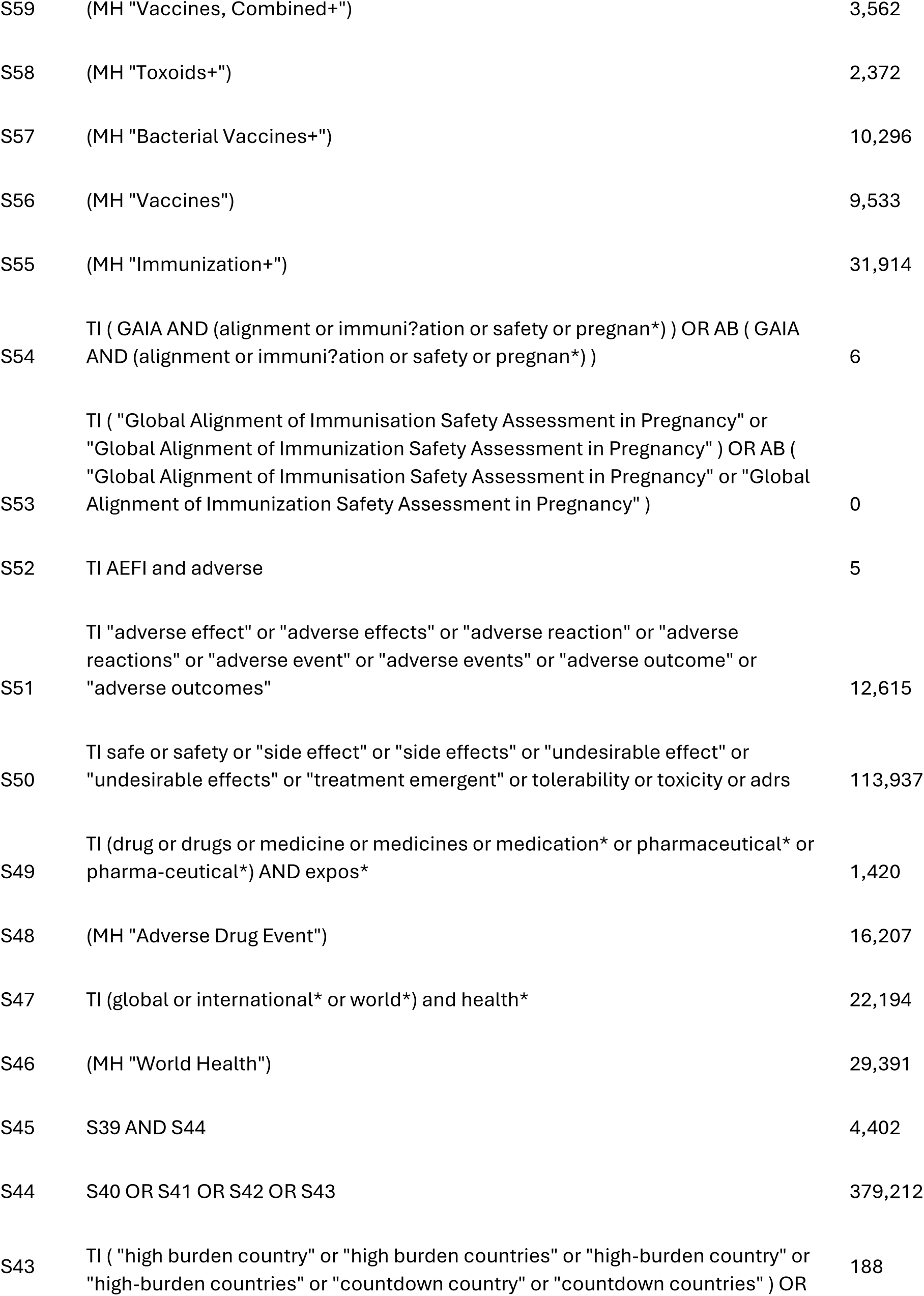

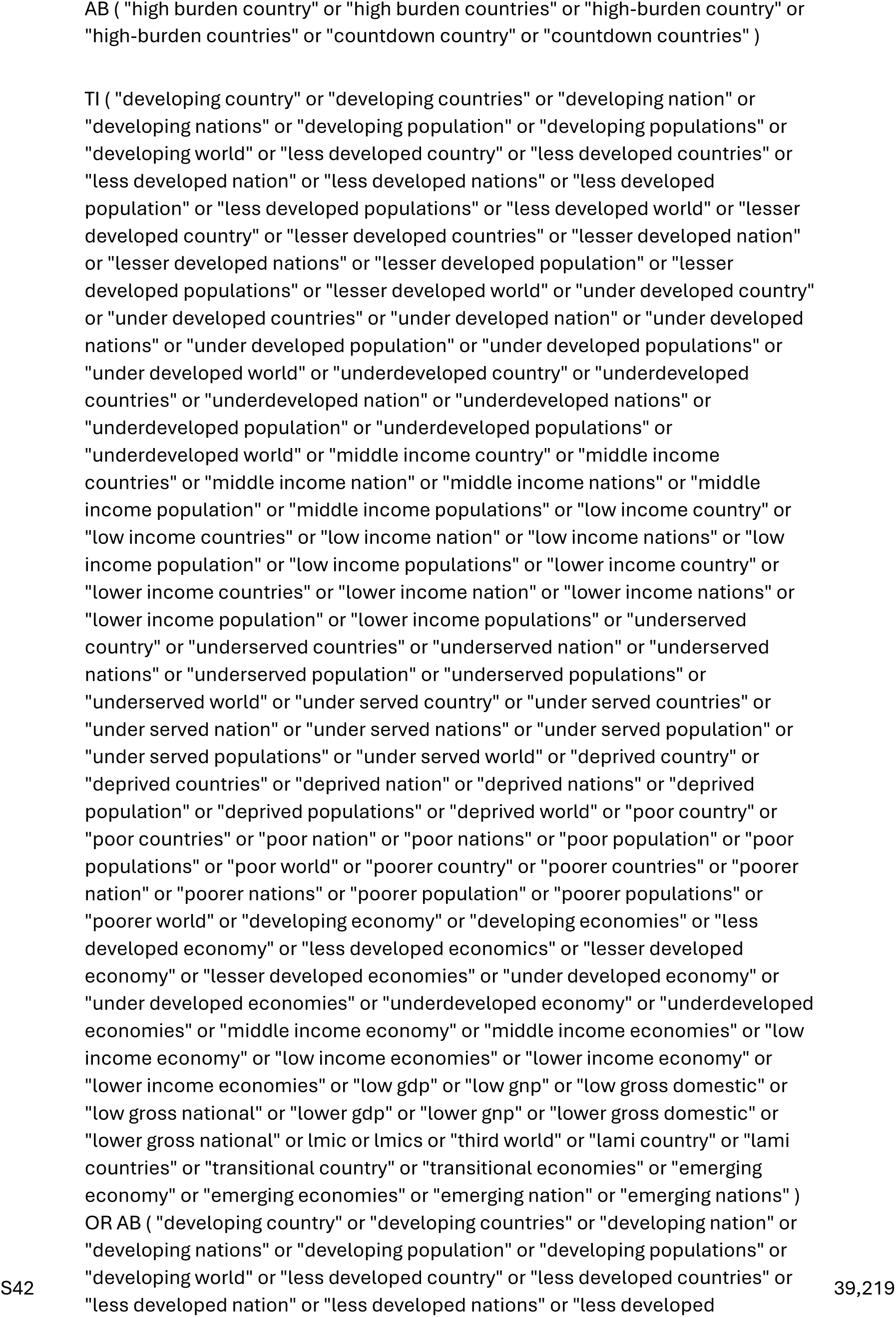

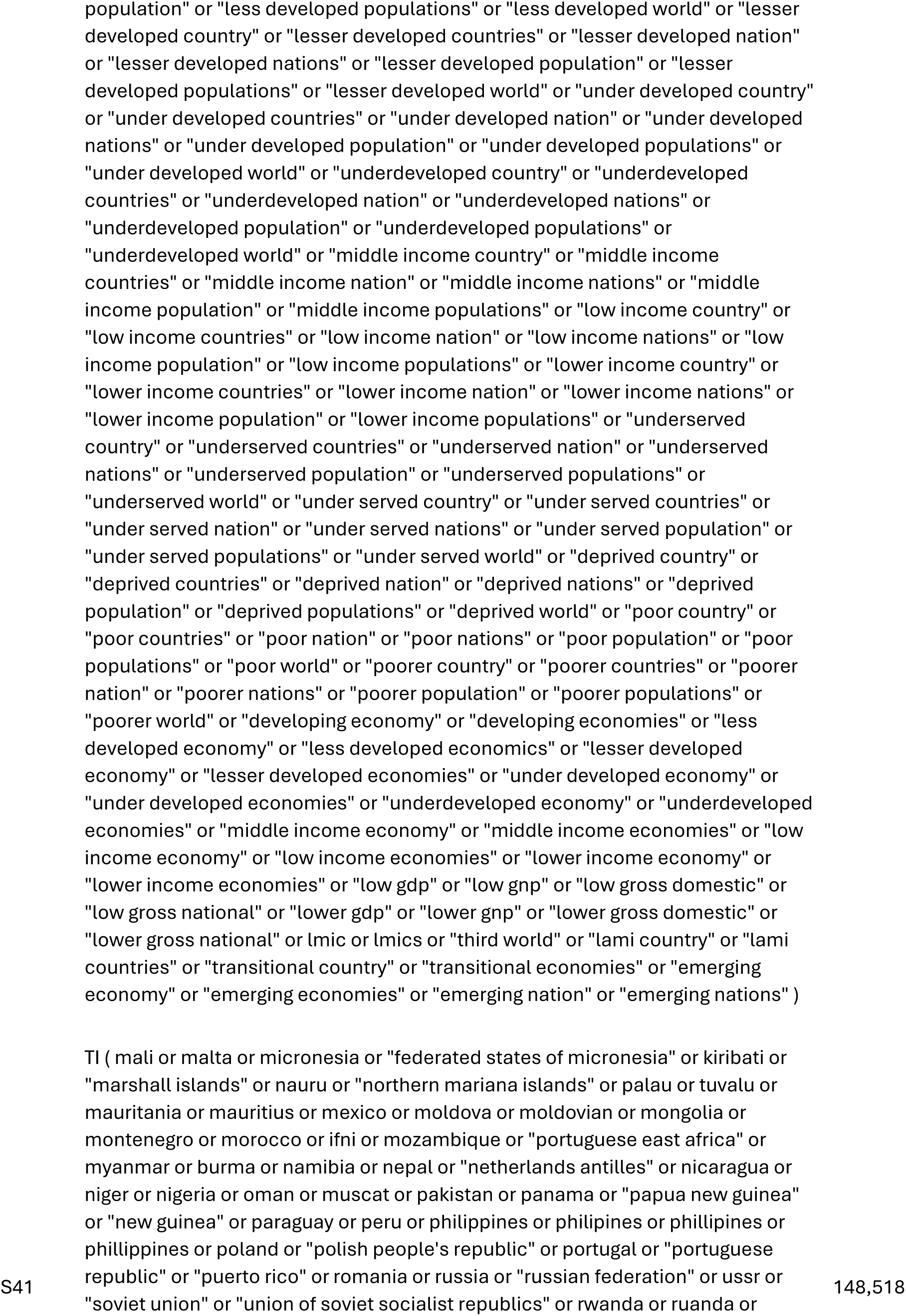

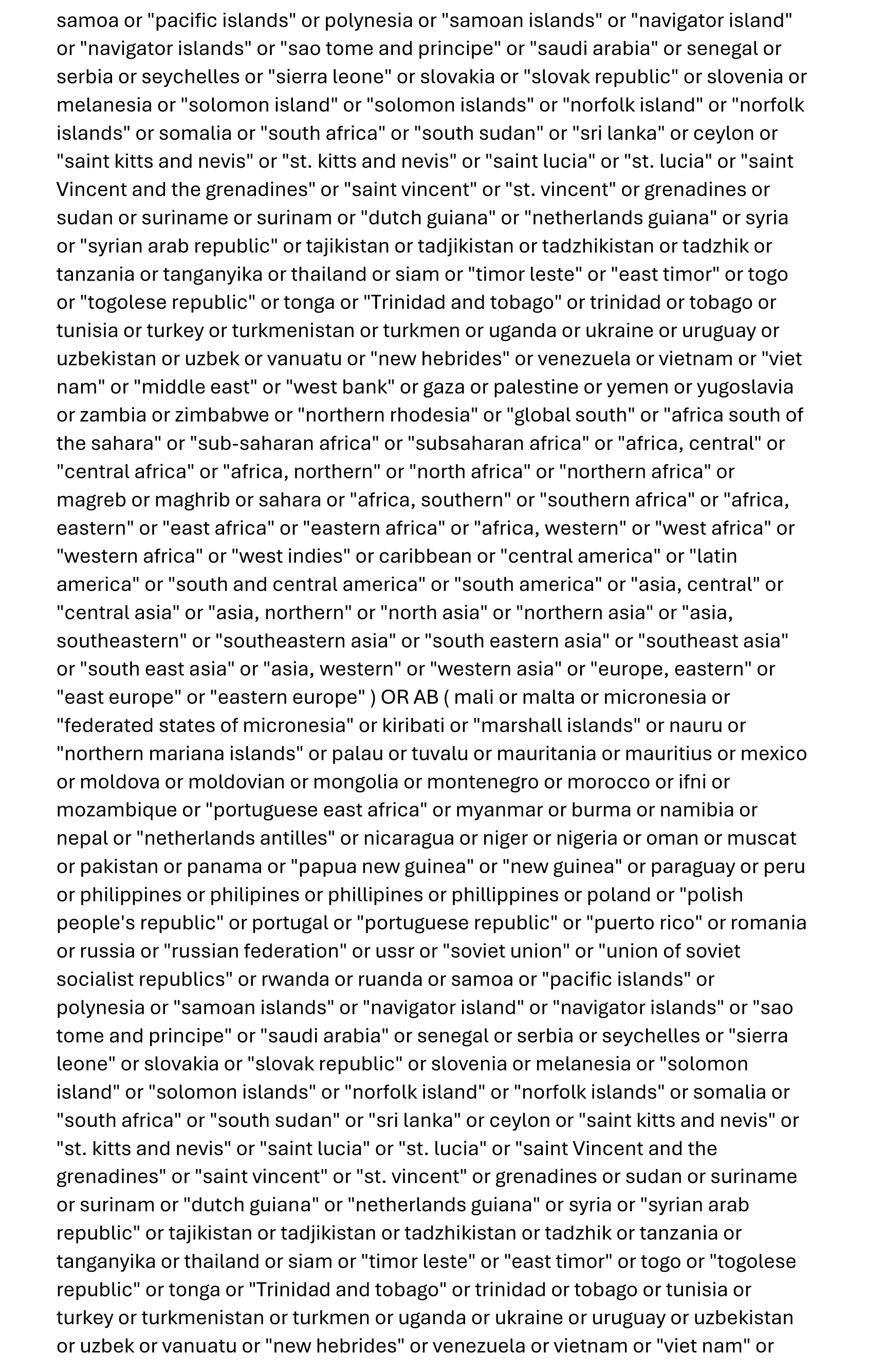

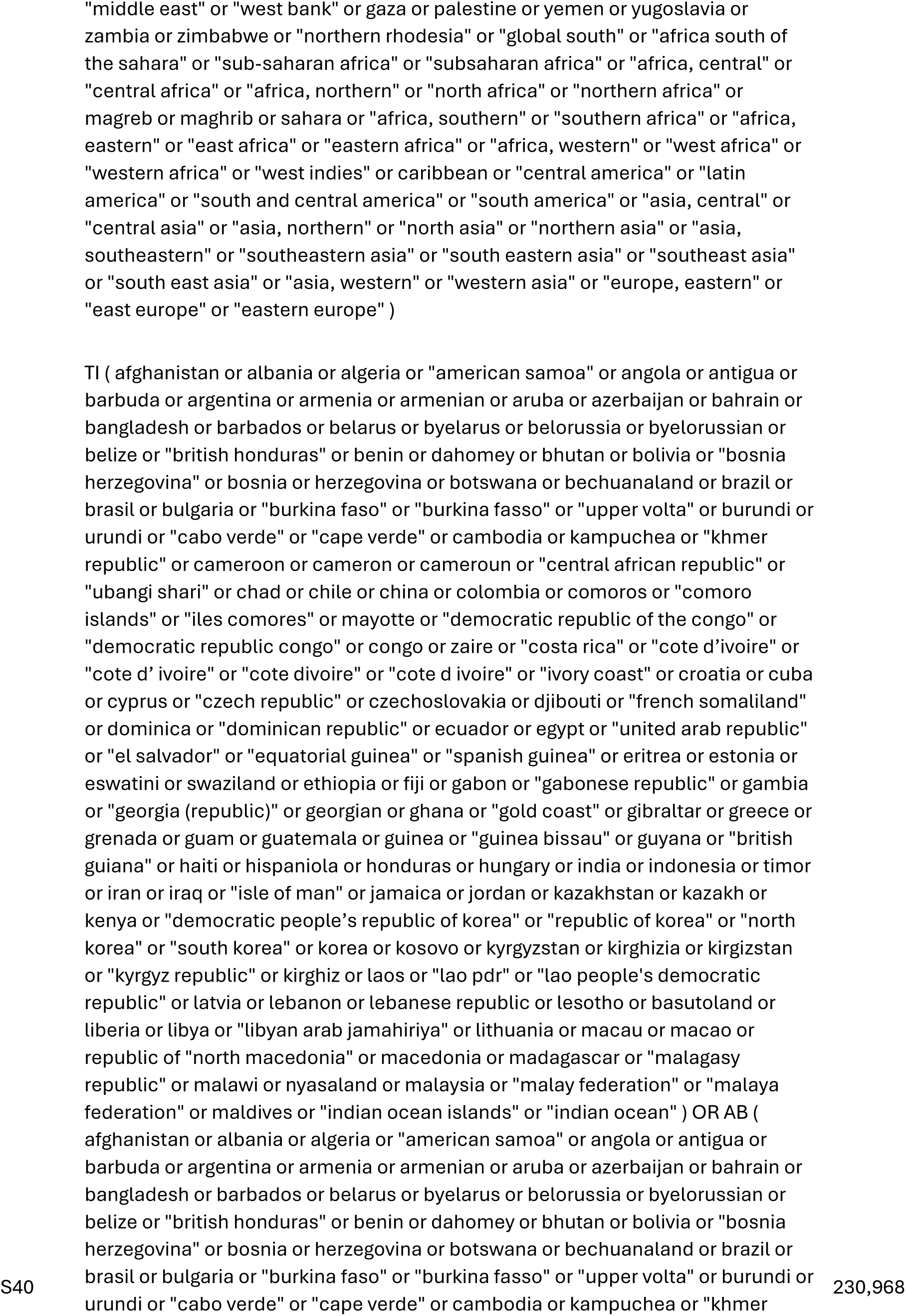

|  |  |  |
| --- | --- | --- |
|  | republic" or cameroon or cameron or cameroun or "central african republic" or "ubangi shari" or chad or chile or china or colombia or comoros or "comoro islands" or "iles comores" or mayotte or "democratic republic of the congo" or "democratic republic congo" or congo or zaire or "costa rica" or "cote d'ivoire" or "cote d' ivoire" or "cote divoire" or "cote d ivoire" or "ivory coast" or croatia or cuba or cyprus or "czech republic" or czechoslovakia or djibouti or "french somaliland" or dominica or "dominican republic" or ecuador or egypt or "united arab republic" or "el salvador" or "equatorial guinea" or "spanish guinea" or eritrea or estonia or eswatini or swaziland or ethiopia or fiji or gabon or "gabonese republic" or gambia or "georgia (republic)" or georgian or ghana or "gold coast" or gibraltar or greece or grenada or guam or guatemala or guinea or "guinea bissau" or guyana or "british guiana" or haiti or hispaniola or honduras or hungary or india or indonesia or timor or iran or iraq or "isle of man" or jamaica or jordan or kazakhstan or kazakh or kenya or "democratic people's republic of korea" or "republic of korea" or "north korea" or "south korea" or korea or kosovo or kyrgyzstan or kirghizia or kirgizstan or "kyrgyz republic" or kirghiz or laos or "lao pdr" or "lao people's democratic republic" or latvia or lebanon or lebanese republic or lesotho or basutoland or liberia or libya or "libyan arab jamahiriya" or lithuania or macau or macao or republic of "north macedonia" or macedonia or madagascar or "malagasy republic" or malawi or nyasaland or malaysia or "malay federation" or "malaya federation" or maldives or "indian ocean islands" or "indian ocean" ) |  |
| S39 | S1 OR S38 | 15,244 |
| S38 | S16 AND S37 | 15,243 |
| S37 | S17 OR S18 OR S19 OR S20 OR S21 OR S22 OR S23 OR S24 OR S25 OR S26 OR S27 OR S28 OR S29 OR S30 OR S31 OR S32 OR S33 OR S34 OR S35 OR S36 | 354,620 |
| S36 | (MH "Product Surveillance+/SN") | 1 |
| S35 | (MH "Maternal Health Services+/SN") | 1,245 |
| S34 | TI pharmacovigilan* or (pharmaco W0 vigilan*) | 846 |
| S33 | (MH "Product Surveillance+") | 2,417 |
| S32 | TI "data system" or "data systems" or "information system" or "information systems" | 4,466 |
| S31 | (MH "Health Information Systems") | 3,566 |
| S30 | TI (decision* AND support* AND clinical*) | 1,865 |

|  |  |  |
| --- | --- | --- |
| S29 | (MH "Decision Support Systems, Clinical") | 6,139 |
| S28 | TI surveillance* | 15,975 |
| S27 | (MH "Population Surveillance+") | 11,277 |
| S26 | TI survey* | 71,609 |
| S25 | (MH "Surveys") | 157,567 |
| S24 | TI registry or registries or eregistr* or "e-registry" or "e-registries" | 13,946 |
| S23 | TI (preliminary or "pilot project" or "pilot projects") AND data | 682 |
| S22 | TI "focus group" or "focus groups" | 2,957 |
| S21 | (MH "Focus Groups") | 48,414 |
| S20 | TI databas* or "data base" or "data bases" or databank* or "data bank" or "data banks" or dataset* or "data set" or "data sets" | 16,019 |
| S19 | TI data AND (accumulat* or accura* or assembl* or captur* or collect* or compil* or coordinat* or co-ordinat* or gather* or hub or hubs) | 4,425 |
| S18 | (MH "Data Curation") | 184 |
| S17 | (MH "Data Collection") | 49,942 |
| S16 | S2 OR S3 OR S4 OR S5 OR S6 OR S7 OR S8 OR S9 OR S10 OR S11 OR S12 OR S13 OR S14 OR S15 | 290,175 |
| S15 | TI maternal AND (fetal or fetus or foetal or foetus) | 4,004 |
| S14 | TI (maternal* or maternity) AND (care or health* or service or services) | 9,718 |
| S13 | (MH "Maternal Health Services") | 11,199 |
| S12 | TI (maternal* or maternity) AND (contact* or expos*) | 1,028 |
| S11 | (MH "Maternal Exposure") | 2,588 |
| S10 | TI perinatal* or "peri-natal" or "peri-natally" or peripartum or "peri-partum" | 12,858 |

|  |  |  |
| --- | --- | --- |
| S9 | (MH "Perinatal Care") | 5,019 |
| S8 | TI prenatal* or antenatal* or "ante-natal" or "ante-natally" or antepartum or "ante-partum" | 20,877 |
| S7 | (MH "Prenatal Care") | 19,422 |
| S6 | TI pregnan* | 80,123 |
| S5 | (MH "Pregnancy Trimesters+") | 13,521 |
| S4 | (MH "Expectant Mothers") | 11,382 |
| S3 | (MH "Pregnancy Complications+") | 106,825 |
| S2 | (MH "Pregnancy") OR (MH "Pregnancy, Multiple+") OR (MH "Pregnancy, Unplanned") OR (MH "Pregnancy, Unwanted") OR (MH "Pregnancy, High Risk") OR (MH "Pregnancy, Prolonged") | 227,240 |
| S1 | TI ( "pregnancy exposure database" OR "pregnancy exposure databases" OR "pregnancy exposure data base" OR "pregnancy exposure data bases" OR "pregnancy exposure registry" OR "pregnancy exposure registries" ) OR AB ( "pregnancy exposure database" OR "pregnancy exposure databases" OR "pregnancy exposure data base" OR "pregnancy exposure data bases" OR "pregnancy exposure registry" OR "pregnancy exposure registries" ) | 7 |

#### Global Index Medicus

tw:((tw:(pregnan* exposure* database*)) OR (tw:(pregnan* exposure* "data base")) OR (tw:(pregnan* exposure* "data bases")) OR (tw:(pregnan* exposure* databank*)) OR (tw:(pregnan* exposure* "data bank")) OR (tw:(pregnan* exposure* "data banks")) OR (tw:(pregnan* exposure* register)) OR (tw:(pregnan* exposure* registries)) OR (tw:(pregnan* exposure* registry)) OR (tw:(pregnan* exposure* registries))) – *108 records*

tw:((ti:((pregnan* OR prenatal* OR antenatal* OR "ante-natal" OR "ante-natally" OR antepartum OR "ante-partum" OR perinatal* OR "peri-natal" OR "peri-natally" OR peripartum OR "peri-partum" OR maternal*) AND (data OR database* OR databank* OR register OR registers OR registry OR registries OR survey* OR surveillance OR pharmacovigilan* OR "pharmaco-vigilance") AND (safe OR safety OR "side effect" of "side effects" OR "undesirable effect" OR "undesirable effects" OR "treatment emergent" OR tolerability OR toxicity OR adrs OR aefi)))) – *361 records*

tw:(ti:((pregnan* OR prenatal* OR antenatal* OR "ante-natal" OR "ante-natally" OR antepartum OR "ante-partum" OR perinatal* OR "peri-natal" OR "peri-natally" OR peripartum OR "peri-partum" OR maternal*) AND (data OR database* OR databank* OR register OR registers OR registry OR registries OR survey* OR surveillance OR pharmacovigilan* OR "pharmaco-vigilance") AND (MNCH or "maternal health" or "child health" or "infant health" or "birth defect" or "birth defects" or "birth outcome" or "birth outcomes" or "pregnancy outcome" or "pregnancy outcomes" or "neonatal outcome" or "neonatal outcomes"))) – *50 records*

tw:(ti:((pregnan* OR prenatal* OR antenatal* OR "ante-natal" OR "ante-natally" OR antepartum OR "ante-partum" OR perinatal* OR "peri-natal" OR "peri-natally" OR peripartum OR "peri-partum" OR maternal*) AND (data OR database* OR databank* OR register OR registers OR registry OR registries OR survey* OR surveillance OR pharmacovigilan* OR "pharmaco-vigilance") AND (vaccine or vaccines or vaccination* or immunization* or immunization* or “maternally-acquired”))) – *0 records*

tw:((ti:((pregnan* OR prenatal* OR antenatal* OR "ante-natal" OR "ante-natally" OR antepartum OR "ante-partum" OR perinatal* OR "peri-natal" OR "peri-natally" OR peripartum OR "peri-partum" OR maternal*) AND (data OR database* OR databank* OR register OR registers OR registry OR registries OR survey* OR surveillance OR pharmacovigilan* OR "pharmaco-vigilance") AND (drug OR drugs OR medicine OR medicines OR medication* OR pharmaceutical* OR "pharma-ceutical" OR "pharma-ceuticals" OR vaccin* OR immun* OR antimalarial* OR "anti-malarial" OR "anti-malarials" OR antiviral* OR "anti-viral" OR "anti-virals" OR antiretroviral* OR "anti-retroviral" OR "anti-retrovirals" OR "Anti-HIV"

OR "Anti-AIDS")))) – *10 records*

tw:((ti:((pregnan* OR prenatal* OR antenatal* OR "ante-natal" OR "ante-natally" OR antepartum OR "ante-partum" OR perinatal* OR "peri-natal" OR "peri-natally" OR peripartum OR "peri-partum" OR maternal*) AND (data OR database* OR databank* OR register OR registers OR registry OR registries OR survey* OR surveillance OR pharmacovigilan* OR "pharmaco-vigilance") AND (fetal* OR foetal* OR fetus* OR foetus* OR neonat* OR newborn* OR infant OR infants OR infanc* OR child*)))) – *125 records*

tw:((ti:((pregnan* OR prenatal* OR antenatal* OR "ante-natal" OR "ante-natally" OR antepartum OR "ante-partum" OR perinatal* OR "peri-natal" OR "peri-natally" OR peripartum OR "peri-partum" OR maternal*) AND (data OR database* OR databank* OR register OR registers OR registry OR registries OR survey* OR surveillance OR pharmacovigilan* OR "pharmaco-vigilance"))) AND (tw:(drug OR drugs OR medicine OR medicines OR medication* OR pharmaceutical* OR "pharma-ceutical" OR "pharma-ceuticals" OR vaccin* OR immunisation* OR immunization* OR antimalarial* OR "anti-malarial" OR "anti-malarials" OR antiviral* OR "anti-viral" OR "anti-virals" OR antiretroviral* OR "anti-retroviral" OR "anti-retrovirals" OR "Anti-HIV" OR "Anti-AIDS"))) – *71 records*

TOTAL: 725 records

#### Google Scholar

"pregnant|pregnancy|prenatal"+"registry|registries|surveillance|pharmacovigilance" + "expose|exposed|exposes|exposure|exposures"+[names of LMICs]

### Data extraction form

#### PART 1: STUDY IDENTIFICATION / INCLUSION STATUS

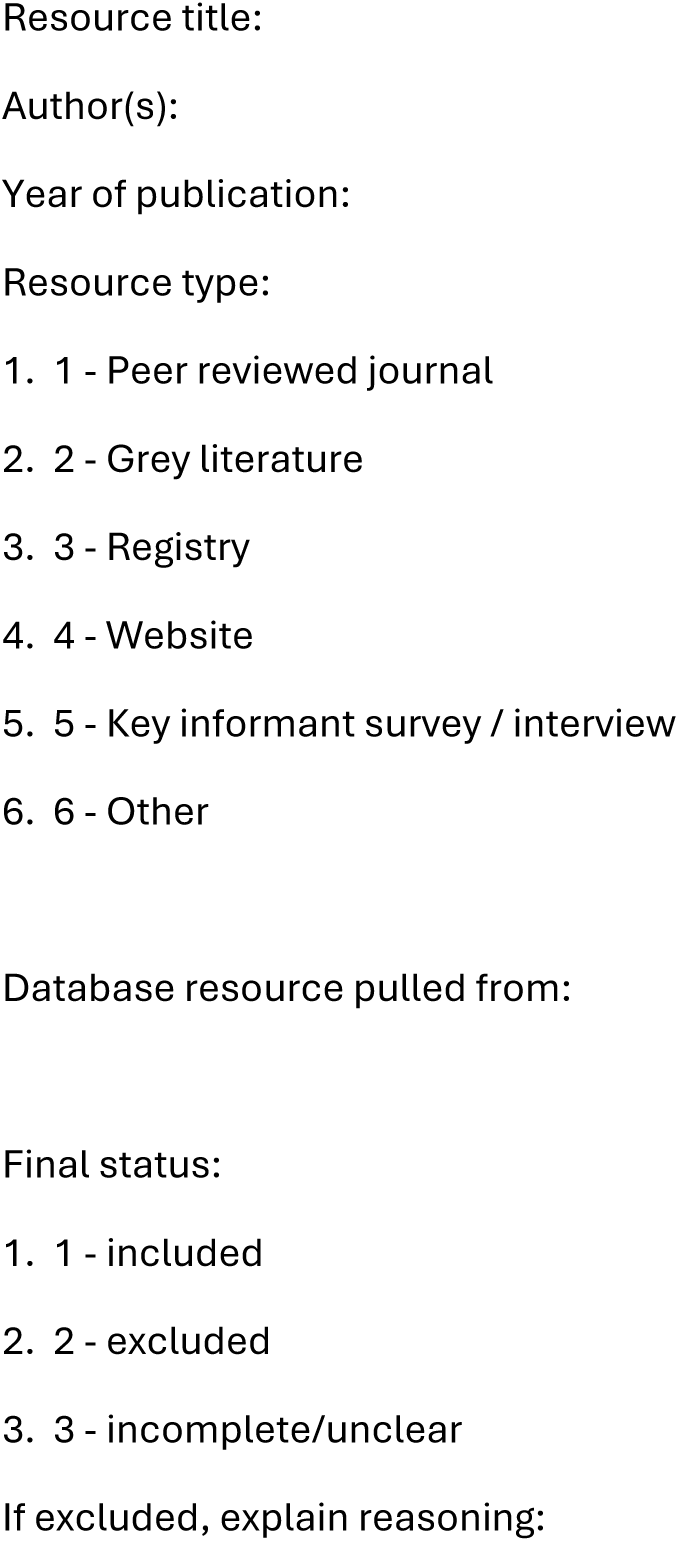

#### PART 2: REGISTRY INFORMATION OR OTHER RESOURCE

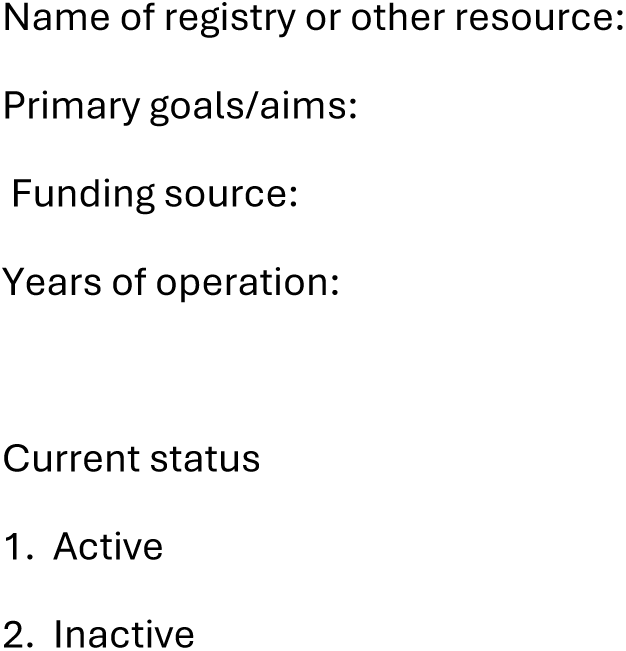

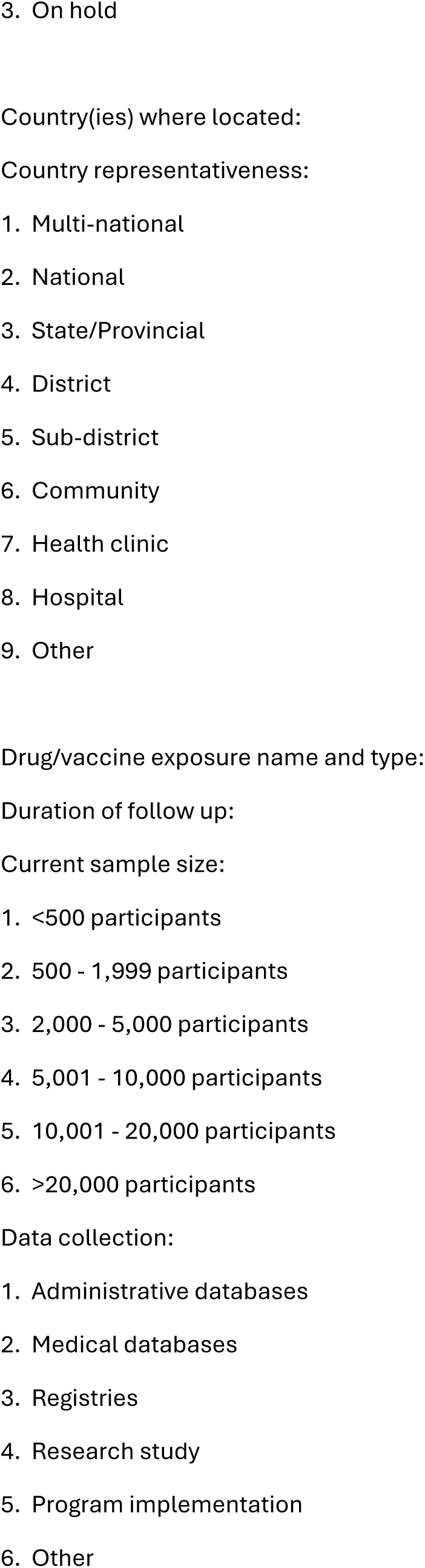
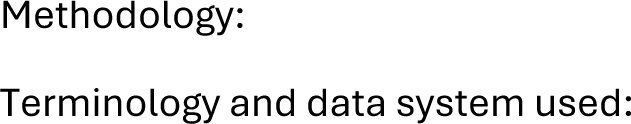

#### PART 3: CHARACTERISTICS OF INCLUDED POPULATION

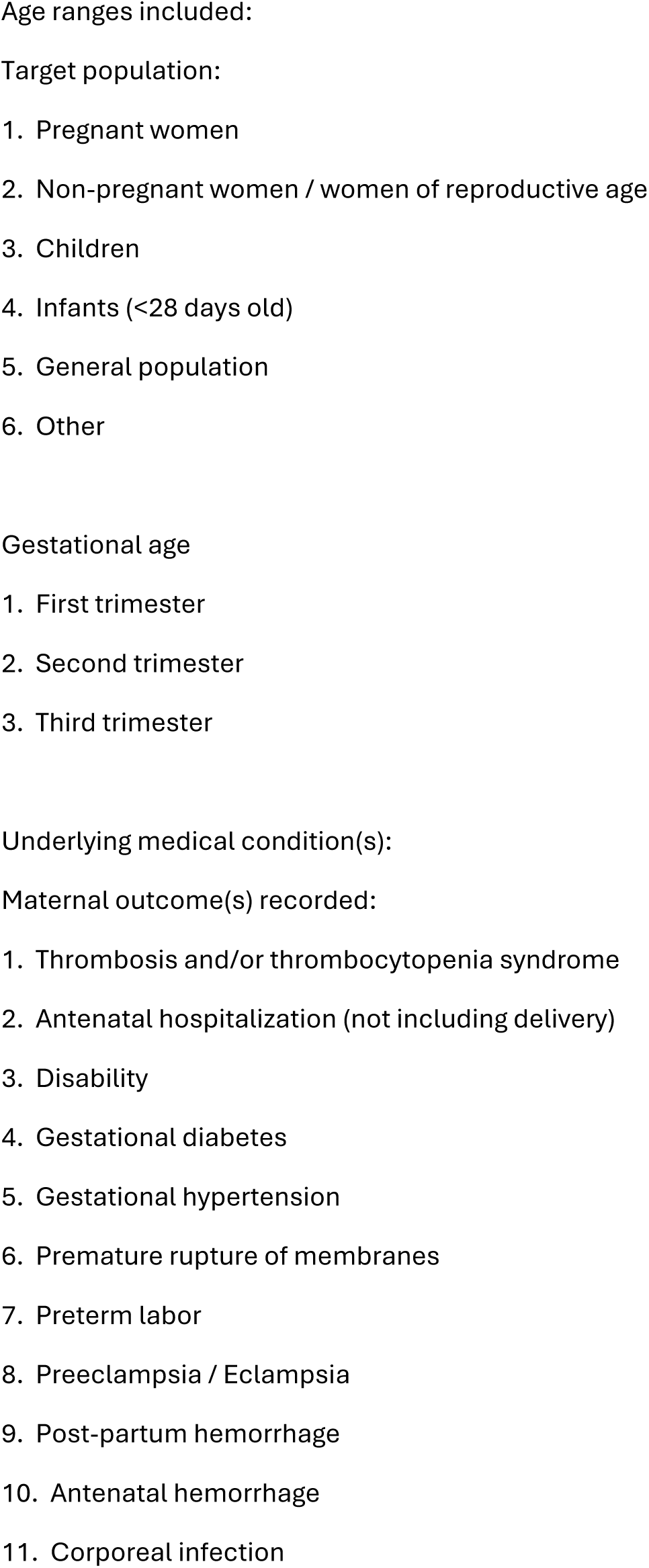

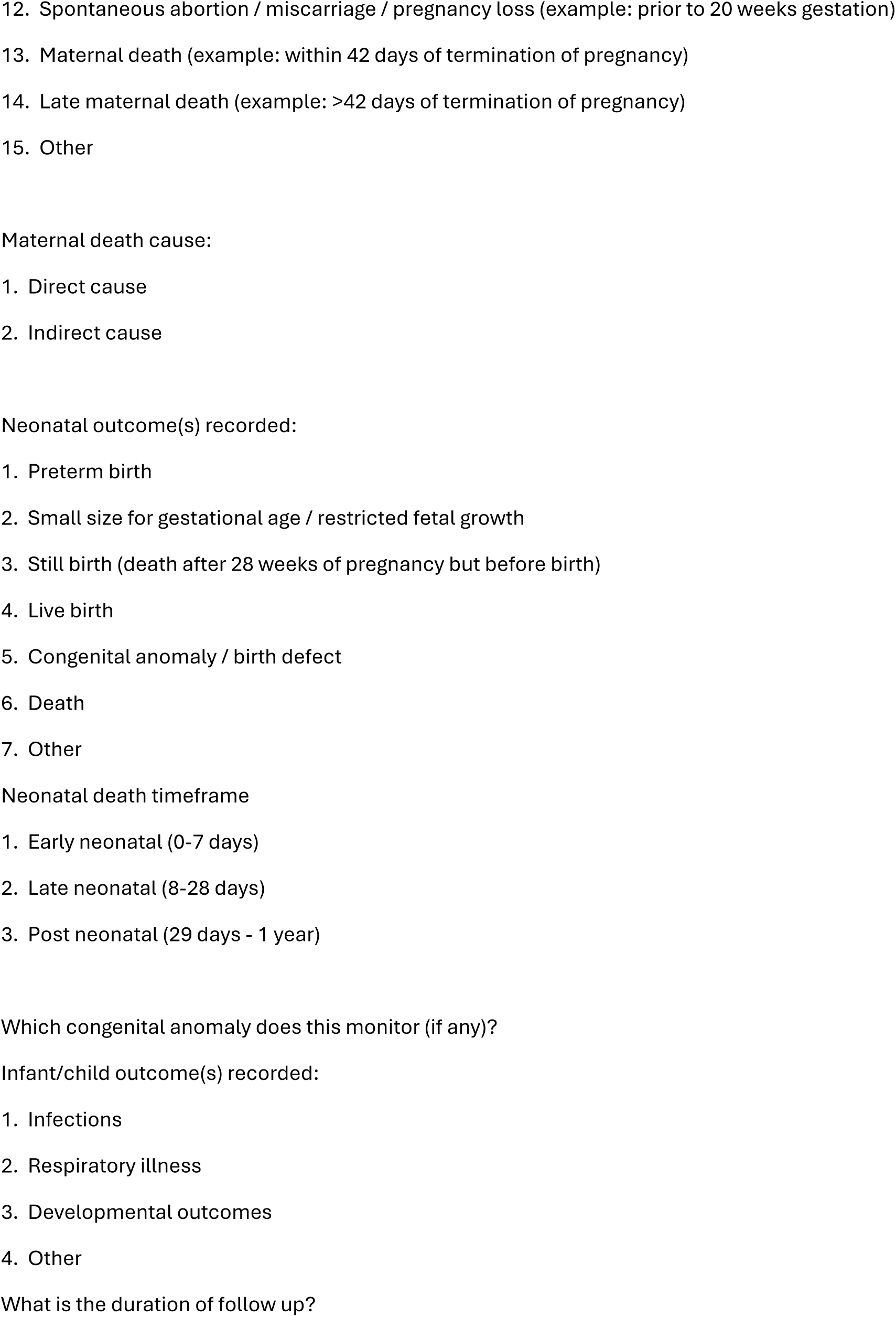

#### PART 4: KEY FINDINGS

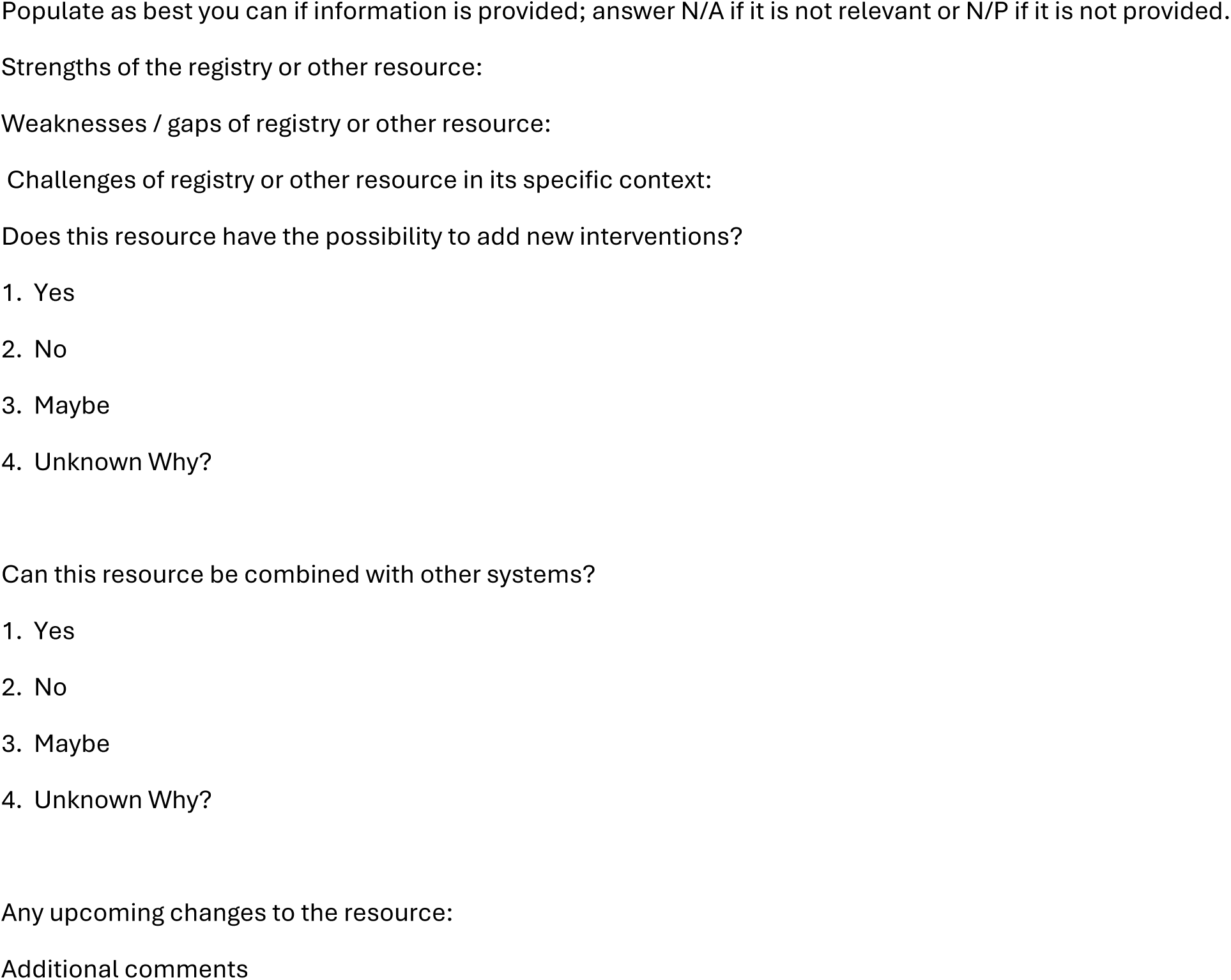

#### PART 1: STUDY IDENTIFICATION / INCLUSION STATUS

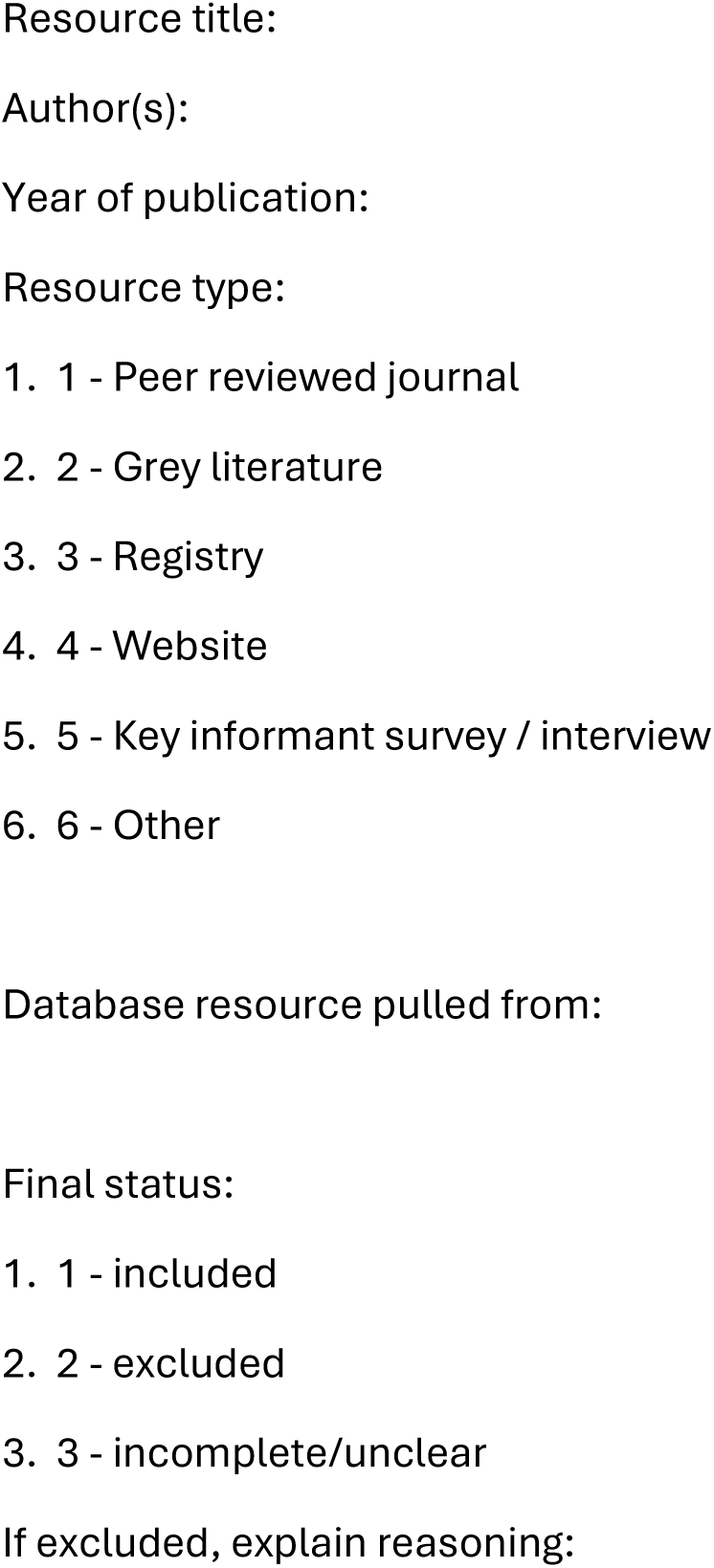

#### PART 2: REGISTRY INFORMATION OR OTHER RESOURCE

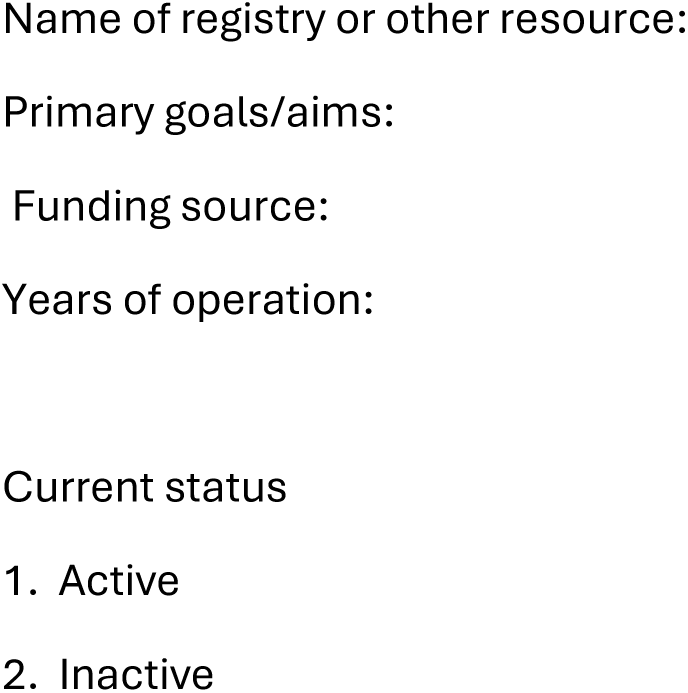

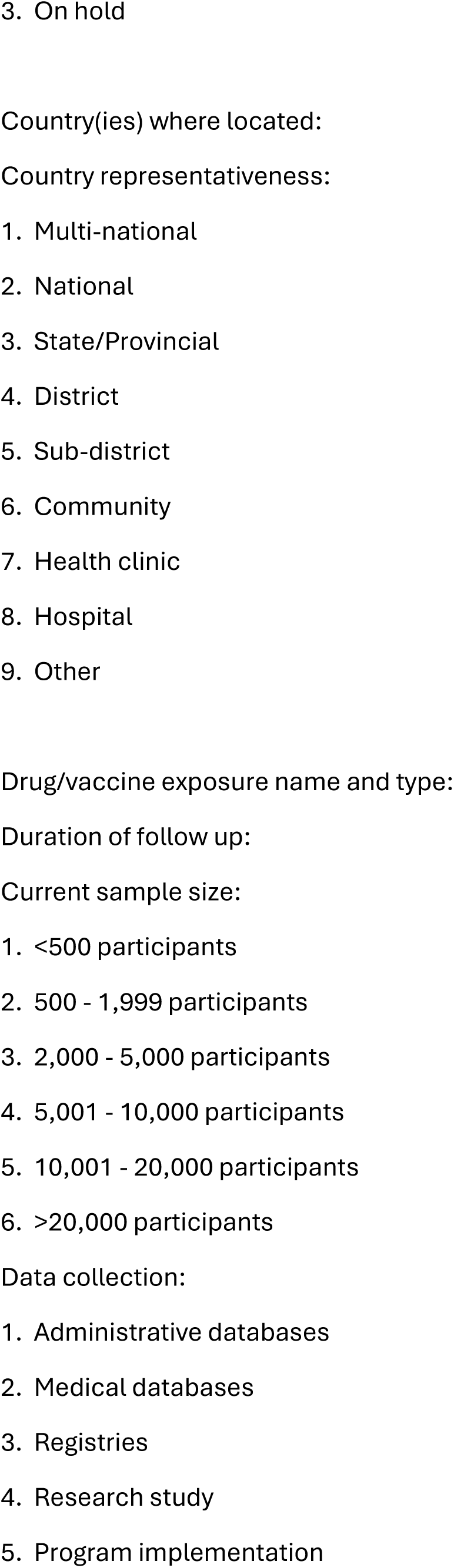

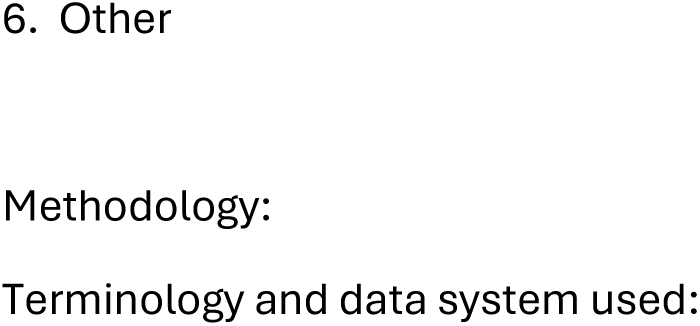

#### PART 3: CHARACTERISTICS OF INCLUDED POPULATION

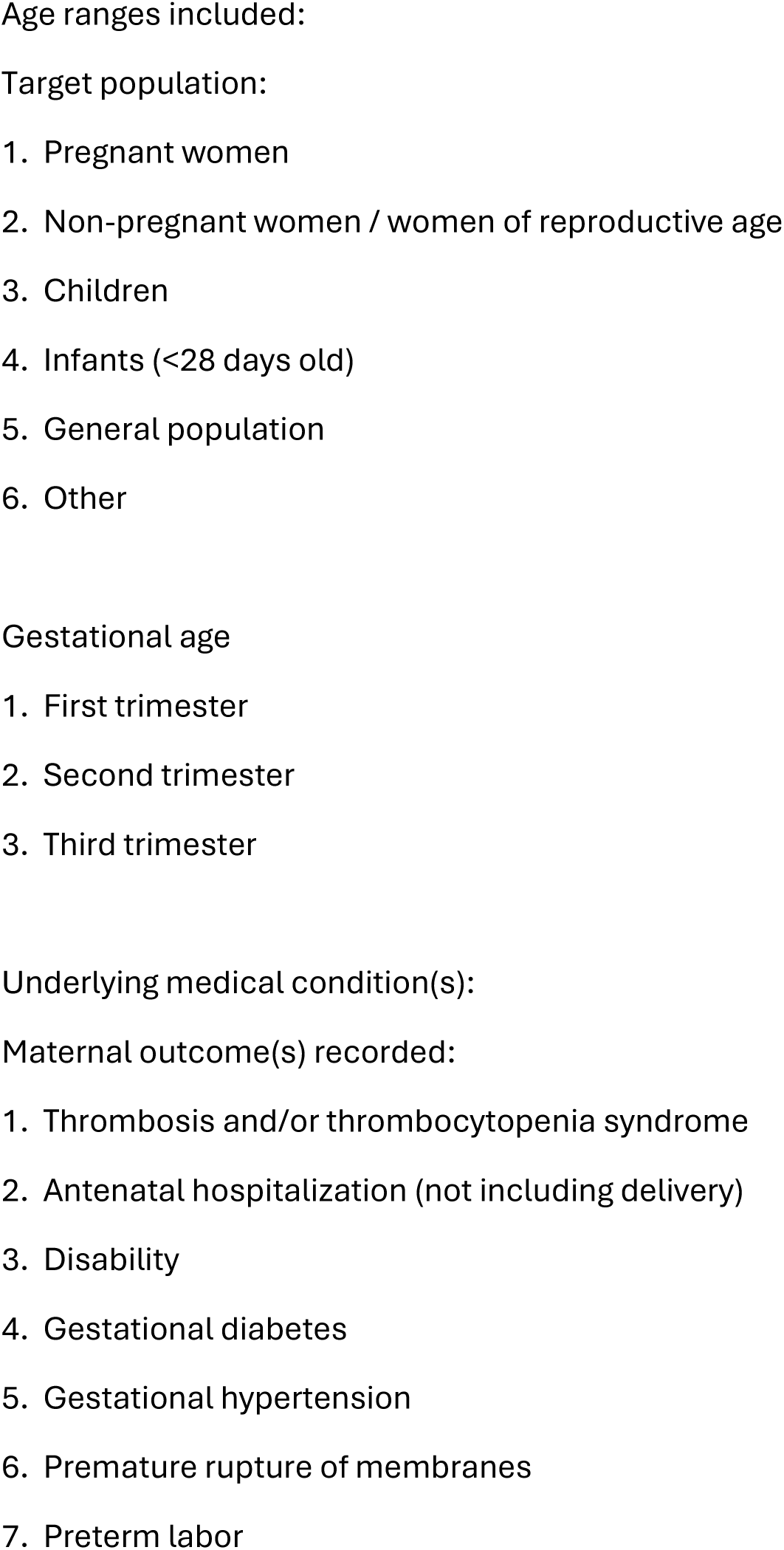

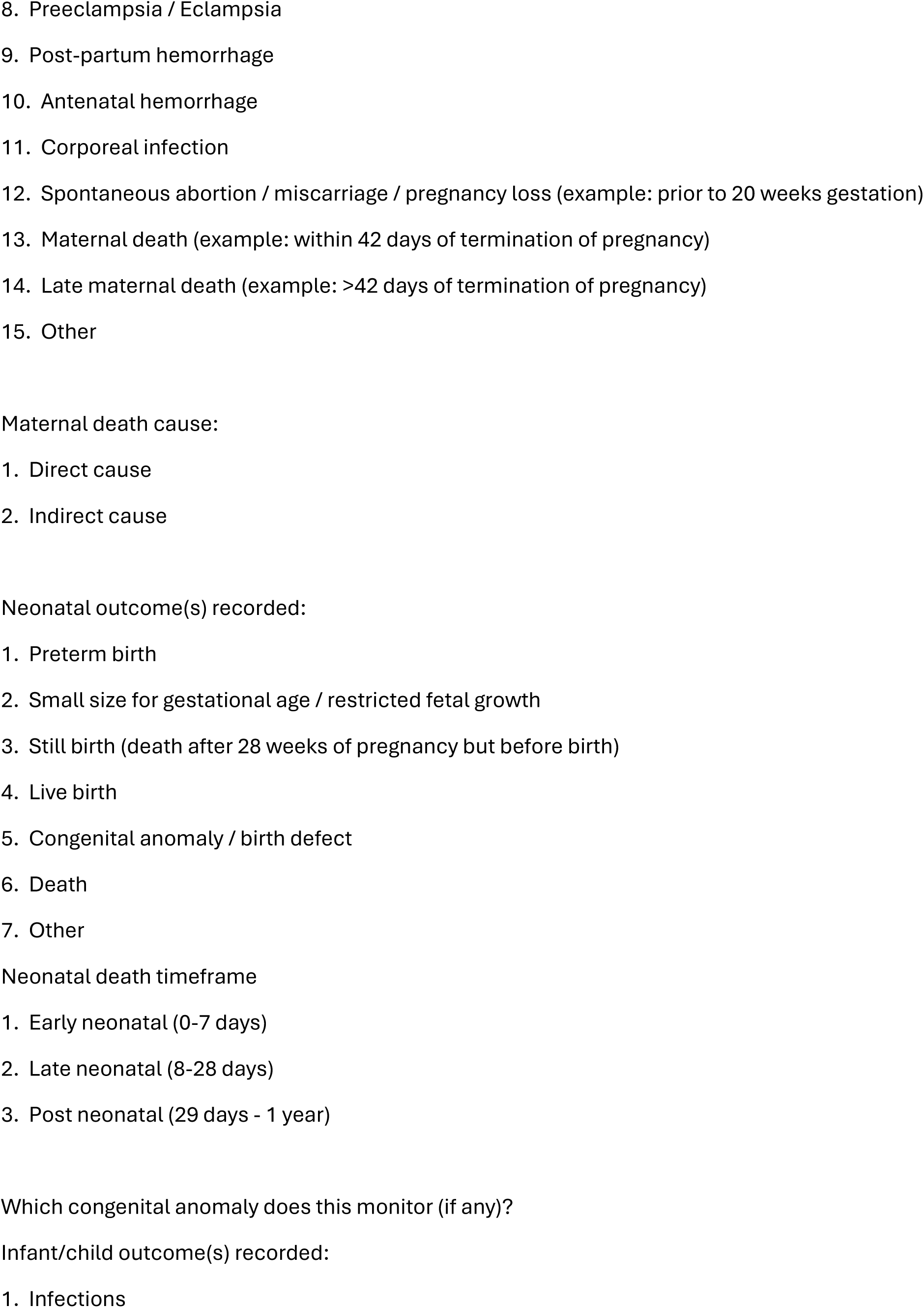

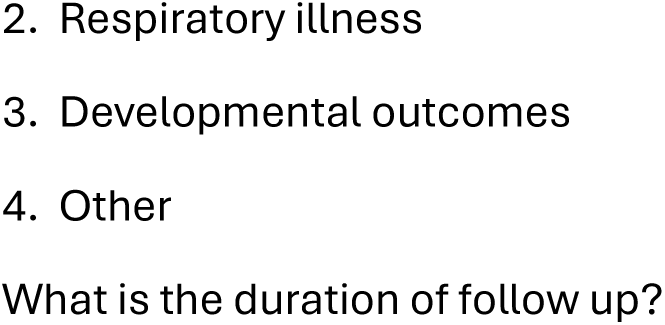

#### PART 4: KEY FINDINGS

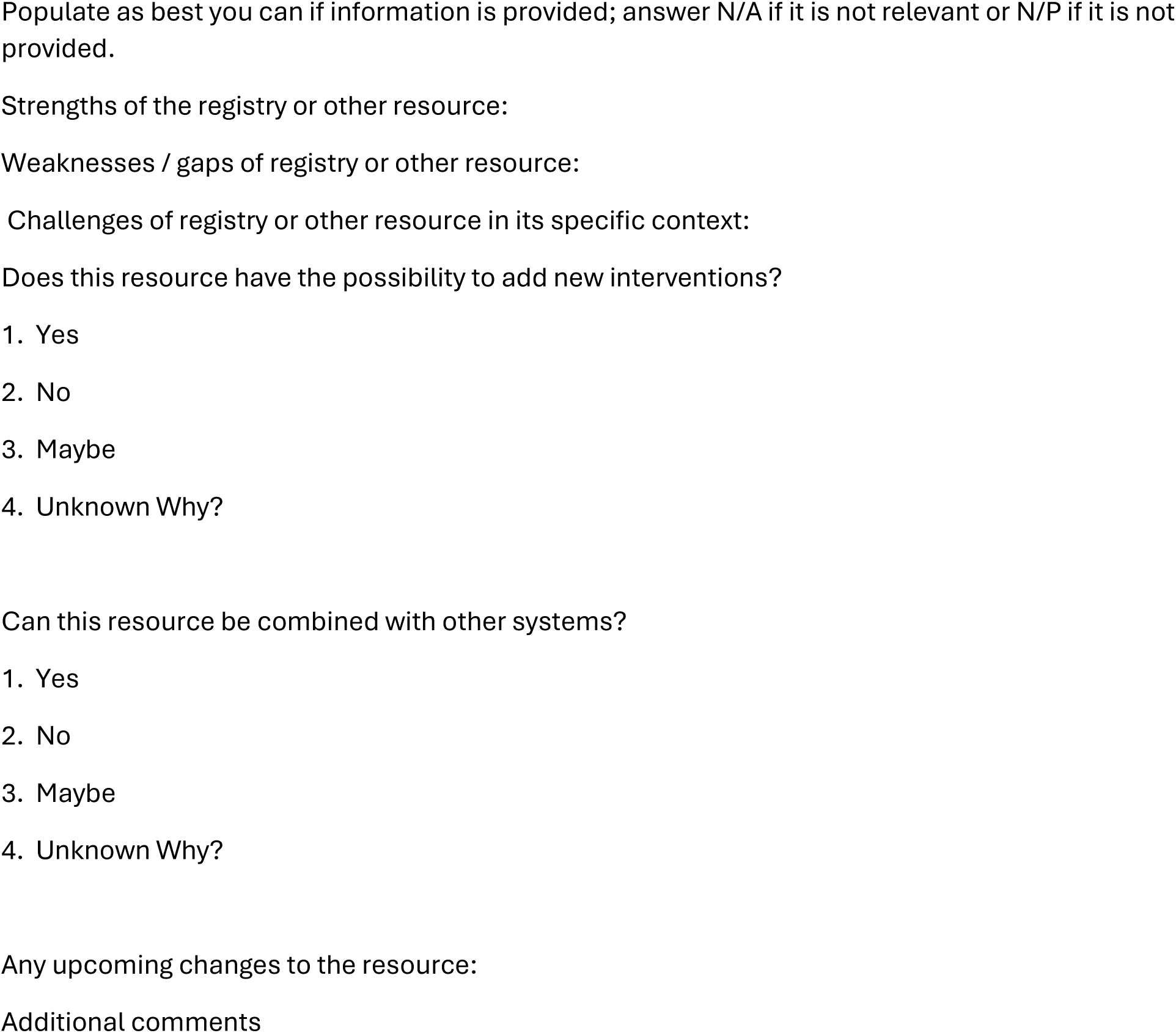

### Key informant survey and interview

#### Pregnancy Exposure Data and Resources Stakeholder Survey

We are conducting a landscape analysis in collaboration with WHO to identify current and recent resources, including pregnancy exposure and surveillance registries, databases, cohort surveys, and routinely collected data, that record exposure to medicines and vaccines during pregnancy and maternal and perinatal outcomes in low- and middle-income countries (LMICs). We are asking for your help in identifying examples of these resources. We may follow up with you to discuss the appropriateness or fit for purpose of the resource you identify. Our goal is to understand what is currently available in LMICs and make connections for future evaluation of maternal use of medicines and vaccines in the product pipeline.

You have been identified as someone who is knowledgeable about or involved with these resources in LMICs. Please complete the following form for each resource you know of. We will ask for your name and contact information so that we may follow up with you for further information, if necessary. All of the personal information you provide will be kept confidential. When we report our findings, if we need to mention something you have said or information you have provided, we will refer to you by a unique study ID to keep your identity confidential. By submitting the form, you are agreeing to participate and allow us to use the information you have provided.

Please fill out the following questions to the best of your knowledge. If there are any specific points that are not included as options in the dropdown menus that are relevant to the resource, please type in the answer and hit “enter”.

#### Participant details

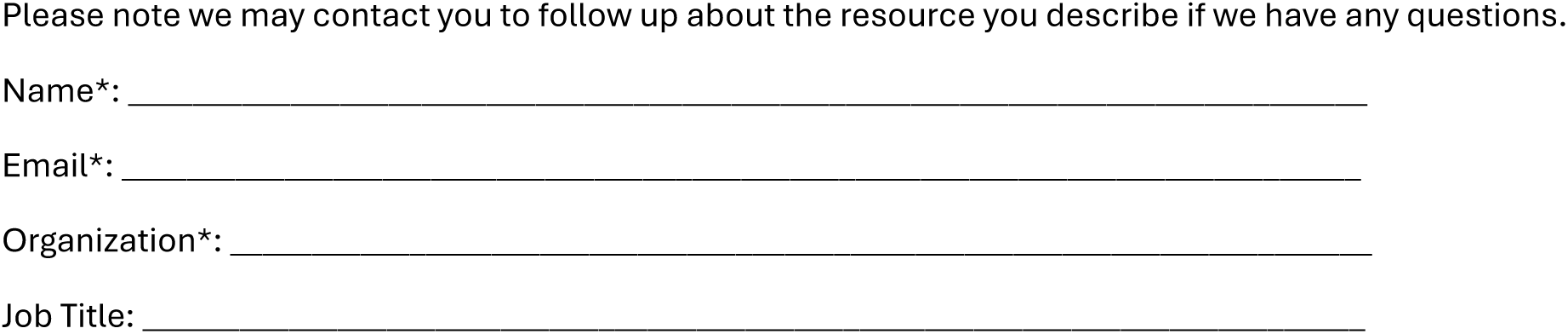

#### Resource details

Below we will be asking you to fill in information about any resources you are familiar with as outlined above. If you know of multiple resources that should be brought to our attention, please fill out a separate survey for each resource. As a reminder, resources can include pregnancy exposure and surveillance registries, databases, cohort surveys, and routinely collected data, that record exposure to medicines and vaccines during pregnancy and maternal and perinatal outcomes in low- and middle-income countries (LMICs)

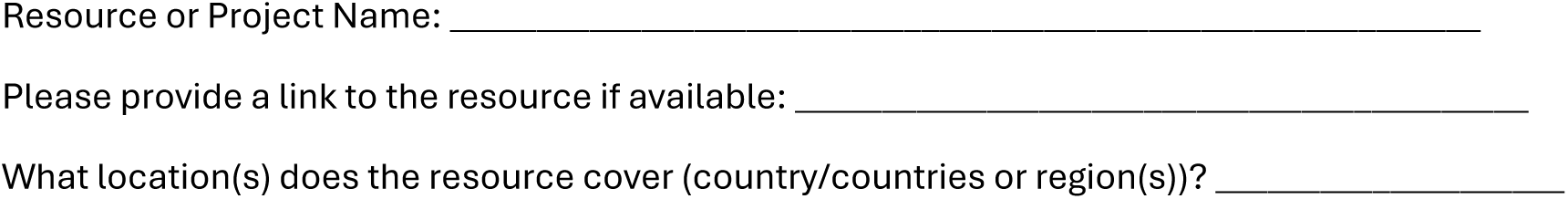

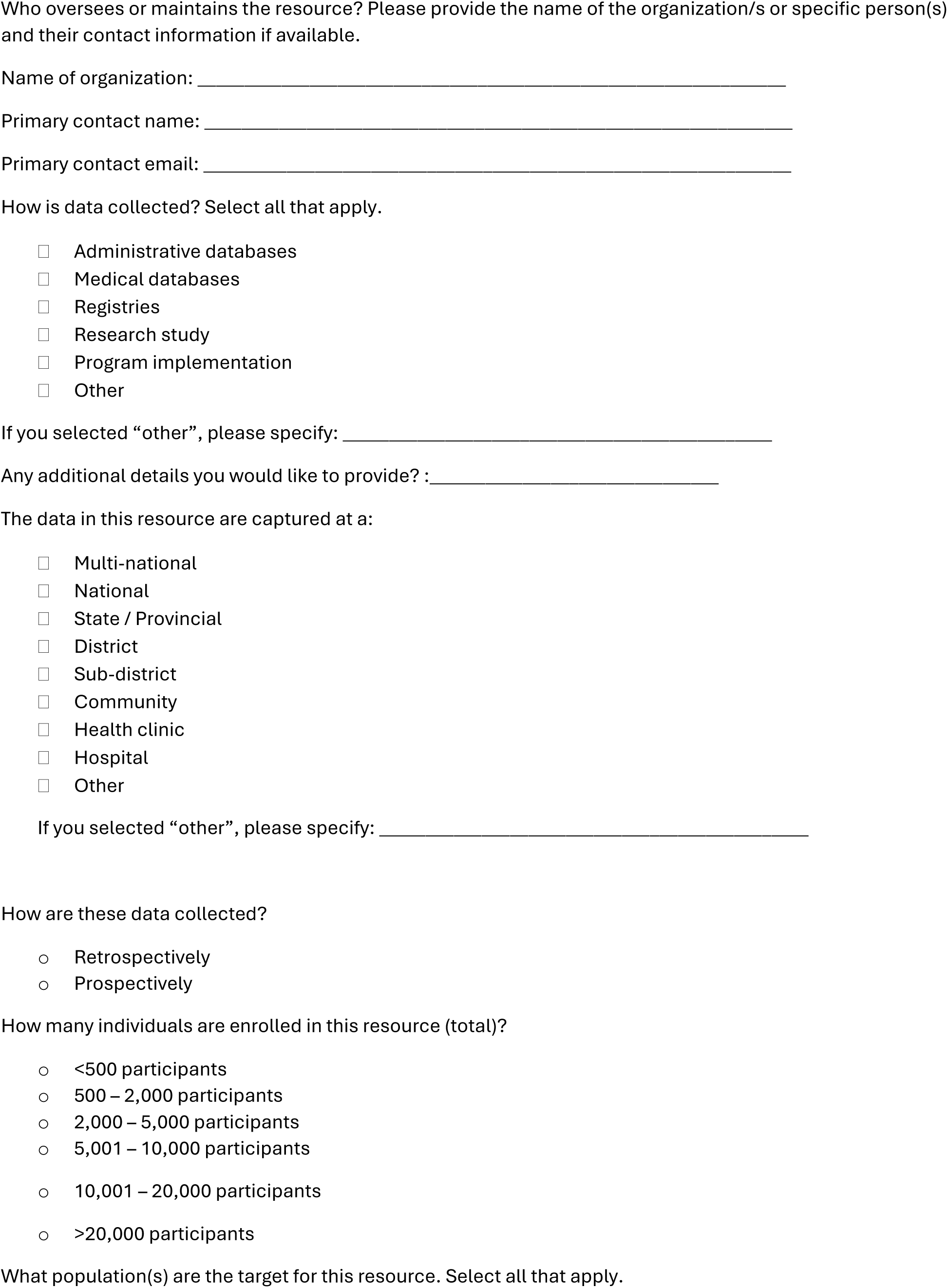

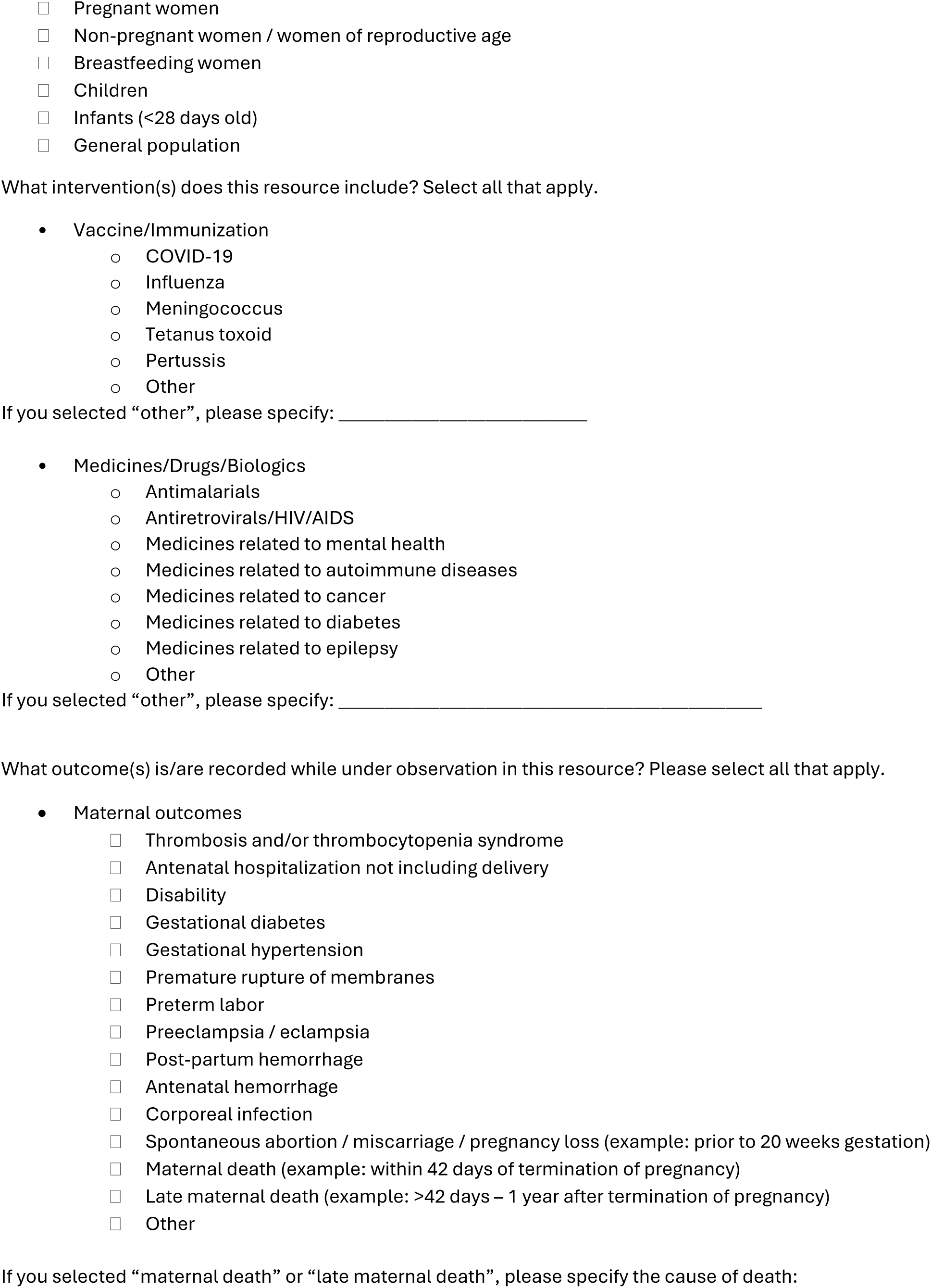

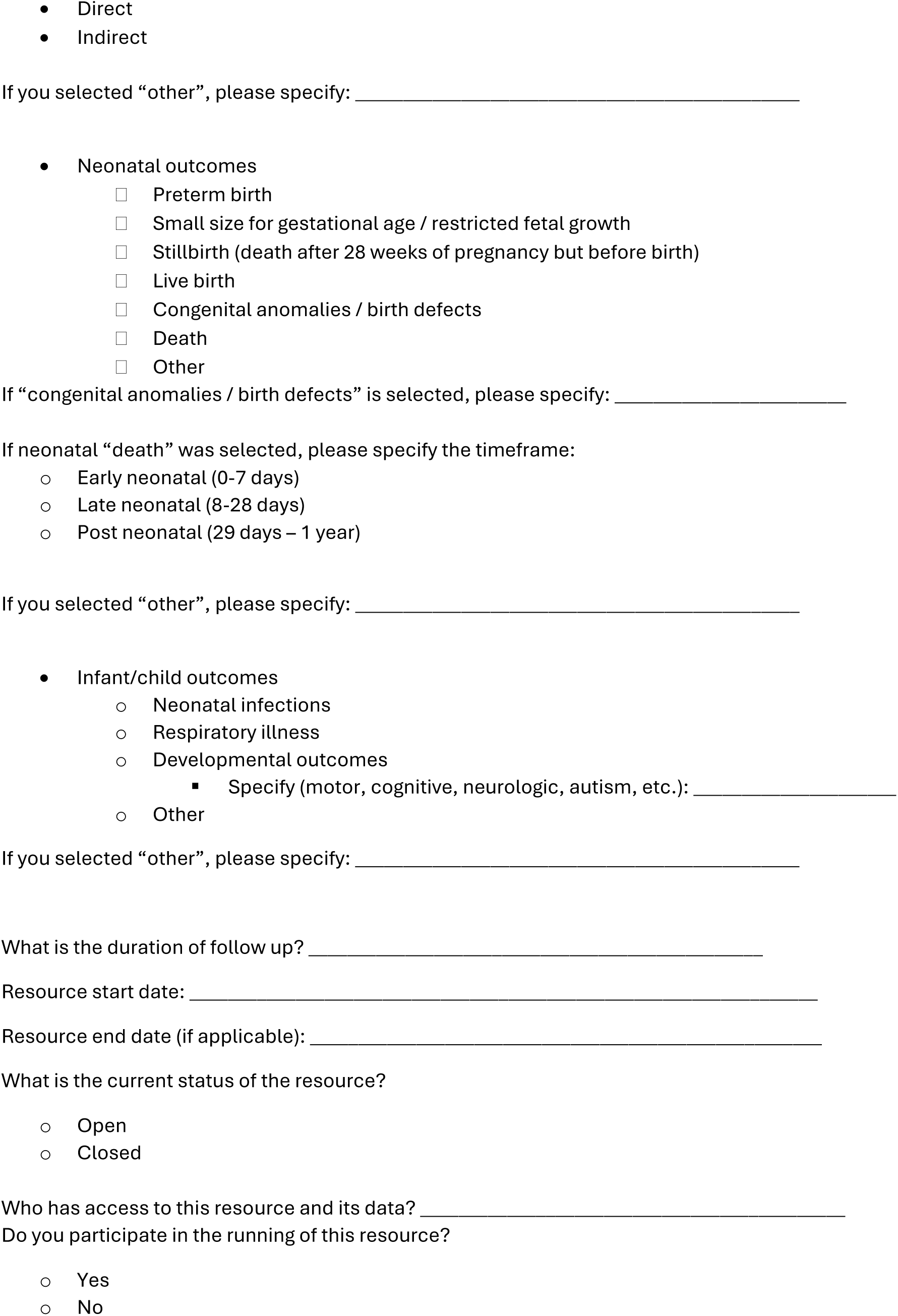

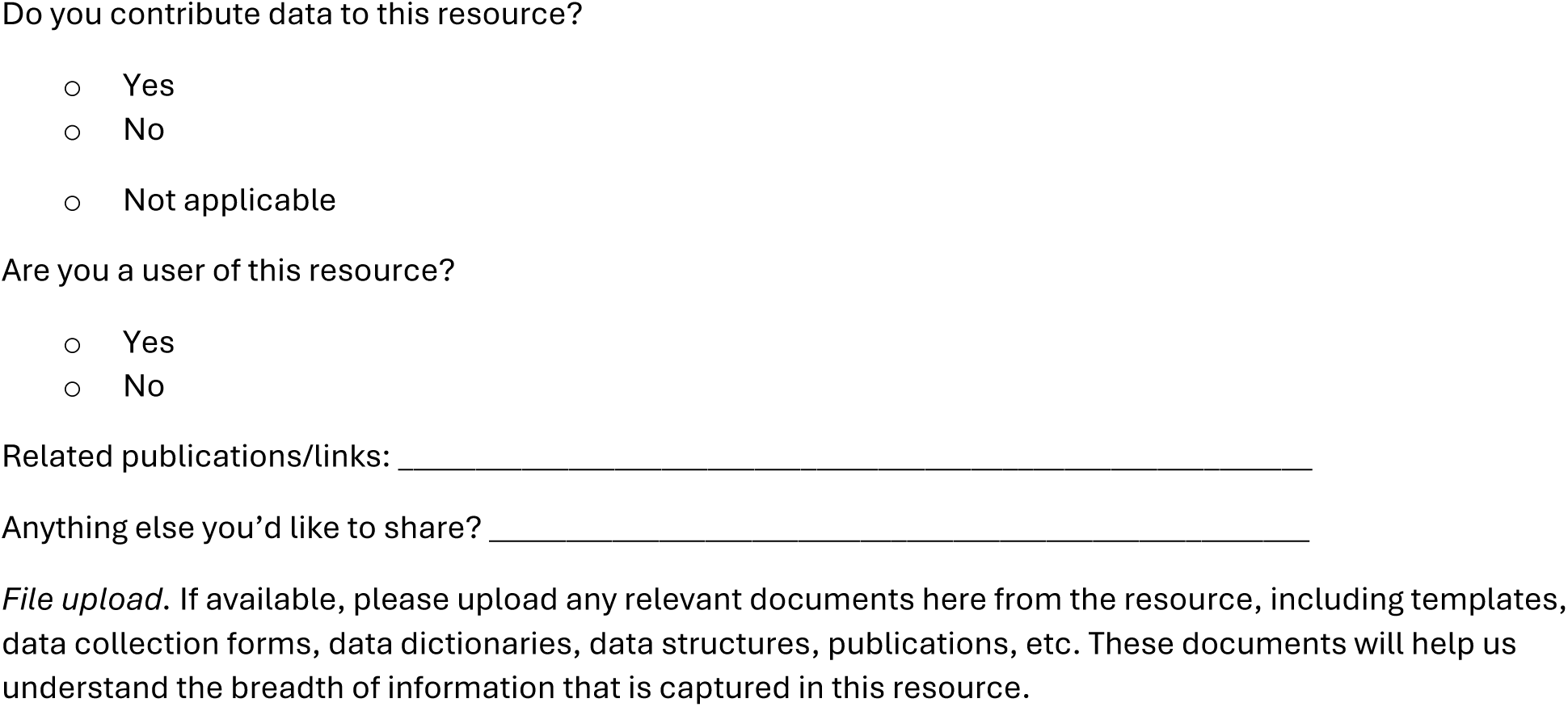

#### PERLA Key Informant Interview

##### Consent script

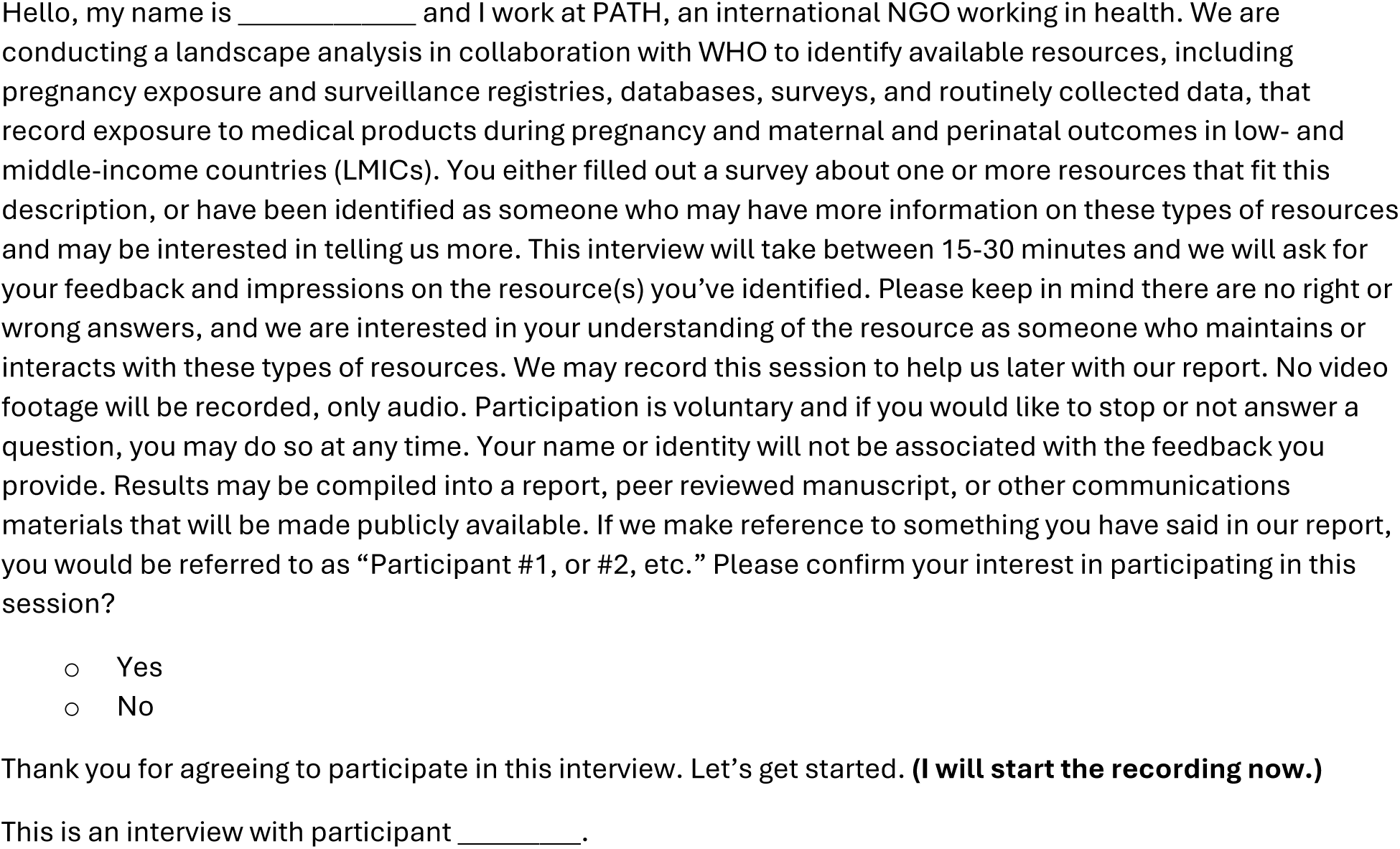

##### Interviewee information

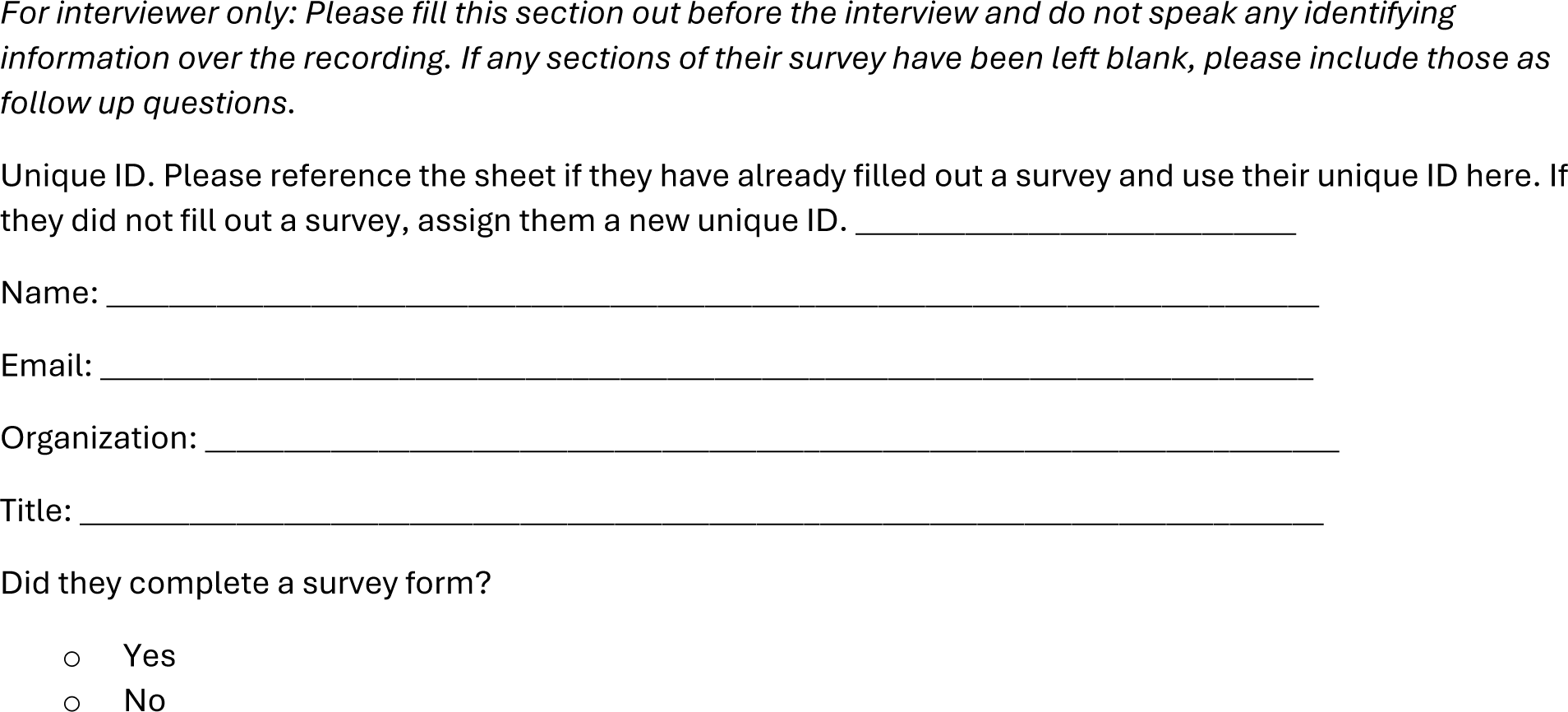

##### Resource information

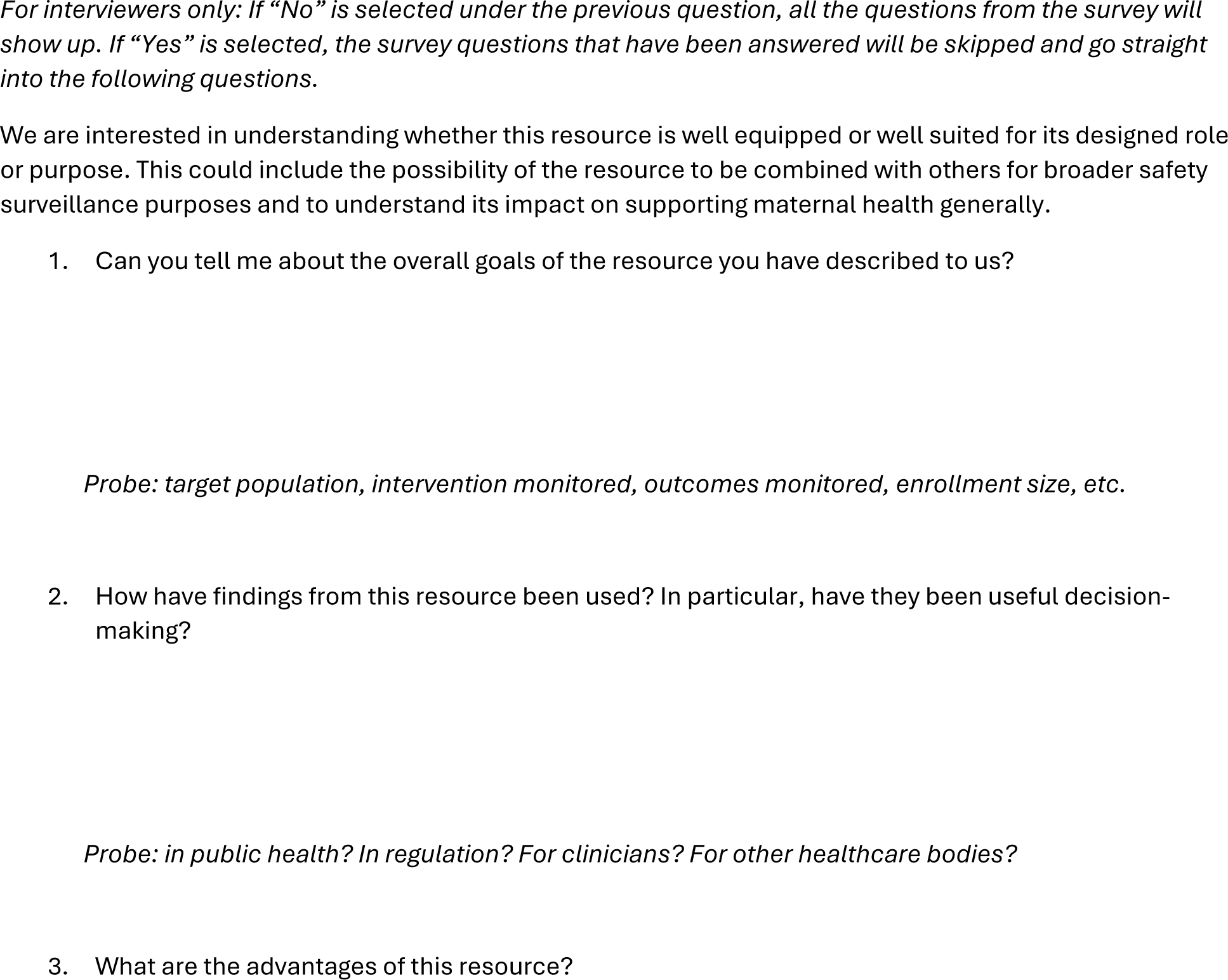

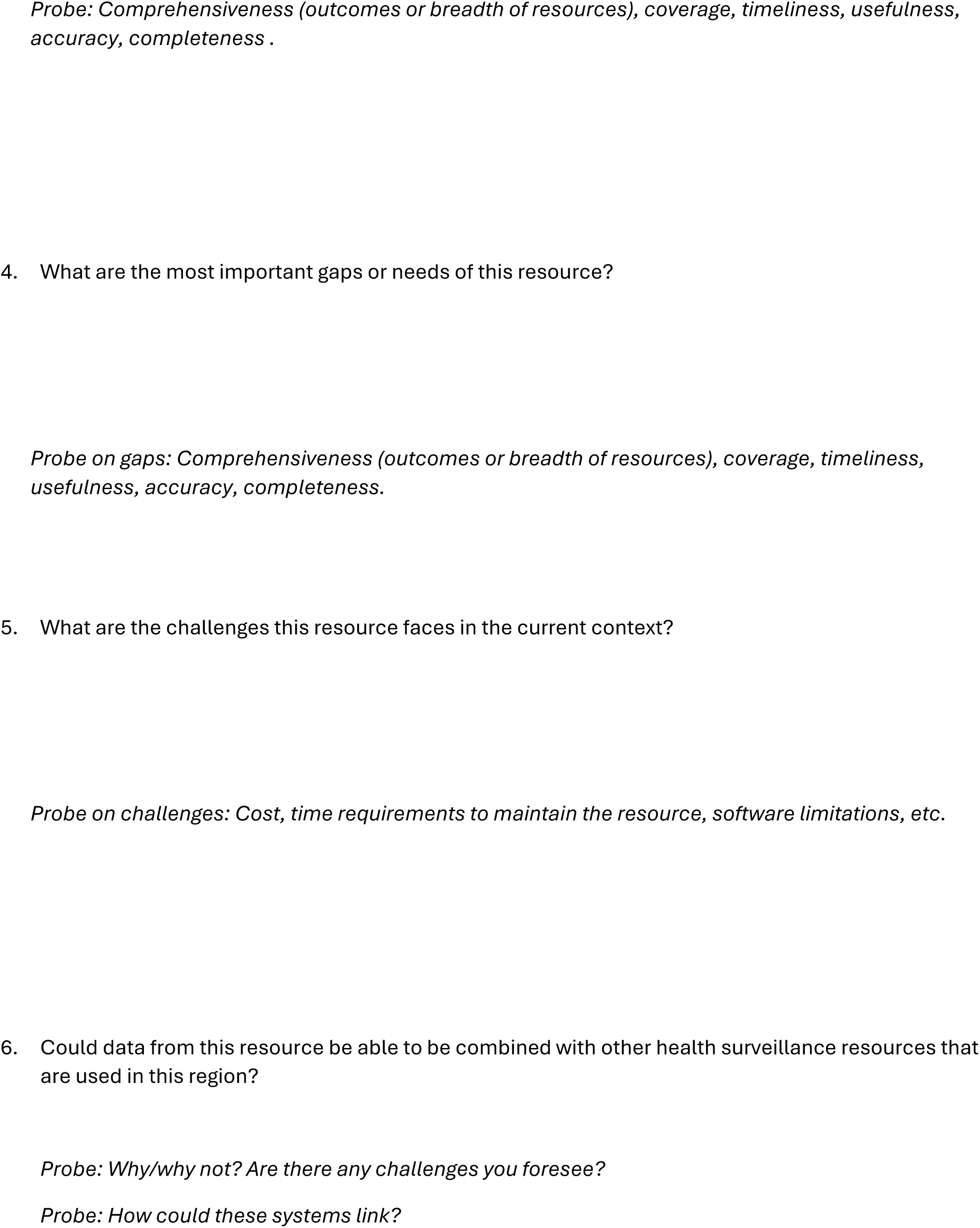

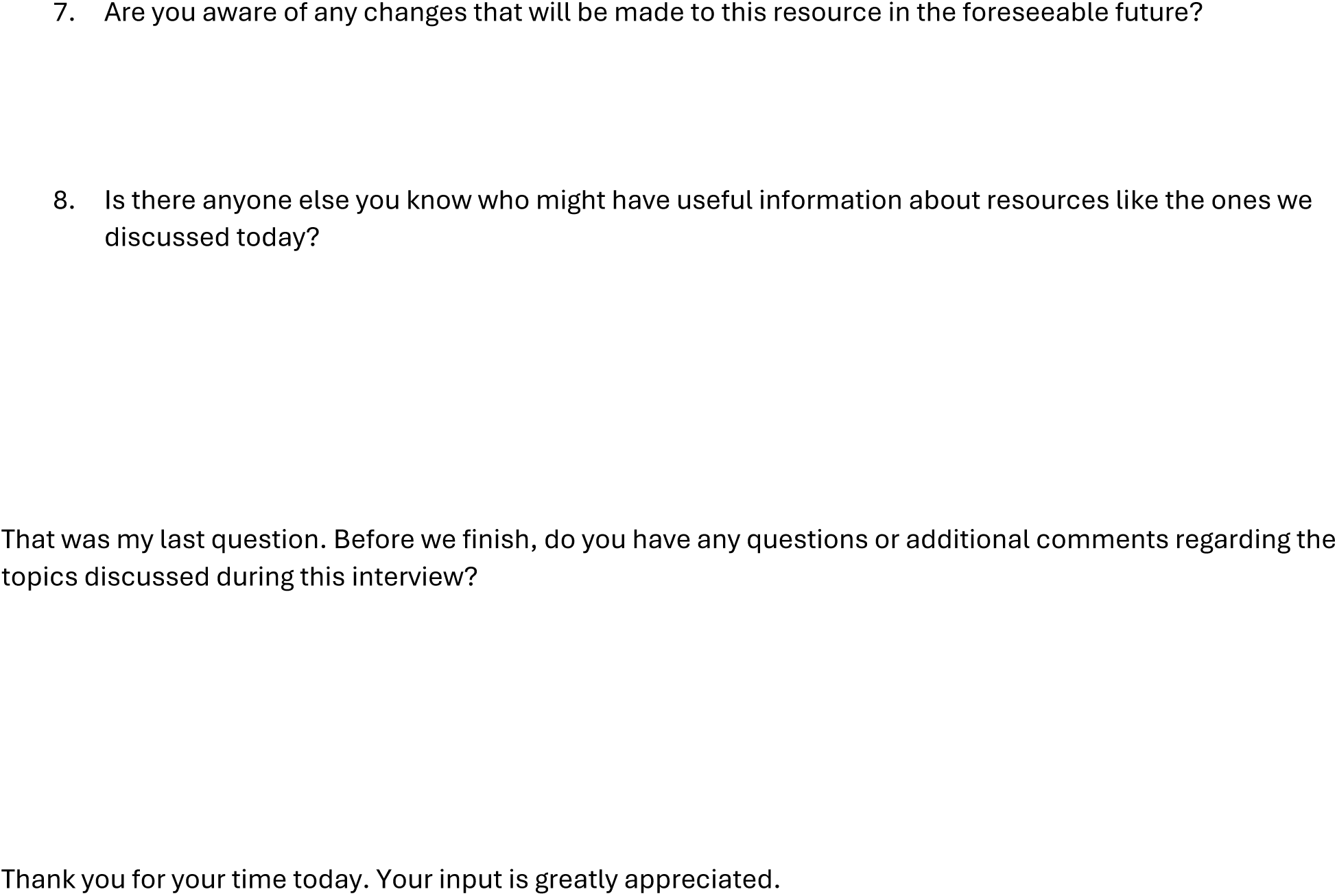

**Table S1.**
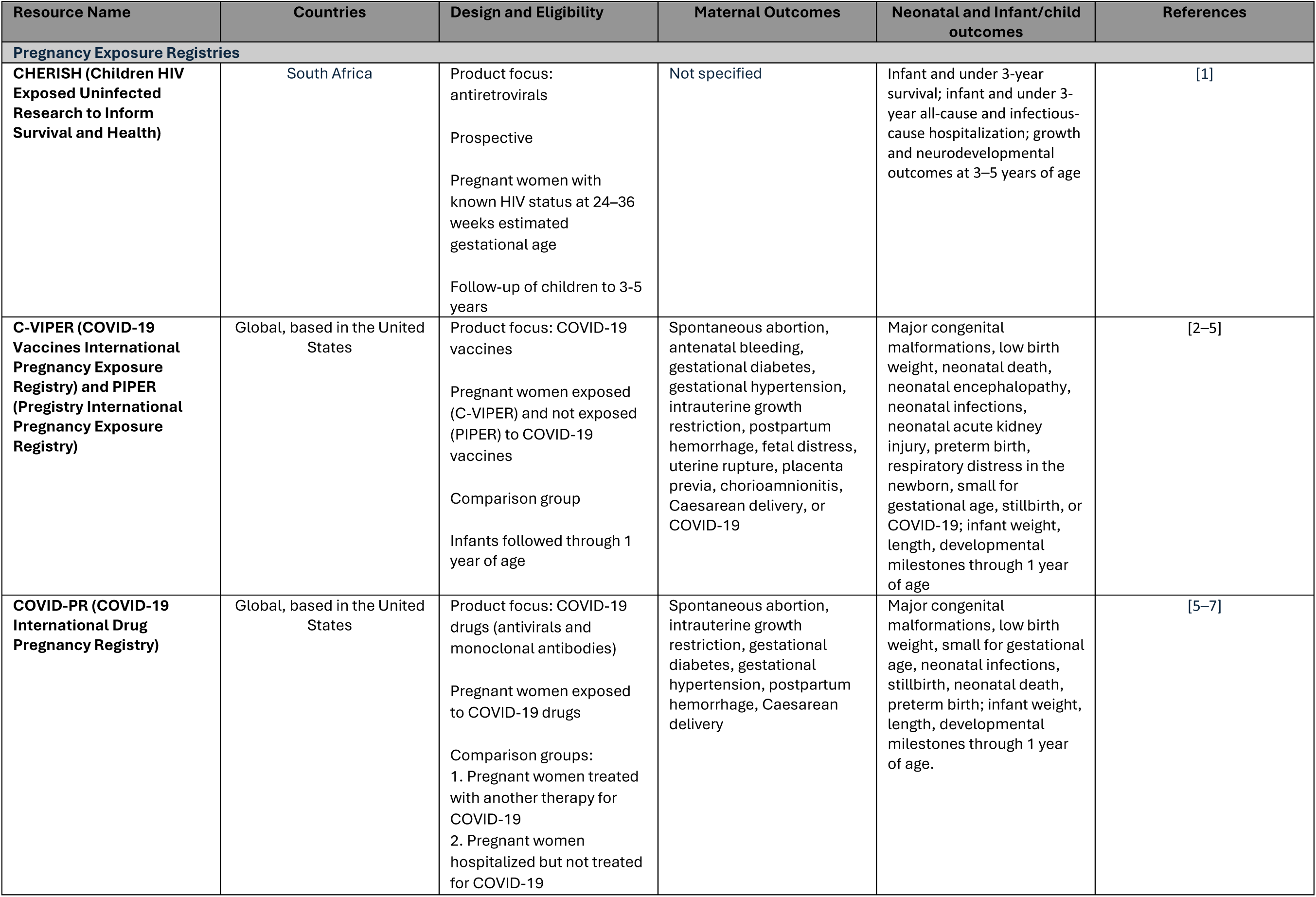
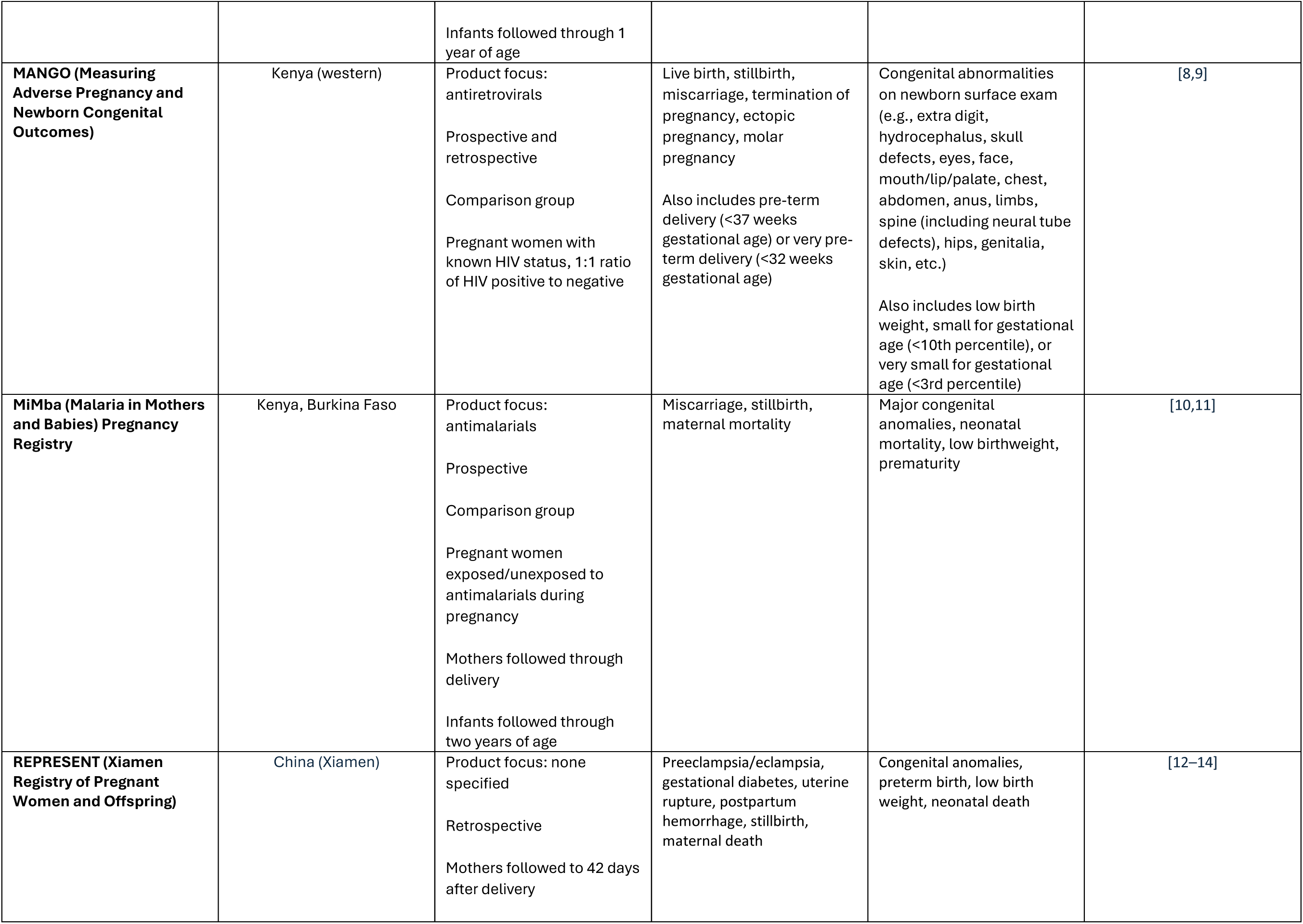
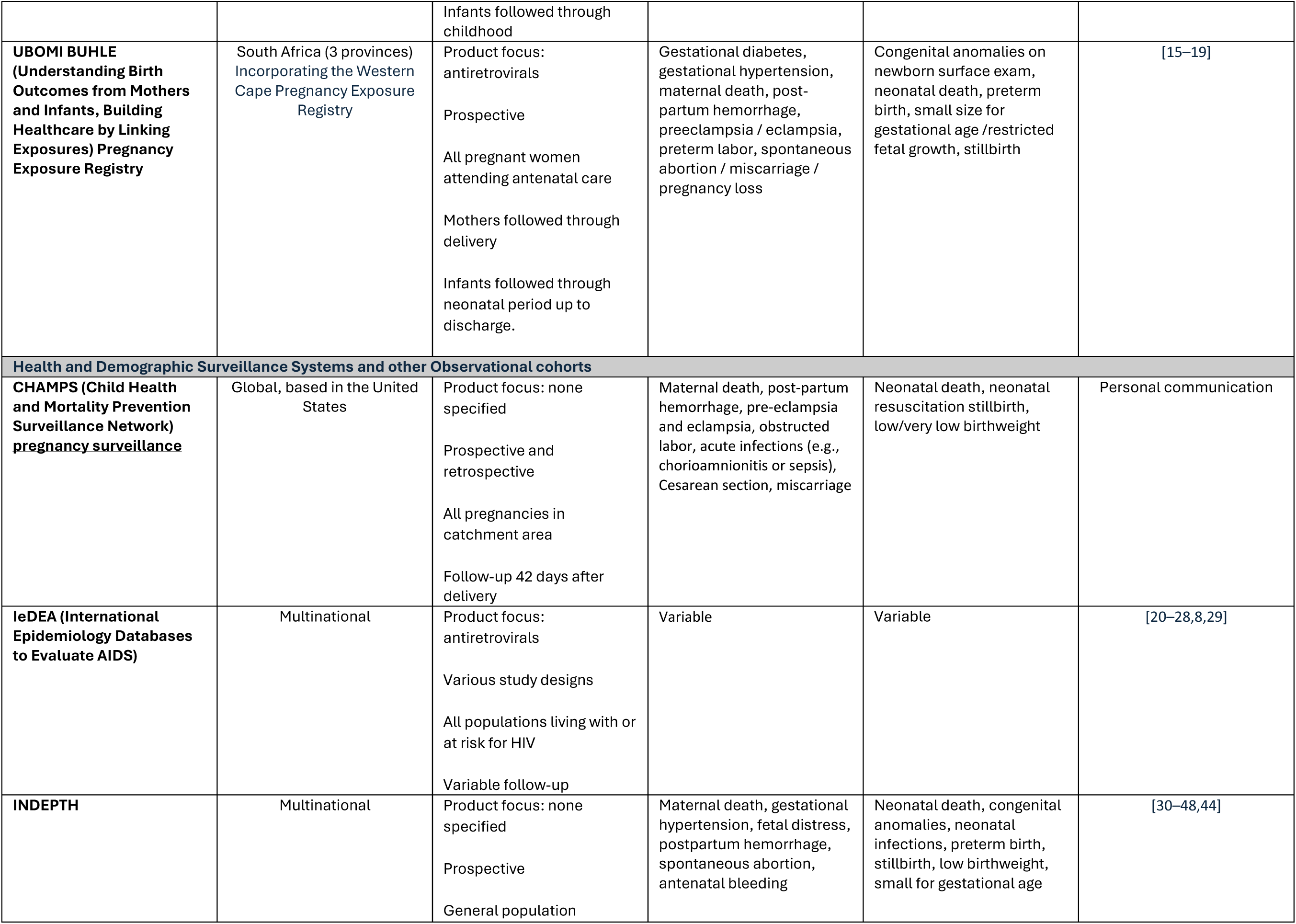
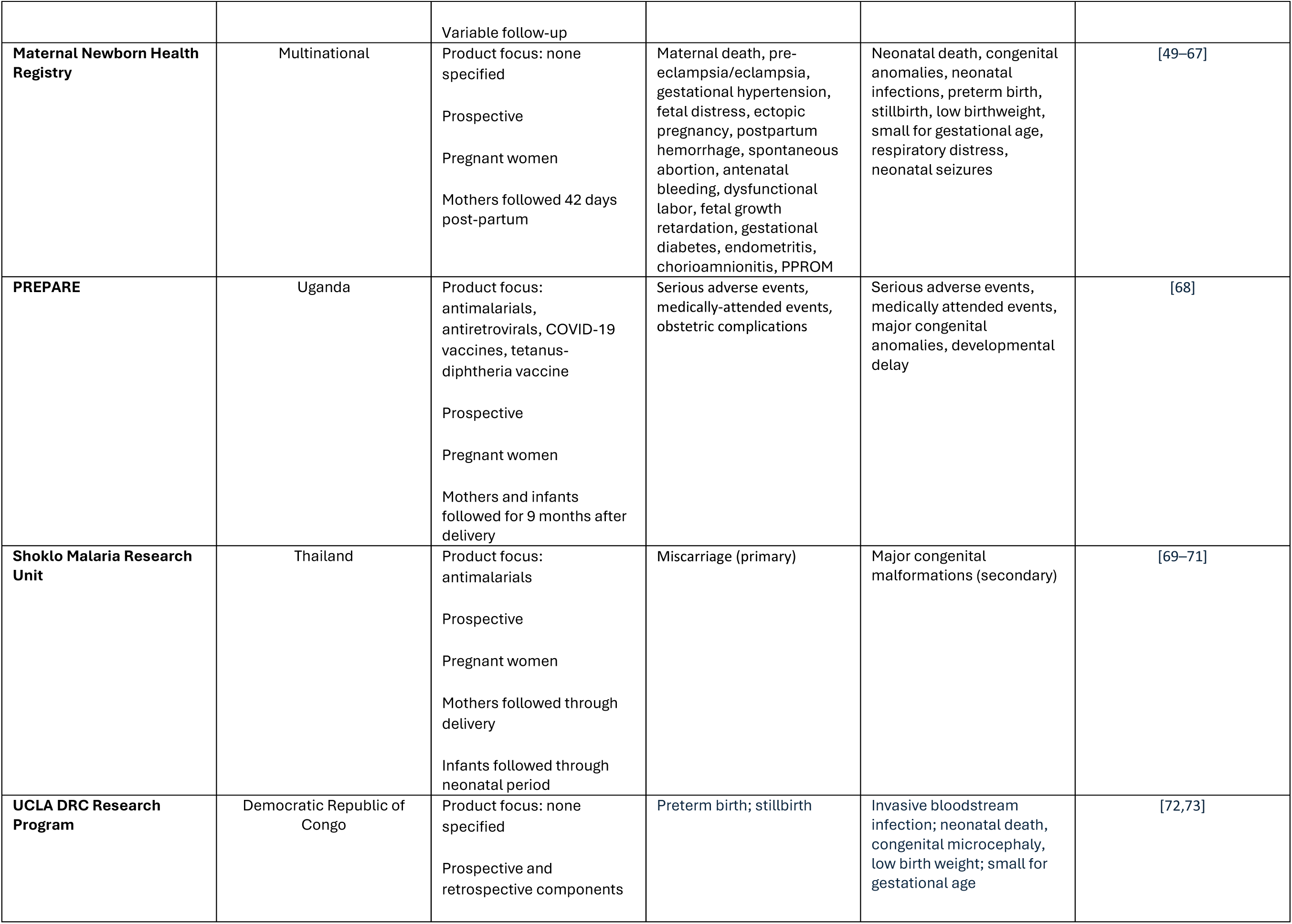
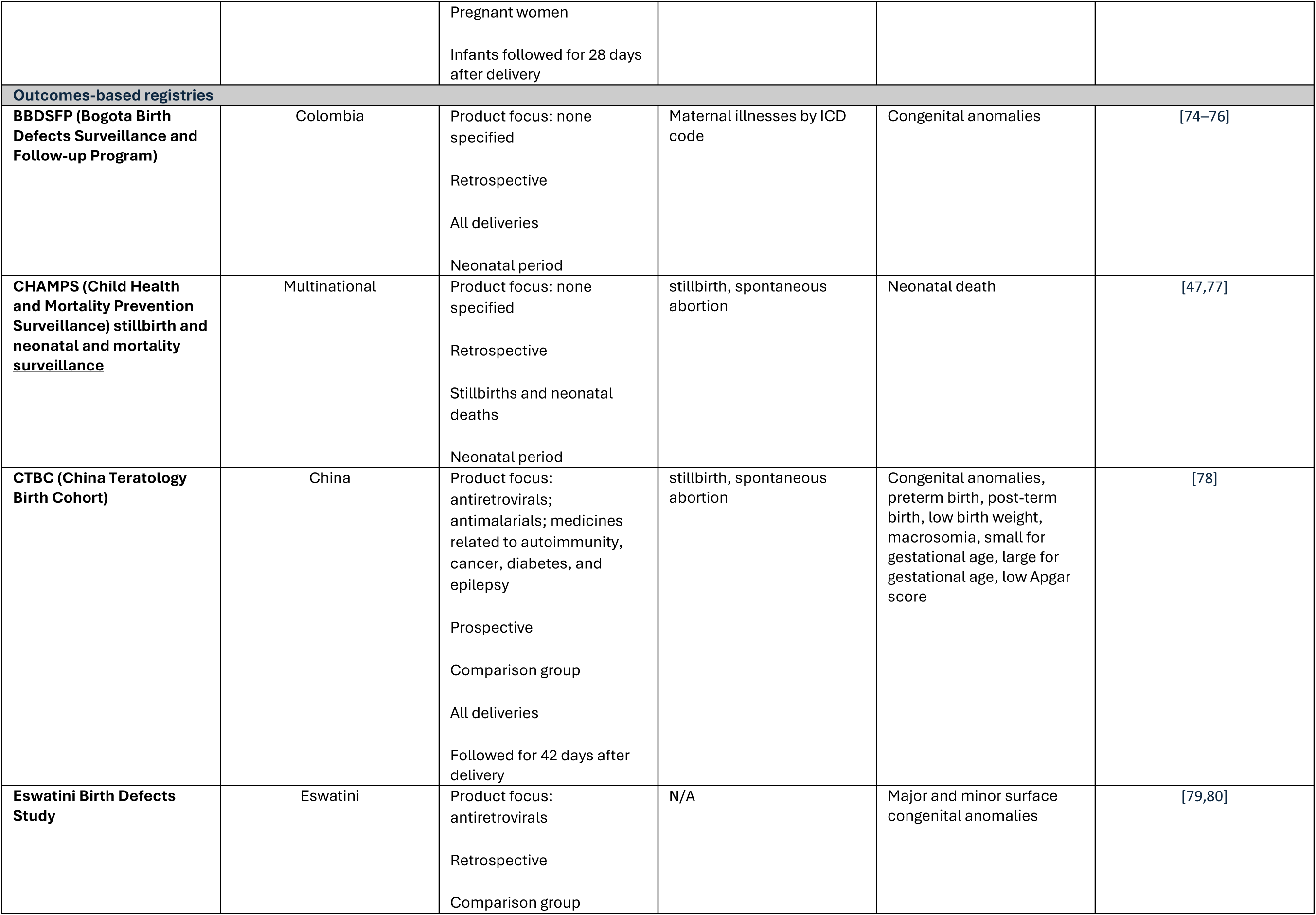
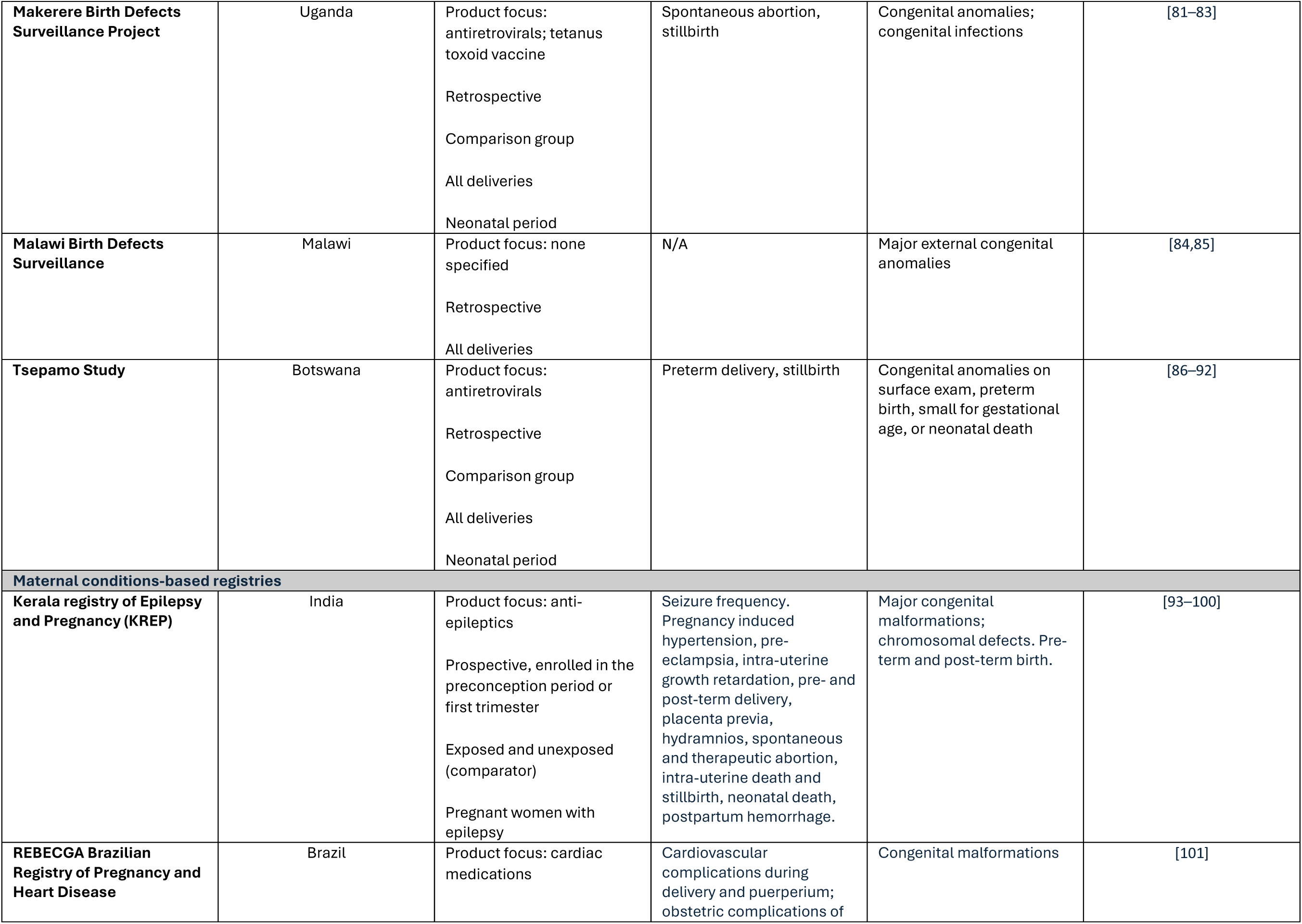
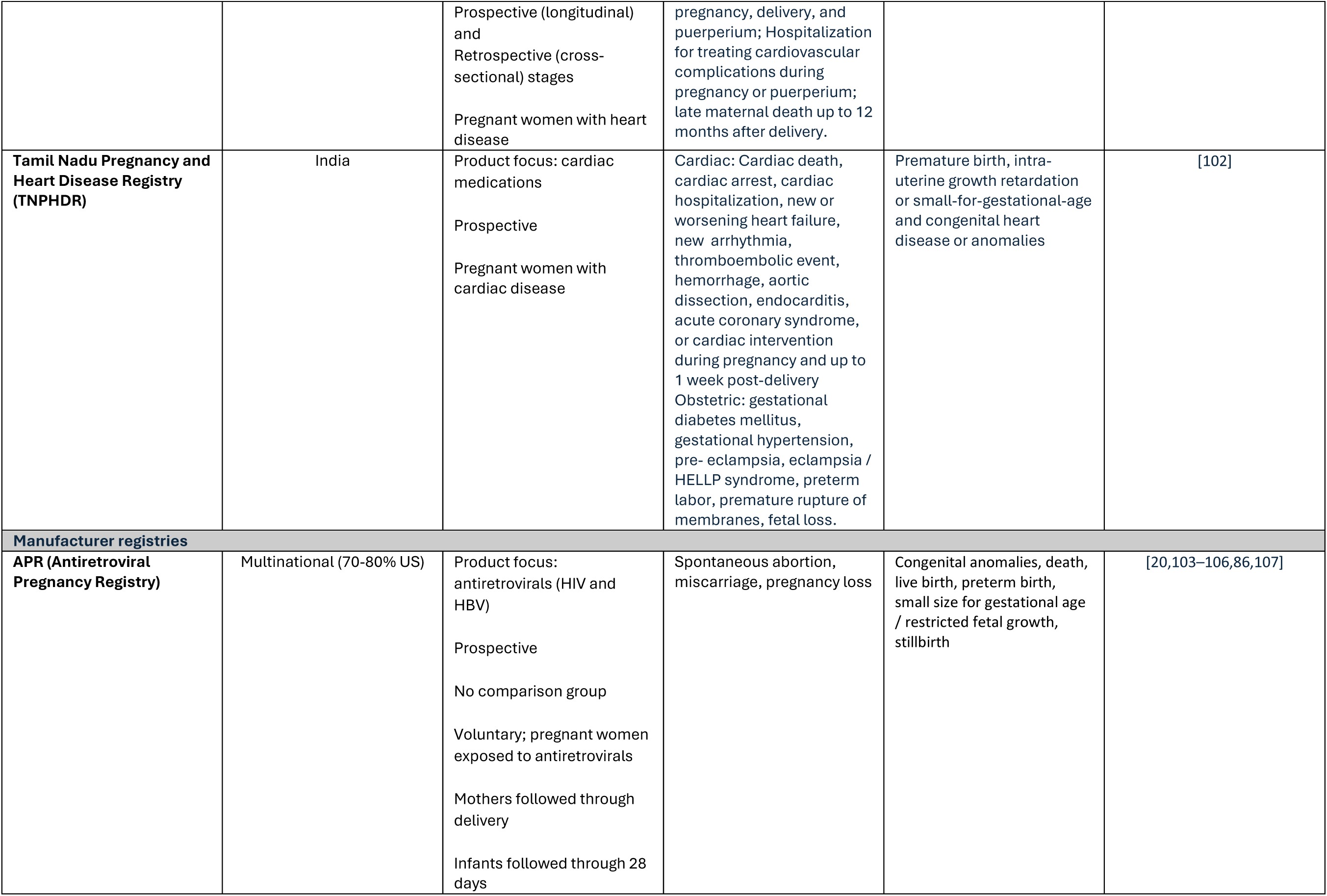
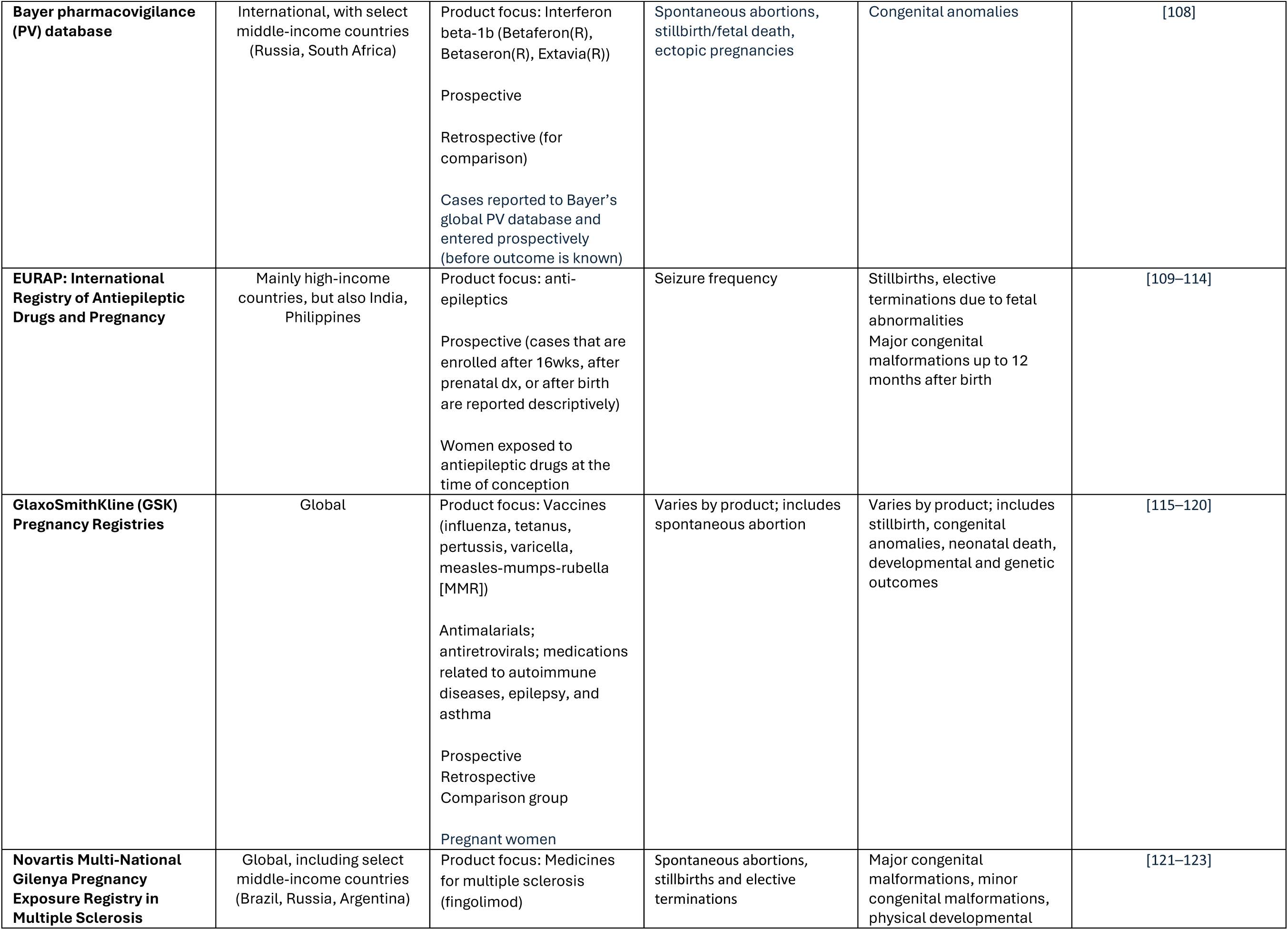
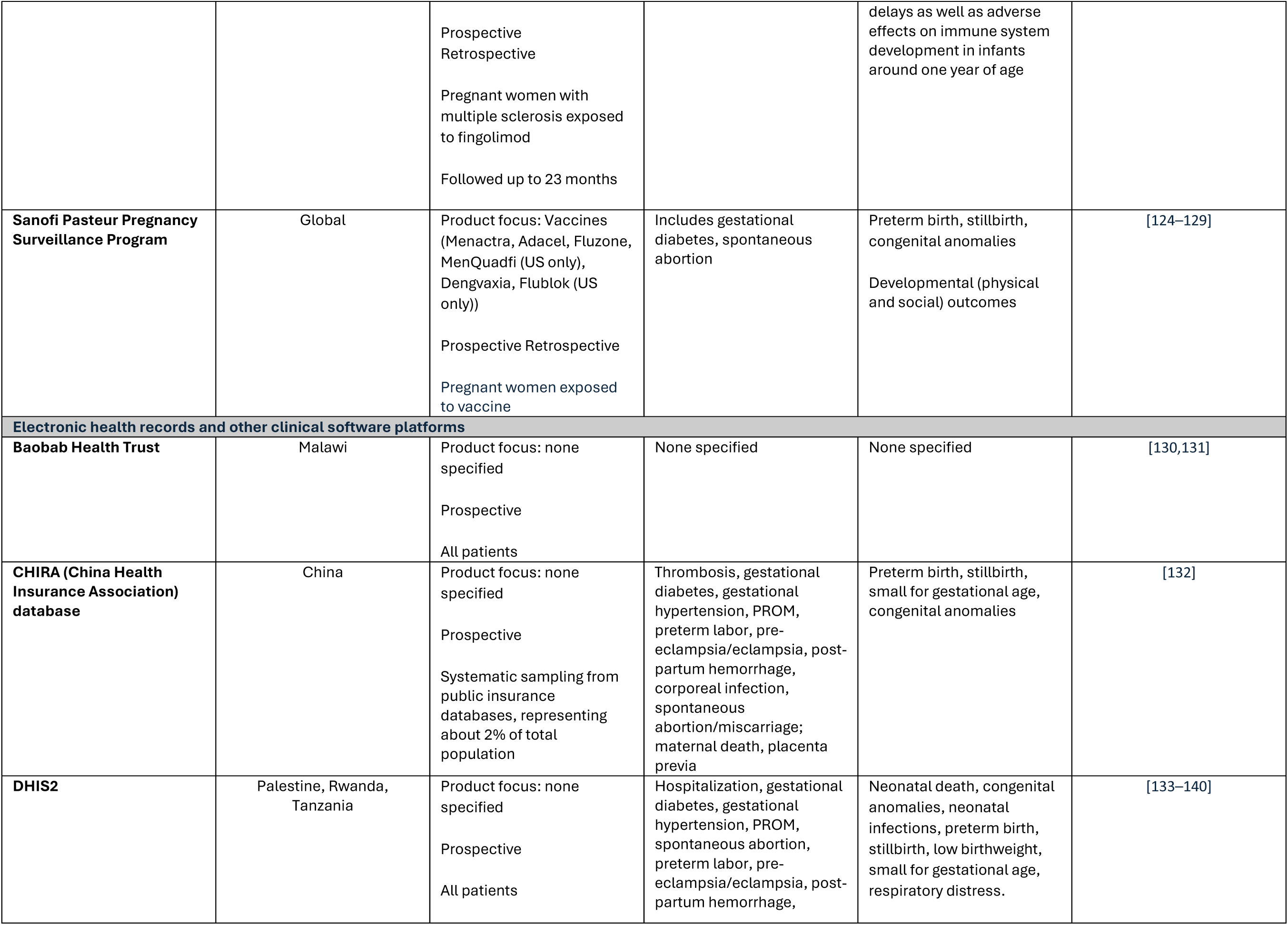
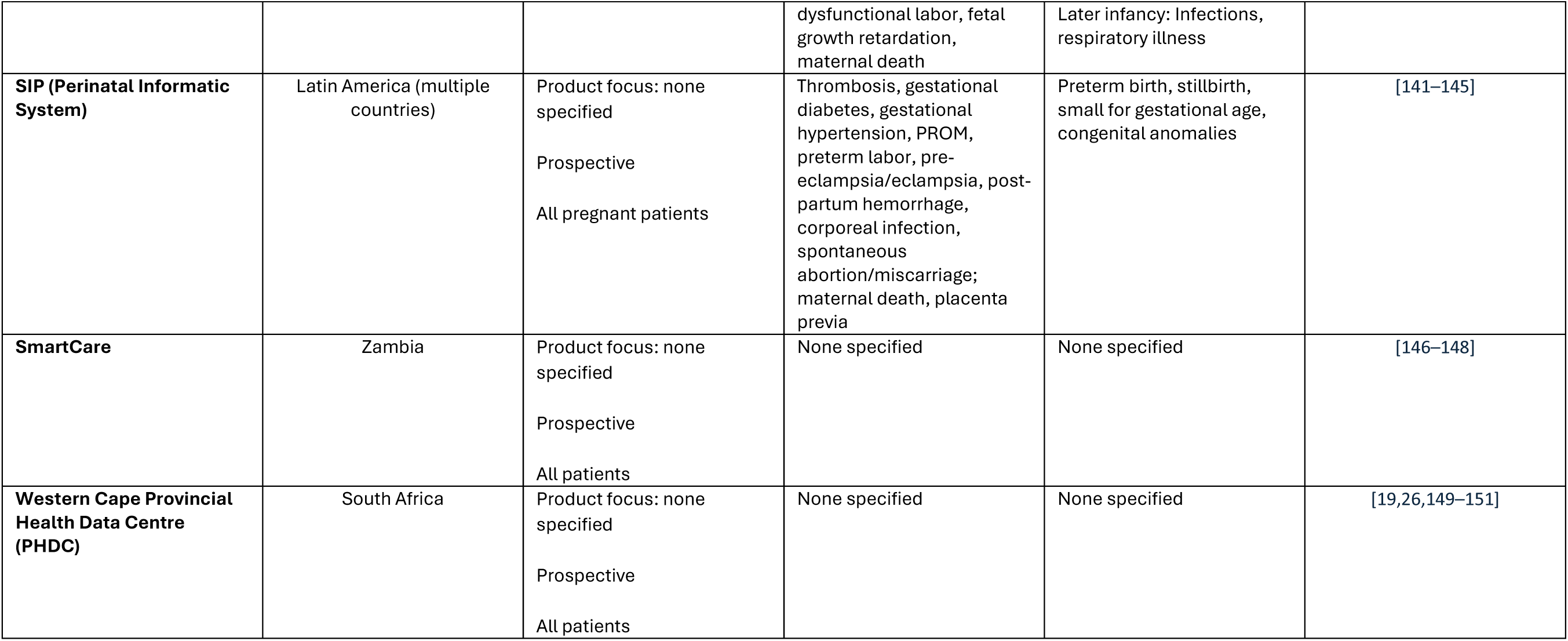
Key characteristics of identified pregnancy exposure registries and related resources.

| Resource Name | Countries | Design and Eligibility | Maternal Outcomes | Neonatal and Infant/child outcomes | References |
| --- | --- | --- | --- | --- | --- |
| **Pregnancy Exposure Registries** |  |  |  |  |  |
| **CHERISH (Children HIV Exposed Uninfected Research to Inform Survival and Health)** | South Africa | Product focus:<br>antiretrovirals<br>Prospective<br>Pregnant women with known HIV status at 24–36 weeks estimated gestational age<br>Follow-up of children to 3-5 years | Not specified | Infant and under 3-year survival; infant and under 3-year all-cause and infectious-cause hospitalization; growth and neurodevelopmental outcomes at 3–5 years of age | [1] |
| **C-VIPER (COVID-19 Vaccines International Pregnancy Exposure Registry) and PIPER (Pregistry International Pregnancy Exposure Registry)** | Global, based in the United States | Product focus: COVID-19 vaccines<br>Pregnant women exposed (C-VIPER) and not exposed (PIPER) to COVID-19 vaccines<br>Comparison group<br>Infants followed through 1 year of age | Spontaneous abortion, antenatal bleeding, gestational diabetes, gestational hypertension, intrauterine growth restriction, postpartum hemorrhage, fetal distress, uterine rupture, placenta previa, chorioamnionitis, Caesarean delivery, or COVID-19 | Major congenital malformations, low birth weight, neonatal death, neonatal encephalopathy, neonatal infections, neonatal acute kidney injury, preterm birth, respiratory distress in the newborn, small for gestational age, stillbirth, or COVID-19; infant weight, length, developmental milestones through 1 year of age | [2–5] |
| **COVID-PR (COVID-19 International Drug Pregnancy Registry)** | Global, based in the United States | Product focus: COVID-19 drugs (antivirals and monoclonal antibodies)<br>Pregnant women exposed to COVID-19 drugs<br>Comparison groups:<br>1. Pregnant women treated with another therapy for COVID-19<br>2. Pregnant women hospitalized but not treated for COVID-19 | Spontaneous abortion, intrauterine growth restriction, gestational diabetes, gestational hypertension, postpartum hemorrhage, Caesarean delivery | Major congenital malformations, low birth weight, small for gestational age, neonatal infections, stillbirth, neonatal death, preterm birth; infant weight, length, developmental milestones through 1 year of age. | [5–7] |
|  |  | Infants followed through 1 year of age |  |  |  |
| **MANGO (Measuring Adverse Pregnancy and Newborn Congenital Outcomes)** | Kenya (western) | Product focus: antiretrovirals<br>Prospective and retrospective<br>Comparison group<br>Pregnant women with known HIV status, 1:1 ratio of HIV positive to negative | Live birth, stillbirth, miscarriage, termination of pregnancy, ectopic pregnancy, molar pregnancy<br>Also includes pre-term delivery (<37 weeks gestational age) or very pre-term delivery (<32 weeks gestational age) | Congenital abnormalities on newborn surface exam (e.g., extra digit, hydrocephalus, skull defects, eyes, face, mouth/lip/palate, chest, abdomen, anus, limbs, spine (including neural tube defects), hips, genitalia, skin, etc.)<br>Also includes low birth weight, small for gestational age (<10th percentile), or very small for gestational age (<3rd percentile) | [8,9] |
| **MiMba (Malaria in Mothers and Babies) Pregnancy Registry** | Kenya, Burkina Faso | Product focus: antimalarials<br>Prospective<br>Comparison group<br>Pregnant women exposed/unexposed to antimalarials during pregnancy<br>Mothers followed through delivery<br>Infants followed through two years of age | Miscarriage, stillbirth, maternal mortality | Major congenital anomalies, neonatal mortality, low birthweight, prematurity | [10,11] |
| **REPRESENT (Xiamen Registry of Pregnant Women and Offspring)** | China (Xiamen) | Product focus: none specified<br>Retrospective<br>Mothers followed to 42 days after delivery | Preeclampsia/eclampsia, gestational diabetes, uterine rupture, postpartum hemorrhage, stillbirth, maternal death | Congenital anomalies, preterm birth, low birth weight, neonatal death | [12–14] |
|  |  | Infants followed through childhood |  |  |  |
| **UBOMI BUHLE (Understanding Birth Outcomes from Mothers and Infants, Building Healthcare by Linking Exposures) Pregnancy Exposure Registry** | South Africa (3 provinces) Incorporating the Western Cape Pregnancy Exposure Registry | Product focus: antiretrovirals<br>Prospective<br>All pregnant women attending antenatal care<br>Mothers followed through delivery<br>Infants followed through neonatal period up to discharge. | Gestational diabetes, gestational hypertension, maternal death, post-partum hemorrhage, preeclampsia / eclampsia, preterm labor, spontaneous abortion / miscarriage / pregnancy loss | Congenital anomalies on newborn surface exam, neonatal death, preterm birth, small size for gestational age /restricted fetal growth, stillbirth | [15–19] |
| **Health and Demographic Surveillance Systems and other Observational cohorts** |  |  |  |  |  |
| **CHAMPS (Child Health and Mortality Prevention Surveillance Network) pregnancy surveillance** | Global, based in the United States | Product focus: none specified<br>Prospective and retrospective<br>All pregnancies in catchment area<br>Follow-up 42 days after delivery | Maternal death, post-partum hemorrhage, pre-eclampsia and eclampsia, obstructed labor, acute infections (e.g., chorioamnionitis or sepsis), Cesarean section, miscarriage | Neonatal death, neonatal resuscitation stillbirth, low/very low birthweight | Personal communication |
| **IeDEA (International Epidemiology Databases to Evaluate AIDS)** | Multinational | Product focus: antiretrovirals<br>Various study designs<br>All populations living with or at risk for HIV<br>Variable follow-up | Variable | Variable | [20–28,8,29] |
| **INDEPTH** | Multinational | Product focus: none specified<br>Prospective<br>General population | Maternal death, gestational hypertension, fetal distress, postpartum hemorrhage, spontaneous abortion, antenatal bleeding | Neonatal death, congenital anomalies, neonatal infections, preterm birth, stillbirth, low birthweight, small for gestational age | [30–48,44] |
|  |  | Variable follow-up |  |  |  |
| **Maternal Newborn Health Registry** | Multinational | Product focus: none specified<br>Prospective<br>Pregnant women<br>Mothers followed 42 days post-partum | Maternal death, pre-eclampsia/eclampsia, gestational hypertension, fetal distress, ectopic pregnancy, postpartum hemorrhage, spontaneous abortion, antenatal bleeding, dysfunctional labor, fetal growth retardation, gestational diabetes, endometritis, chorioamnionitis, PPROM | Neonatal death, congenital anomalies, neonatal infections, preterm birth, stillbirth, low birthweight, small for gestational age, respiratory distress, neonatal seizures | [49–67] |
| **PREPARE** | Uganda | Product focus: antimalarials, antiretrovirals, COVID-19 vaccines, tetanus-diphtheria vaccine<br>Prospective<br>Pregnant women<br>Mothers and infants followed for 9 months after delivery | Serious adverse events, medically-attended events, obstetric complications | Serious adverse events, medically attended events, major congenital anomalies, developmental delay | [68] |
| **Shoklo Malaria Research Unit** | Thailand | Product focus: antimalarials<br>Prospective<br>Pregnant women<br>Mothers followed through delivery<br>Infants followed through neonatal period | Miscarriage (primary) | Major congenital malformations (secondary) | [69–71] |
| **UCLA DRC Research Program** | Democratic Republic of Congo | Product focus: none specified<br>Prospective and retrospective components | Preterm birth; stillbirth | Invasive bloodstream infection; neonatal death, congenital microcephaly, low birth weight; small for gestational age | [72,73] |
|  |  | Pregnant women |  |  |  |
|  |  | Infants followed for 28 days after delivery |  |  |  |
| **Outcomes-based registries** |  |  |  |  |  |
| **BBDSFP (Bogota Birth Defects Surveillance and Follow-up Program)** | Colombia | Product focus: none specified<br>Retrospective<br>All deliveries<br>Neonatal period | Maternal illnesses by ICD code | Congenital anomalies | [74–76] |
| **CHAMPS (Child Health and Mortality Prevention Surveillance) <u>stillbirth and neonatal and mortality surveillance</u>** | Multinational | Product focus: none specified<br>Retrospective<br>Stillbirths and neonatal deaths<br>Neonatal period | stillbirth, spontaneous abortion | Neonatal death | [47,77] |
| **CTBC (China Teratology Birth Cohort)** | China | Product focus: antiretrovirals; antimalarials; medicines related to autoimmunity, cancer, diabetes, and epilepsy<br>Prospective<br>Comparison group<br>All deliveries<br>Followed for 42 days after delivery | stillbirth, spontaneous abortion | Congenital anomalies, preterm birth, post-term birth, low birth weight, macrosomia, small for gestational age, large for gestational age, low Apgar score | [78] |
| **Eswatini Birth Defects Study** | Eswatini | Product focus: antiretrovirals<br>Retrospective<br>Comparison group | N/A | Major and minor surface congenital anomalies | [79,80] |
| **Makerere Birth Defects Surveillance Project** | Uganda | Product focus: antiretrovirals; tetanus toxoid vaccine<br>Retrospective<br>Comparison group<br>All deliveries<br>Neonatal period | Spontaneous abortion, stillbirth | Congenital anomalies; congenital infections | [81–83] |
| **Malawi Birth Defects Surveillance** | Malawi | Product focus: none specified<br>Retrospective<br>All deliveries | N/A | Major external congenital anomalies | [84,85] |
| **Tsepamo Study** | Botswana | Product focus: antiretrovirals<br>Retrospective<br>Comparison group<br>All deliveries<br>Neonatal period | Preterm delivery, stillbirth | Congenital anomalies on surface exam, preterm birth, small for gestational age, or neonatal death | [86–92] |
| **Maternal conditions-based registries** |  |  |  |  |  |
| **Kerala registry of Epilepsy and Pregnancy (KREP)** | India | Product focus: anti-epileptics<br>Prospective, enrolled in the preconception period or first trimester<br>Exposed and unexposed (comparator)<br>Pregnant women with epilepsy | Seizure frequency. Pregnancy induced hypertension, pre-eclampsia, intra-uterine growth retardation, pre- and post-term delivery, placenta previa, hydramnios, spontaneous and therapeutic abortion, intra-uterine death and stillbirth, neonatal death, postpartum hemorrhage. | Major congenital malformations; chromosomal defects. Pre-term and post-term birth. | [93–100] |
| **REBECGA Brazilian Registry of Pregnancy and Heart Disease** | Brazil | Product focus: cardiac medications | Cardiovascular complications during delivery and puerperium; obstetric complications of | Congenital malformations | [101] |
|  |  | Prospective (longitudinal) and Retrospective (cross-sectional) stages<br>Pregnant women with heart disease | pregnancy, delivery, and puerperium; Hospitalization for treating cardiovascular complications during pregnancy or puerperium; late maternal death up to 12 months after delivery. |  |  |
| **Tamil Nadu Pregnancy and Heart Disease Registry (TNPHDR)** | India | Product focus: cardiac medications<br>Prospective<br>Pregnant women with cardiac disease | Cardiac: Cardiac death, cardiac arrest, cardiac hospitalization, new or worsening heart failure, new arrhythmia, thromboembolic event, hemorrhage, aortic dissection, endocarditis, acute coronary syndrome, or cardiac intervention during pregnancy and up to 1 week post-delivery<br>Obstetric: gestational diabetes mellitus, gestational hypertension, pre-eclampsia, eclampsia / HELLP syndrome, preterm labor, premature rupture of membranes, fetal loss. | Premature birth, intra-uterine growth retardation or small-for-gestational-age and congenital heart disease or anomalies | [102] |
| **Manufacturer registries** |  |  |  |  |  |
| **APR (Antiretroviral Pregnancy Registry)** | Multinational (70-80% US) | Product focus: antiretrovirals (HIV and HBV)<br>Prospective<br>No comparison group<br>Voluntary; pregnant women exposed to antiretrovirals<br>Mothers followed through delivery<br>Infants followed through 28 days | Spontaneous abortion, miscarriage, pregnancy loss | Congenital anomalies, death, live birth, preterm birth, small size for gestational age / restricted fetal growth, stillbirth | [20,103–106,86,107] |
| **Bayer pharmacovigilance (PV) database** | International, with select middle-income countries (Russia, South Africa) | Product focus: Interferon beta-1b (Betaferon(R), Betaseron(R), Extavia(R))<br>Prospective<br>Retrospective (for comparison)<br>Cases reported to Bayer's global PV database and entered prospectively (before outcome is known) | Spontaneous abortions, stillbirth/fetal death, ectopic pregnancies | Congenital anomalies | [108] |
| **EURAP: International Registry of Antiepileptic Drugs and Pregnancy** | Mainly high-income countries, but also India, Philippines | Product focus: anti-epileptics<br>Prospective (cases that are enrolled after 16wks, after prenatal dx, or after birth are reported descriptively)<br>Women exposed to antiepileptic drugs at the time of conception | Seizure frequency | Stillbirths, elective terminations due to fetal abnormalities<br>Major congenital malformations up to 12 months after birth | [109–114] |
| **GlaxoSmithKline (GSK) Pregnancy Registries** | Global | Product focus: Vaccines (influenza, tetanus, pertussis, varicella, measles-mumps-rubella [MMR])<br>Antimalarials; antiretrovirals; medications related to autoimmune diseases, epilepsy, and asthma<br>Prospective<br>Retrospective<br>Comparison group<br>Pregnant women | Varies by product; includes spontaneous abortion | Varies by product; includes stillbirth, congenital anomalies, neonatal death, developmental and genetic outcomes | [115–120] |
| **Novartis Multi-National Gilenya Pregnancy Exposure Registry in Multiple Sclerosis** | Global, including select middle-income countries (Brazil, Russia, Argentina) | Product focus: Medicines for multiple sclerosis (fingolimod) | Spontaneous abortions, stillbirths and elective terminations | Major congenital malformations, minor congenital malformations, physical developmental | [121–123] |
|  |  | Prospective<br>Retrospective<br>Pregnant women with multiple sclerosis exposed to fingolimod<br>Followed up to 23 months |  | delays as well as adverse effects on immune system development in infants around one year of age |  |
| **Sanofi Pasteur Pregnancy Surveillance Program** | Global | Product focus: Vaccines (Menactra, Adacel, Fluzone, MenQuadfi (US only), Dengvaxia, Flublok (US only))<br>Prospective Retrospective<br>Pregnant women exposed to vaccine | Includes gestational diabetes, spontaneous abortion | Preterm birth, stillbirth, congenital anomalies<br>Developmental (physical and social) outcomes | [124–129] |
| **Electronic health records and other clinical software platforms** |  |  |  |  |  |
| **Baobab Health Trust** | Malawi | Product focus: none specified<br>Prospective<br>All patients | None specified | None specified | [130,131] |
| **CHIRA (China Health Insurance Association) database** | China | Product focus: none specified<br>Prospective<br>Systematic sampling from public insurance databases, representing about 2% of total population | Thrombosis, gestational diabetes, gestational hypertension, PROM, preterm labor, pre-eclampsia/eclampsia, post-partum hemorrhage, corporeal infection, spontaneous abortion/miscarriage; maternal death, placenta previa | Preterm birth, stillbirth, small for gestational age, congenital anomalies | [132] |
| **DHIS2** | Palestine, Rwanda, Tanzania | Product focus: none specified<br>Prospective<br>All patients | Hospitalization, gestational diabetes, gestational hypertension, PROM, spontaneous abortion, preterm labor, pre-eclampsia/eclampsia, post-partum hemorrhage, | Neonatal death, congenital anomalies, neonatal infections, preterm birth, stillbirth, low birthweight, small for gestational age, respiratory distress. | [133–140] |
|  |  |  | dysfunctional labor, fetal growth retardation, maternal death | Later infancy: Infections, respiratory illness |  |
| **SIP (Perinatal Informatic System)** | Latin America (multiple countries) | Product focus: none specified<br>Prospective<br>All pregnant patients | Thrombosis, gestational diabetes, gestational hypertension, PROM, preterm labor, pre-eclampsia/eclampsia, post-partum hemorrhage, corporeal infection, spontaneous abortion/miscarriage; maternal death, placenta previa | Preterm birth, stillbirth, small for gestational age, congenital anomalies | [141–145] |
| **SmartCare** | Zambia | Product focus: none specified<br>Prospective<br>All patients | None specified | None specified | [146–148] |
| **Western Cape Provincial Health Data Centre (PHDC)** | South Africa | Product focus: none specified<br>Prospective<br>All patients | None specified | None specified | [19,26,149–151] |

## Notes

### Competing Interest Statement

The authors have declared no competing interest.

### Clinical Protocols

https://bmjopen.bmj.com/content/13/5/e070543

